# Single-nucleus transcriptome-wide association study of human brain disorders

**DOI:** 10.1101/2024.11.04.24316495

**Authors:** Sanan Venkatesh, Zhenyi Wu, Marios Anyfantakis, Christian Dillard, N.M. Prashant, David Burstein, Deepika Mathur, Roman Kosoy, Chris Chatzinakos, Bukola Ajanaku, Fotis Tsetsos, Biao Zeng, Aram Hong, Clara Casey, Marcela Alvia, Zhiping Shao, Stathis Argyriou, Karen Therrien, VA Million Veteran Program, PsychAD Consortium, Tim Bigdeli, Pavan Auluck, David A. Bennett, Stefano Marenco, Vahram Haroutunian, Kiran Girdhar, Jaroslav Bendl, Donghoon Lee, John F. Fullard, Gabriel E. Hoffman, Georgios Voloudakis, Panos Roussos

**Affiliations:** Department of Psychiatry, Icahn School of Medicine at Mount Sinai, New York, NY, USA; Center for Disease Neurogenomics, Icahn School of Medicine at Mount Sinai, New York, NY USA; Friedman Brain Institute, Icahn School of Medicine at Mount Sinai, New York, NY, USA; Department of Genetics and Genomic Science, Icahn School of Medicine at Mount Sinai, New York, NY, USA; Mental Illness Research, Education, and Clinical Center (VISN 2 South), James J. Peters VA Medical Center, Bronx, NY, USA; Center for Precision Medicine and Translational Therapeutics, James J. Peters VA Medical Center, Bronx, NY, USA; Department of Psychiatry and Behavioral Sciences, SUNY Downstate Health Sciences University, Brooklyn, NY, USA; Institute for Genomics in Health (IGH), SUNY Downstate Health Sciences University, Brooklyn, NY, USA; VA New York Harbor Healthcare System, Brooklyn, NY, USA; Department of Epidemiology and Biostatistics, School of Public Health, SUNY Downstate Health Sciences University, Brooklyn, NY, USA; Human Brain Collection Core, National Institute of Mental Health-Intramural Research Program, Bethesda, MD, USA; Rush Alzheimer’s Disease Center, Rush University Medical Center; Department of Artificial Intelligence and Human Health, Icahn School of Medicine at Mount Sinai, New York, NY, USA

## Abstract

Neuropsychiatric and neurodegenerative disorders exhibit cell-type-specific characteristics^1–8^, yet most transcriptome-wide association studies have been constrained by the use of homogenate brain tissue^9–11^, limiting their resolution and power. Here, we present a single-nucleus transcriptome-wide association study (**snTWAS**) leveraging single-nucleus RNA sequencing of over 6 million nuclei from the dorsolateral prefrontal cortex of 1,494 donors across three ancestries—European, African, and Admixed American. We constructed ancestry-specific single-nucleus-derived transcriptomic imputation models (**snTIMs**) including up to 27 non-overlapping cellular populations, enhancing the resolution of genetically regulated gene expression (**GReX**) in the brain and uncovering novel gene-trait associations across 12 neuropsychiatric and neurodegenerative traits. Our snTWAS framework revealed cell-type-specific dysregulation of GReX, identifying over 4,000 novel gene-trait associations not detected by bulk tissue approaches. By applying these snTIMs to the Million Veteran Program, we validated major findings and explored the pleiotropy of cell-type-specific GReX, revealing cross-ancestry concordance and fine-mapping causal genes. This approach enhances the discovery of biologically relevant pathways and gene targets, highlighting the importance of cell-type resolution and ancestry-specific models in understanding the genetic architecture of complex brain disorders.

## Main

The genetic architecture of neuropsychiatric and neurodegenerative disorders (**NPD/NDD**s) is complex, with a significant portion of their heritability attributed to common genetic variants within non-coding regions of the genome. Genome-wide association studies (**GWAS**) have successfully associated numerous variants with NPDs and NDDs, yet pinpointing causal genes and understanding the underlying biological mechanisms remains challenging. Transcriptome-wide association studies (**TWAS**) have emerged as a powerful approach to bridge this gap by integrating GWAS data with predictive models of genetically regulated gene expression (**GReX**), thereby linking genetic variants to gene expression changes that may drive disease risk^12–14^. Traditional TWAS efforts, however, have predominantly utilized bulk tissue RNA sequencing^7,15–17^, which masks the cell-type-specificity of gene expression and genetic regulation^1–3,5–7,18–21^. Given the heterogeneous cellular composition of the human brain and its high level of transcriptional diversity encompassing both cells with distinct embryologic origins and transcriptionally similar but functionally distinct subtypes of neurons^22^, such an approach is suboptimal for studying NPDs and NDDs, where risk variants often exhibit cell-type-specific effects. Recent evidence underscores the importance of cell-type-specific investigations, as different cell-types in the brain contribute uniquely to the pathogenesis of disorders such as schizophrenia (**SCZ**)^1–6^, Alzheimer’s disease (**AD**)^7,8^, and major depressive disorder (**MDD**)^23,24^.

The PsychAD Consortium (**Supplementary Notes “PsychAD dataset”**)^22,25^ generated a population-level single-nucleus RNA sequencing (**snRNA-seq**) dataset comprising over 6 million nuclei isolated from the dorsolateral prefrontal cortex (**DLPFC**) of 1,494 individual donors. Using this data, we constructed single-nucleus-derived transcriptomic imputation models (**snTIM**s) for 32 (27 non-overlapping) cellular populations, enabling us to capture GReX with unprecedented resolution. These snTIMs were then leveraged to perform TWAS for 12 NPD and NDDs, revealing novel gene-trait associations (**GTA**s) that are specific to individual cell-types. In addition to identifying novel associations, we validated our findings through a large-scale phenome-wide association study (**PheWAS**) in approximately 600,000 individuals from the Million Veteran Program (**MVP**). This validation not only confirmed the robustness of our results but also highlighted the pleiotropic effects of cell-type-specific GReX across various neurological, mental health, and sensory organ disorders. Importantly, our ancestry-specific models enabled cross-ancestry comparisons, uncovering shared genetic architectures and enhancing the fine-mapping of causal genes.

### Brain snTIMs capture brain GReX with cell-type specificity across ancestries

We developed an analytical framework to interrogate brain cell-type-specific GReX contributions in NPD and NDDs (Extended Data Fig. 1). By utilizing genotype and snRNA-seq data from the PsychAD Consortium^22,25^, we trained 94 snTIMs across 3 levels of cell-type hierarchy including 32 cellular populations in the DLPFC (Extended Data Fig. 2; Table S1). At the highest level of resolution, we obtained 27 non-overlapping cellular populations (4 classes and 23 subclasses). We were able to reliably (R^2^_Cross Validation (**CV**)_ ≥ 0.01, p_CV_ ≤ 0.05, 10-fold model training cross-validation) impute 20,189 (59.9%), 18,742 (55.6%) and 13,923 (41.3%) of the 33,688 assayed genes within the PsychAD dataset^22,25^ (Extended Data Fig. 3A; Tables S2-7) in individuals of European (**EUR**; n = 920), African (**AFR**; n = 321) and Admixed American (**AMR**; n = 118) ancestry, respectively.

EUR PsychAD snTIMs confidently predict more genes than a similarly sized (n = 924) EUR homogenate transcriptomic imputation model (**TIM**) for the same brain region (DLPFC^26^ EpiXcan^9^ TIM^27^, hereafter referred to as “Bulk”) across “snBulk” (all cells pooled together), “Class” and “Subclass” (best gene model trained at the class and subclass level, respectively) (Fig. 1A). Specifically, snTIMs were able to impute expression for 11,266 genes which were not imputed in Bulk, and outperformed the bulk model in more than 67% of common imputable genes (6,023/8,923) (sign-test p-value = 1.98 × 10^-2,683^). In summary, snTIMs can identify more genes across the R^2^_CV_ spectrum than Bulk at both the snBulk level and across individual cell-type-specific snTIMs (Fig. 1B). Conversely, 1,364 genes are only imputable in Bulk and not in snTIMs. These discrepancies can be partially explained by differences in sequencing technology and the detection of extranuclear RNA in bulk homogenate, as indicated by the capture of different species of long non-coding RNAs (**lncRNAs**) which are known to have preferential subcellular localizations^28^. Uniquely identified transcripts of both Bulk and snTIMs were enriched for lncRNAs (Fisher’s exact test, odds ratio (**OR**) = 1.38 and 5.74, p-value = 4.68 × 10^-5^ and 2.80 × 10^-496^, respectively). Within EUR snTIMs, increasing cellular resolution to the class and subclass levels allows for the imputation of 4,465 genes not imputable in snBulk, whereas class and subclass levels can impute approximately the same number of genes (17,261 and 16,518, respectively), despite the higher resolution of the latter (Table S8).

**Fig. 1.**
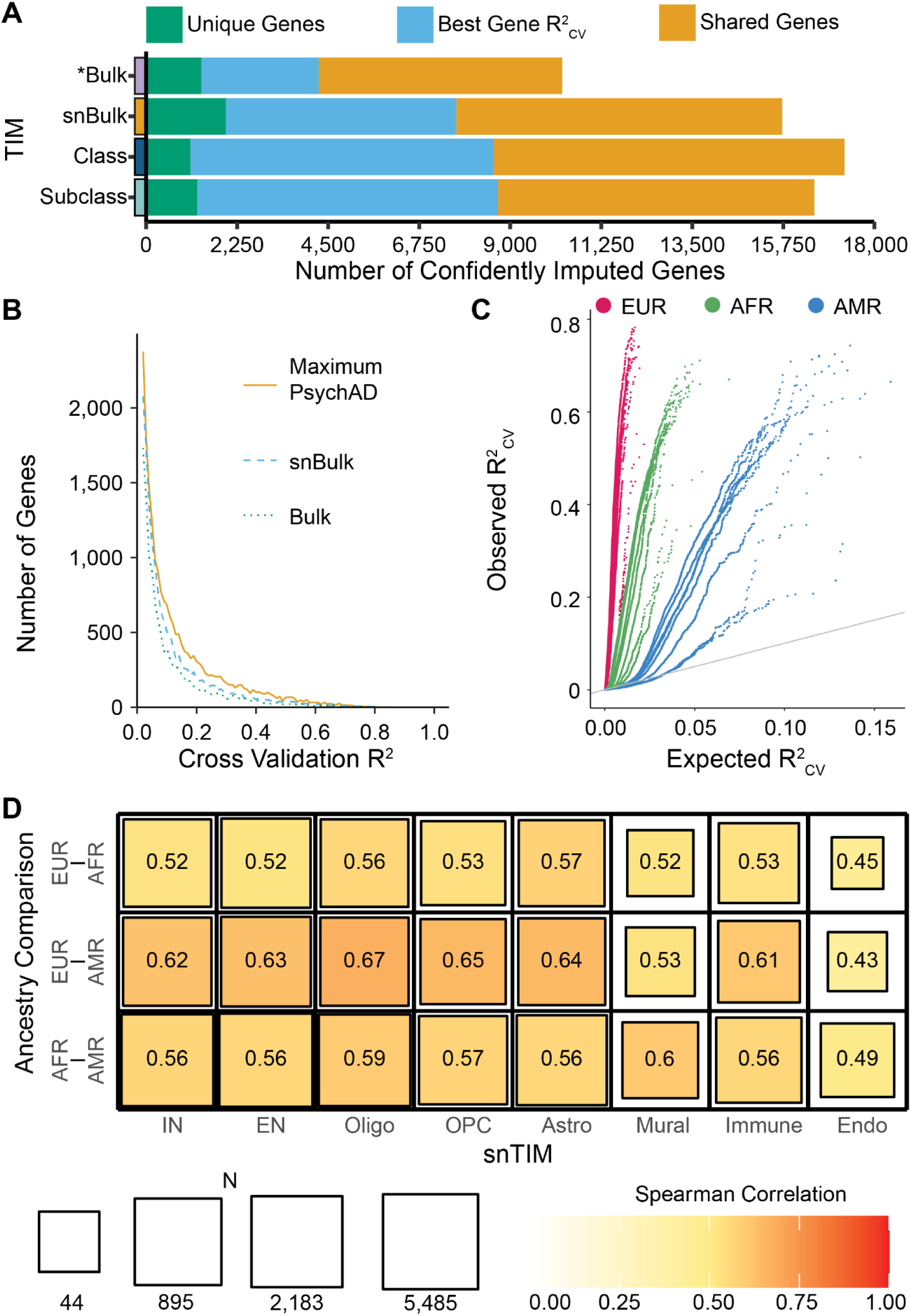
snTIM performance across cell-types and ancestries. **A,** Gene imputability across different cell-type resolutions. Bar lengths represent the number of reliably imputable genes. For each category, genes are split into three groups: uniquely captured genes (unique genes), best predicted genes (best gene R^2^_CV_: SNP predictors explain a higher percentage of the gene expression variance in this category), shared genes (imputable genes that are better captured in other categories). “Bulk” only assesses imputable genes among the 33,688 assayed genes in the psychAD dataset. “Class” and “Subclass” utilize the best gene model (max R^2^_CV_) among all participating snTIMs for that level. (*) Bulk is compared against the aggregate of all snTIMs (snBulk, class and subclass aggregates); snTIMs are compared against the other snTIM categories. **B,** Distribution of R^2^_CV_ among EUR snTIMs. Maximum PsychAD refers to the distribution of the maximum R^2^_CV_ per gene across all EUR snTIMs (snBulk, Class and Subclass). **C,** Power comparison of snTIMs across ancestries. Quantile-quantile plot of the observed against expected performance (R^2^_CV_) for every class-level model across 3 genetic ancestries. EUR: European; AFR: African; AMR: Admixed-American. **D,** Heatmap of cross-ancestry Spearman’s correlation coefficients (ρ) of R^2^_CV_ at the class level. For each cell-type, Spearman correlation is assessed among all shared genes in the ancestry comparison pair. Full correlation statistics are described in Table S16.

We explored the major factors associated with EUR snTIMs and found that the number of individual donors and median nuclei per cellular population per individual explain 65.7% of variance in the number of imputable genes (Extended Data Fig. 3B; Table S9). Previously, we found that sample size was the main driver for unique GTA discovery in tissue homogenates^9^. However, in single-cell experiments, when sequencing depth is not a limiting factor, snTIM performance also heavily depends on the number of cells being profiled. Consequently, there is a fine balance between increasing resolution and retaining information when using standard snRNA-seq approaches. Specifically, using a pseudobulk approach reduces measurement noise, while increasing statistical power from sparse single cell measurements^29–32^. Thus, for low abundance transcripts, imputation performance improves when we pool more cells together; for example, imputable genes by EUR snBulk that were not predictable via finer cell-type resolution (n = 1,970) were less abundant (48 mean counts per individual versus 2,495; Kruskal-Wallis p-value = 1.87 × 10^-422^).

Finally, we compared the EUR, AFR and AMR snTIMs. Unsurprisingly, given the differences in the donor sample size, the number of imputable genes (Extended Data Fig. 3A; Tables S2-7) and gene expression variation explained by *cis* genetic variants (R^2^_CV_) were higher in EUR, followed by AFR and AMR (Fig. 1C; Table S8). Despite differences in power, AFR and AMR snTIMs reliably imputed an additional 2,378 and 1,558 genes, respectively, that were not imputable in EUR (Table S10-15). For all shared gene-cell-type models, we observed strong R^2^ correlation across ancestries (EUR-AMR: r = 0.60, p = 2.32 × 10^-2,598^; EUR-AFR: r = 0.56, p = 1.26 × 10^-5,011^; AFR-AMR: r = 0.51, p = 7.86 × 10^-1,456^ Spearman’s correlation analysis; Supplementary Fig. 1) that is consistent across cell-types (Fig. 1D; Supplementary Fig. 2A), suggesting that the extent to which expression for a given gene is under genetic regulation is similar across ancestries. Our analysis may underestimate the magnitude of the correlation due to power insufficiency; notably, we observed approximately half the number of shared gene-cell-type models in ancestry comparisons that included the least powered AMR models (Jaccard index is 0.19 and 0.22 for EUR-AMR and AFR-AMR, respectively) when contrasted against the EUR-AFR comparison (Jaccard index = 0.41) (Supplementary Fig. 1). Moreover, there are fewer shared gene models at the subclass level (Supplementary Fig. 2B) despite demonstrating similar correlation (Supplementary Fig. 2A), most notably for the AFR and AMR snTIMs (Table S16).

### Brain snTWAS increases power for both known and novel gene-trait association discovery

To understand transcriptional dysregulation associated with NPD and NDDs, we used matched ancestry EUR snTIMs to perform summary-level snTWAS (**S-snTWAS**) on a set of 12 EUR GWAS summary statistics (Table S17 and Data S1). For each trait, we found the greatest number of FDR^33^ significantly associated genes in the subclass level (4,953 unique genes across all 12 traits), followed by Class (4,183) and snBulk (2,472), demonstrating that increased cell-type resolution contributes more significant GTAs (Fig. 2A; Supplementary Fig. 3). More genes were identified in Subclass despite the greater number of unique imputable genes for Class (Fig. 1A), suggesting the greater importance of finer resolution versus the number of imputable genes. Unsurprisingly, across NPD and NDDs, we found a similar number of associated genes in snBulk (2,472) and Bulk (2,287).

**Fig. 2.**
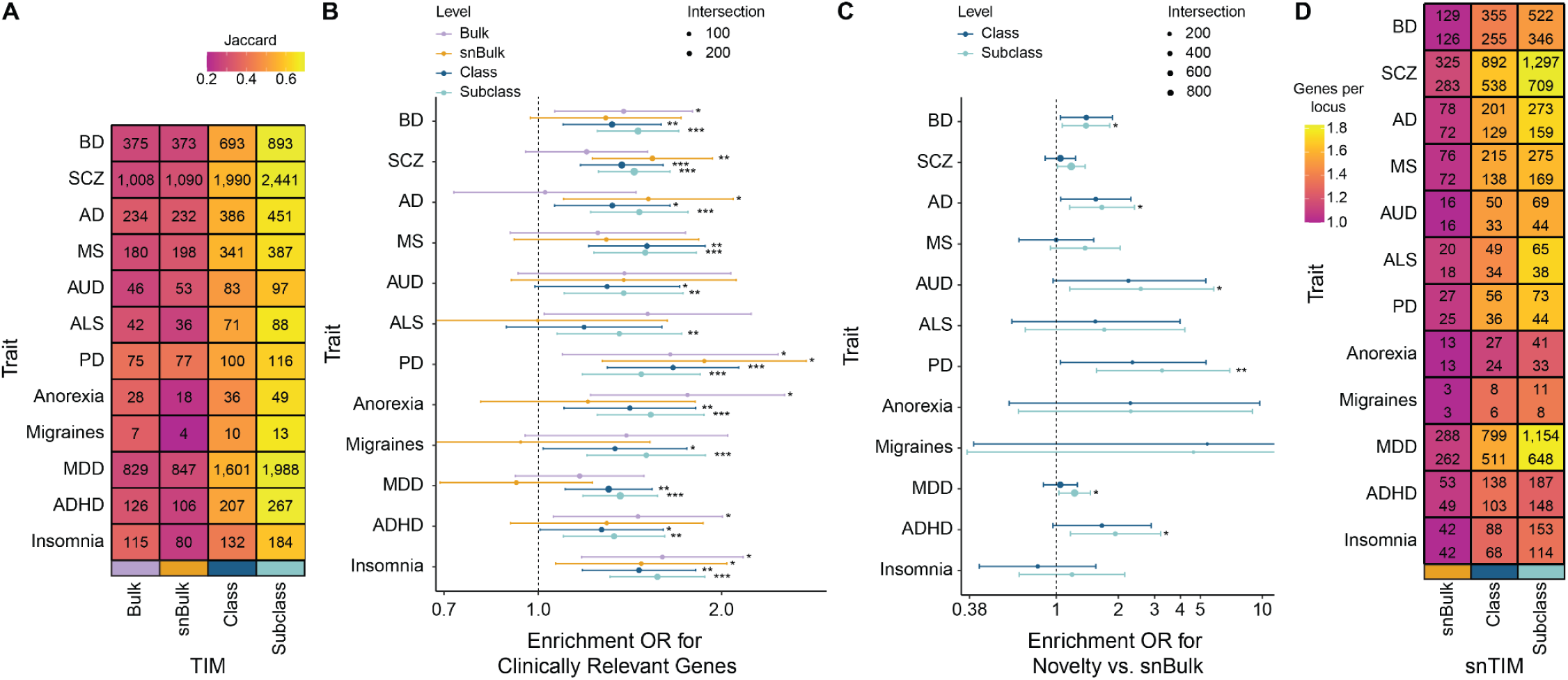
Discovery power of summary-level snTWAS in EUR. **A,** Heatmap of S-TWAS significant gene-trait associations by cell-type hierarchy level. The number within each cell represents the number of significant gene-trait associations (GTAs; FDR^33^ for multiple testing correction) within each category. Color denotes the Jaccard index 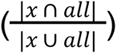 of the significant genes identified at the specific cell-type category (*x*) against the union of all significant trait-associated genes across all categories (Bulk, snBulk, Class and Subclass). **B,** Enrichment for clinically relevant genes comprising genes known to be associated with CNS-related neurological and behavioral/psychiatric symptoms (see Methods) across different cellular resolution levels. **C,** Enrichment for novel to known gene identification. Forest plot demonstrating preferential identification of “novel” vs. known GTAs in Class and Subclass versus snBulk. Novel genes are considered the genes that are not identified in Bulk TWAS and MAGMA analyses. **D,** Heatmap of S-snTWAS gene fine-mapping. The numbers at the top and bottom of each cell indicate the count of fine-mapped genes and loci after FDR multiple testing correction, respectively. Color indicates the number of genes per locus. Fine-mapping was performed with FOCUS for each cell-type. Asterisks indicate significance (* FDR ≤ 0.05, ** FDR ≤ 0.01, *** FDR ≤ 0.001).

To validate our S-snTWAS findings, we utilized two different cohorts. First, we compared our microglia subclass (Subclass-MG; n = 904) snTIM with a EUR TIM created using bulk RNA-seq data from microglia isolated by fluorescence activated cell sorting^7,34,35^ (**FACS-MG**; n = 271). Even though gene expression data for these largely independent models (22 donors in common) was obtained using different methods, AD (selected for its association with microglia^7,8^) S-TWAS applied to both TIMs exhibited a very strong correlation (Pearson’s r = 0.84, p = 2.25×10^-263^; Extended Data Fig. 4A). Second, we compared our snTIMs with external snTIMs^24^. We matched cell-types between cohorts at the class level (Table S18), and performed correlation analysis of the respective S-snTWAS using MDD GWAS summary statistics^36^. Despite the utilization of different cohorts (PsychAD vs. ROSMAP), TIM creation methods (PrediXcan vs. FUSION^37^) and sample sizes (920 vs. 424), we observed strong correlation (Pearson’s r = 0.764, p = 1.43×10^-3,308^) between both sets of snTIMs (Extended Data Fig. 4B). Furthermore, for commonly identified associations, we observed a 24% increase in power^9^ utilizing PsychAD snTIMs.

To determine the extent to which the higher number of GTAs identified by using cell-type-specific TWAS are clinically relevant, we performed gene set enrichment analysis (**GSEA**) for genes known to be associated with CNS-related neurological and behavioral/psychiatric symptoms as indicated by the Online Mendelian Inheritance in Man (**OMIM**) database^38^. Bulk and snBulk analyses revealed significant enrichment for genes linked to 5, and 4 of the 12 NPDs and NDDs analyzed, respectively, while Class and Subclass captured 11 and all 12, respectively (Fig. 2B; Table S19), further demonstrating the superior performance of cell-type-specific snTWAS.

Since our prior analysis suggests a functional role for the new GTAs, we next investigated the propensity for different cellular resolutions to identify “novel” vs. “known” GTAs. We classified significant genes as “novel” if the GTA was not identified by Bulk S-TWAS (Table S20) or MAGMA gene-based analysis (Table S21)^39,40^. Across all considered traits and snTIMs, we found that 49.4% of our significant GTAs are novel (4,274/8,656; Table S22 and S23). In addition, subclass level snTIMs are more likely to identify novel vs. known GTAs (Fig. 2C; significant enrichment in 6/12 traits; Fisher’s exact test; Table S24) than snBulk. The highest novel GTA enrichment was observed at the Subclass-EN L2-3-IT, Class-OPC and Subclass-Adaptive cellular populations, and among traits for ADHD, Anorexia, Migraines, and Parkinson’s disease (**PD**) (Supplementary Fig. 4; Table S22). A higher ratio for novel gene identification at single cell resolution suggests that GTAs may be “masked” in bulk analyses, especially those that are differentially regulated among cell-types.

Due to causal eQTL sharing or linkage disequilibrium (**LD**) between eQTLs^41^, TWASs frequently identify multiple GTAs per locus^14,42^. Thus, to identify putatively causal GTAs per cell-type, we performed probabilistic fine-mapping with FOCUS^42^ and retained, on average, 64.7% of the GTAs (Fig. 2D; Table S25 and S26). With individual-cell-type FOCUS analysis, one gene was fine-mapped (posterior inclusion probability (**PIP**) ≥ 0.5) in most loci (Supplementary Fig. 5A). At the aggregate level, we observed that the genes/loci ratio increased in the class and subclass levels, suggesting that differential fine-mapping of genes across cellular populations within the same locus warrants further investigation. We note that performing multi-cell-type vs. individual-cell-type fine-mapping among class snTIMs showed, on average, a similar but lower increase in fine-mapped genes per locus (5.15%) (p = 5.60 × 10^-5^; Kruskal-Wallis test) when compared to the class aggregate (40.27%) (p = 1.53 × 10^-8^; Kruskal-Wallis test) (Supplementary Fig. 5B; Table S27). Expectedly, multi-cell-type vs. individual-cell-type fine-mapping better prioritized genes in known, trait-relevant, cell populations.

### Summary level TWAS identifies cell-type specific disease signatures

To formally estimate the degree of cell-type specificity of GTAs within each trait, we utilized multivariate adaptive shrinkage (**mash**^43^) to obtain posterior probabilities of cell-type-specific GTAs. Across all considered traits, we found 18.4% (1,029/5,604) of GTAs have cell-type specific effects (Table S28). Of those, we found 36.2% were present in a single cell-type (Fig. 3A). At the class level, the most cell-type-specific GTAs were found in the Immune class across various traits (Fig. 3B; Extended Data Fig. 5A; Supplementary Fig. 6; Table S29). Among the top hits, we also note high cell-type specificity for *CACNA1C* in Class-IN in SCZ^44^ (Extended Data Fig. 5A; Supplementary Fig. 6; Table S29).

**Fig. 3.**
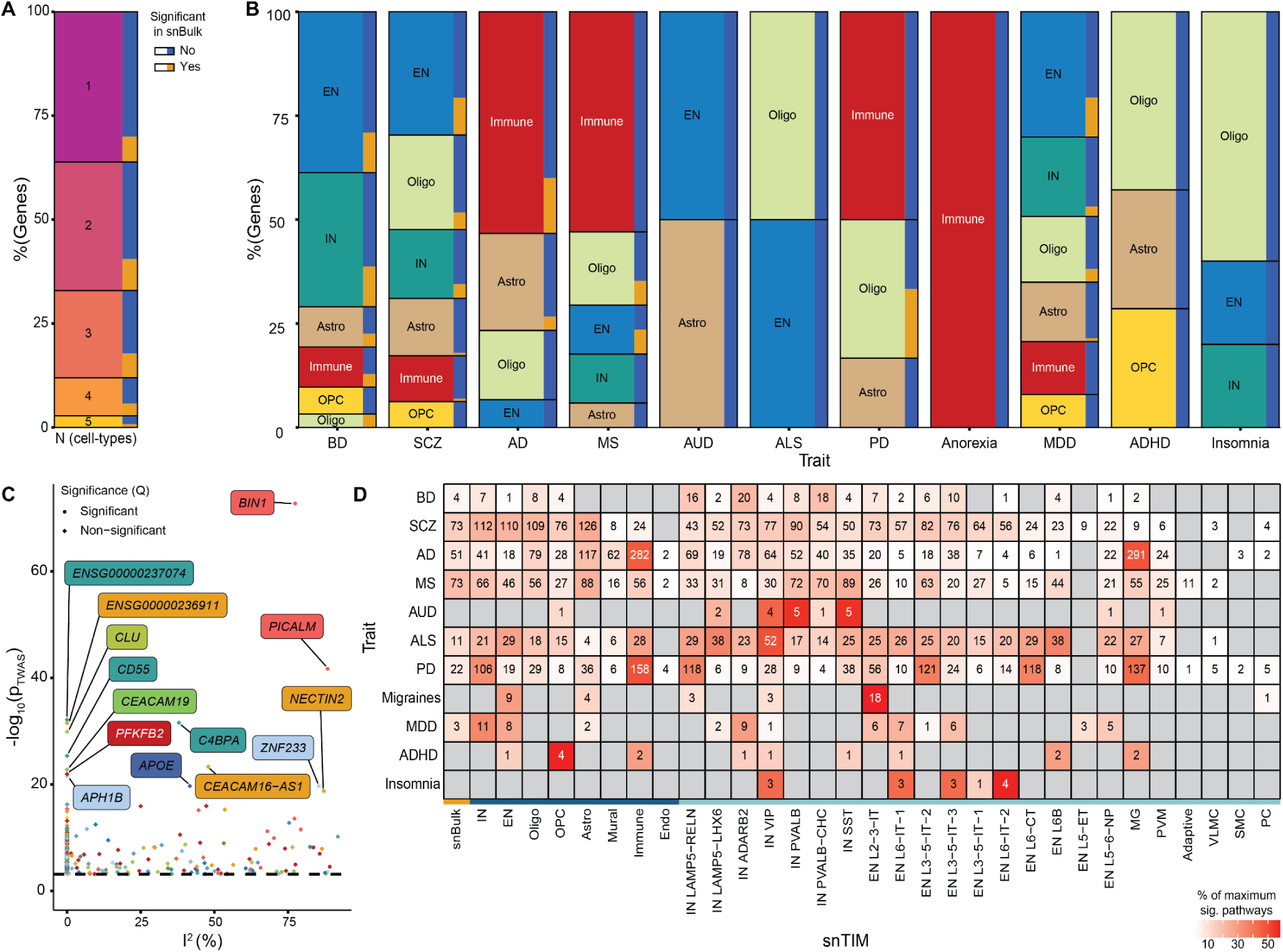
Cell-type specificity in snTWAS. **A,** Multivariate adaptive shrinkage (mash) was applied to the S-snTWAS results to jointly test GReX effects at the class level for all traits. Relative frequency stacked bar charts visualize the percentage of genes with significant post-mash GTAs in 1, 2, 3, 4 and 5 cell-types (left panel) and their respective breakdown to GTAs either significant (orange) or not (blue) in snBulk S-snTWAS. Endo and Mural classes were excluded from the mash analysis due to high sparsity in comparison to other class-level snTIMs (IN, EN, Oligo, OPC, Astro, Immune). Only S-snTWAS GTAs that were MASH significant (local false sign rate ≥ 0.95) in at least 1 out of 6 cell-types and not MASH significant in all 6 cell-types are visualized. **B,** Relative frequency stacked bar charts to visualize the breakdown of post-mash cell-type-specific GTAs (GTA only found in 1 cell-type) from (A). **C,** Gene association effect size heterogeneity (I^2^) across cell-types for AD. Significance denotes the presence of different effects of individual cell-types in the analysis (Cochran’s Q test). Only genes with a significant GTA are visualized. Color corresponds to cell-type (Extended Data Fig. 2). **D,** LD-aware snTWAS competitive pathway enrichment. The number in each cell represents the number of FDR^33^-significant (corrected across all snTIMs) pathways after hierarchical pruning (see Methods). Color represents the proportion of all significant pathways identified in each cellular population for each trait.

Approximation of S-TWAS GTA effect sizes with existing methods^37,45^ hinders our ability to assess the cell-type heterogeneity across GTAs independent of power considerations. To overcome this limitation, we imputed GReX across the EUR population of the Million Veteran Program (**MVP**) and performed individual-level snTWAS (**I-snTWAS**) analysis. Subsequently, the I-snTWAS was used to obtain NPD/NDD-specific heterogeneity scores for each gene-trait combination across all imputable cell-types as proxied by the I^2^ value^46^ (ranging from 0% to 100% for minimal to very high heterogeneity, respectively; Table S30). In AD, our top gene, *BIN1*, had one of the largest observed I^2^ (77.36%; Fig. 3C; Table S30; Supplementary Fig. 7) due to the variation of its effect sizes across cell-types (Supplementary Fig. 8), largely driven by cell-type specific single-nucleus expression quantitative trait loci (**sn-eQTL**; Extended Data Fig. 5B). Conversely, other genes, including *CLU*, exhibited consistent effect sizes (*CLU-AD* I^2^ = 0%; 11 imputable cell-types; Fig. 3C).

We next used an LD-aware competitive pathway enrichment method^47^ to determine relevant biological pathways perturbed in our S-snTWAS analyses and to characterize their cell-type specificity. Overall, increasing cellular resolution in TWAS leads to the identification of more perturbed pathways (Fig. 3D; Data S2; Supplementary Fig. 9). This is particularly important for traits such as AD and alcohol use disorder (**AUD**), where snTWAS analysis for microglia and inhibitory neuronal cellular populations, respectively, leads to relevant biological insights. For example, in AD, one of the top implicated pathways, primarily driven by dysregulation in *BIN1* and *APOE*, was Tau Protein Binding (GO:0048156) in Class-Immune (FDR^33^ = 3.76 × 10^-62^) and Subclass-MG (FDR = 5.92 × 10^-66^) (Extended Data Fig. 5C). Interestingly, shared AD pathways across cell-types were driven by different genes; the Positive Regulation of Neuron Death category (GO:1901216) was primarily driven by *PICALM*, *APOE* and *CASP2* in Class-Immune (FDR = 2.07 × 10^-37^), whereas *CLU* was an additional driver for other cellular populations (Classes: Astro, and Oligo; Subclasses: IN ADARB2). Similarly, the Regulation of APP Catabolic Process (GO:1902991), was driven by *PICALM*, *APOE*, and *SORL1* in Class-Immune (FDR = 2.53 × 10^-43^) and Subclass-MG (FDR = 1.80 × 10^-41^), and additionally driven by *ABCA7* in Class-Oligo (FDR = 3.35 × 10^-9^) and Subclass-IN PVALB (FDR = 1.03 × 10^-3^). Likewise, in SCZ, a trait demonstrating broad genetically driven biological pathway dysregulation across many cellular populations (Fig. 3B), one of the strongest hits, driven primarily by *CACNA1C* and *GPM6A*, was Divalent Inorganic Cation Transmembrane Transporter Activity (GO:0072509). Interestingly, this was implicated only in Subclass-IN ADARB2 (Extended Data Fig. 5D; FDR = 6.38 × 10^-13^), providing a putative cellular context for a known clinical biomarker of SCZ^48^. Of note, most of the aforementioned genes were also fine-mapped in the relevant cell-types (Table S31).

### Cell-type specificity enhances cross-disorder investigations

Given the high comorbidity^49,50^ and genetic correlation^51^ among NPDs and NDDs, we aimed to characterize the cell-type-specificity of shared biology across traits. In every pairwise trait combination, we queried the overlap of all gene-cell-type combinations (Fisher’s exact test), and found FDR^33^-significant enrichment in ∼68% (45/66) of trait combinations (Fig. 4A; Table S32). To address potential inflation resulting from gene representation in multiple cell-types, we note comparable findings when looking at shared significant genes regardless of cell-type (Supplementary Fig. 10A; Table S33). Genes shared among strongly genetically correlated disorders tend to have increasing concordance of effects with decreasing p-value thresholds (Supplementary Fig. 10B; Supplementary Fig. 11), and demonstrate strong correlation in a progressive thresholding correlation analysis (**PTCA;** see Methods; Supplementary Fig. 10C). Utilizing the same approach on our pathway analysis summary statistics, we similarly observed shared biology between NPDs and NDDs at the pathway level. Across all tested traits, ∼21% (14/66) of trait combinations demonstrated FDR-significant sharing enrichment of significant pathway-cell-type combinations (Fig. 4A; Table S34) with similar results when considering pathways identified in any cell-type (Supplementary Fig. 10A; Table S35). Similarly, concordance evaluation (Supplementary Fig. 10B) and PTCA (Supplementary Fig. 10C) support, on average, shared direction of pathway dysregulation.

**Fig. 4.**
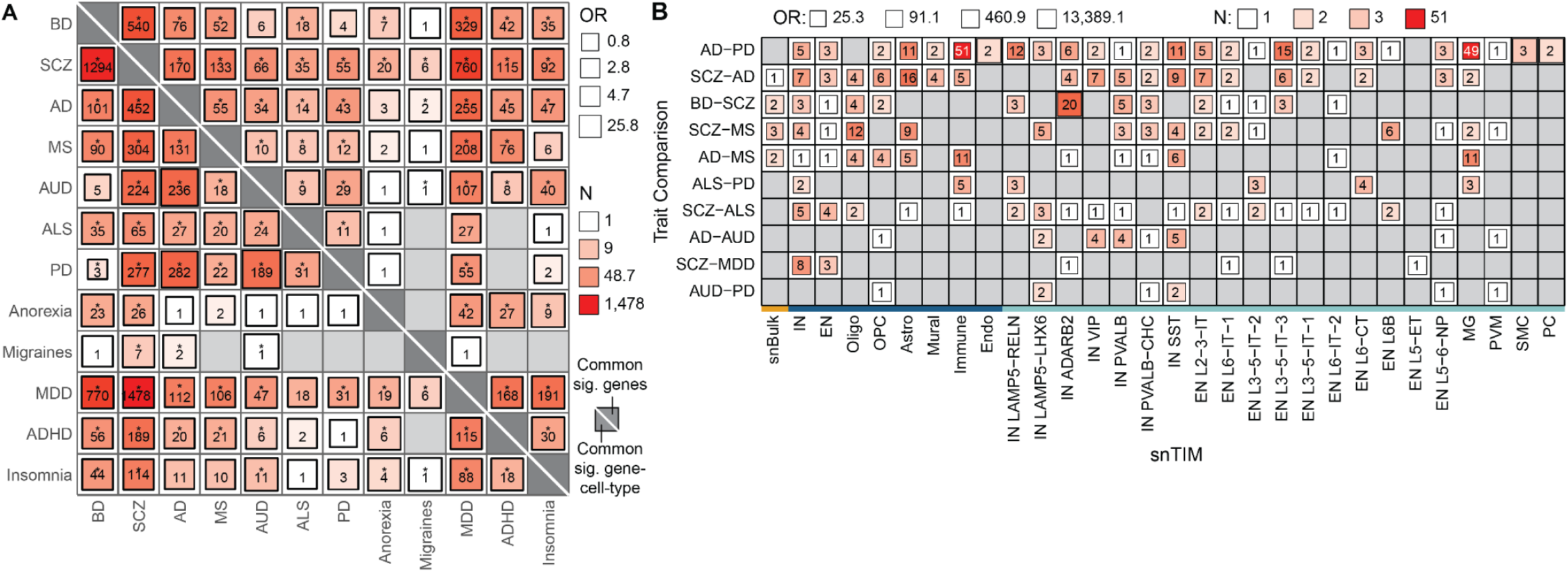
snTWAS uncovers shared genetic effects on gene expression dysregulation across disorders. **A,** Heatmap of cross-disorder significant association sharing for genes. The number and color in each cell describe the number of shared significant gene-cell-type combinations (*lower-left triangle*) or genes regardless of cell-type (*upper-right triangle*) between each pair of disorders. Size of the square indicates Fisher’s exact test odds ratio. Fisher’s exact test p-values were FDR^33^-corrected, and significant values are annotated by asterisks. **B,** Cross-disorder pathway sharing. All trait pairs (y-axis) with shared pathways are visualized in this heatmap across all snTIMs (x-axis). The number and color in each cell indicate the number of pathways significant (FDR ≤ 0.05) in both traits after hierarchical pruning (see Methods), and the size of the square denotes the strength of enrichment for pathway sharing between the two traits within the cell-type (Fisher’s exact test).

Identifying cross-disorder shared biological pathways is greatly enhanced by performing cell-type-specific analyses (Fig. 4B; Table S36). For example, cross-disorder comparisons of AD with PD and multiple sclerosis (**MS**), show the highest number of shared pathways in Class-Immune and Subclass-MG (Fig. 4B). Other examples where shared biology is better uncovered at single cell resolution are the bipolar disorder (**BD**)-SCZ and SCZ-MDD comparisons, where Subclass-IN ADARB2 and Class-IN uncover the highest number of shared pathways, respectively (Fig. 4B; Table S36). Thus, increased cellular resolution aids in the identification of shared biological pathways across NPDs and NDDs that cannot be detected in snBulk (Fig. 4B) despite evidence for high global (e.g. for SCZ-BD-MDD^51^) or local (e.g. for AD-PD^52^) genetic correlation.

### Ancestry-specific snTWAS uncovers shared biology and aids in fine-mapping of risk genes

Previous work highlighted the poor portability of TIMs across ancestries as a main limitation of multi-ancestry TWAS studies^53,54^. Towards a large-scale cross-ancestry exploration of GReX dysregulation in NPDs and NDDs, we bypassed such limitations by performing I-snTWAS in MVP with matched ancestry individuals and snTIMs for 9 NPDs and NDDs (reduced number of traits compared to S-snTWAS due to sample size considerations). Importantly, our PTCA analysis showed strong correlation among top ranked GTAs across ancestries that was comparable to the within-ancestry comparison of S-snTWAS and I-snTWAS in EUR (Fig. 5A; Table S17). Similarly, we showed high concordance between EUR and AFR I-snTWAS-derived pathway analysis (Fig. 5B). In spite of the lower effective sample size in MVP compared to GWAS studies (Table S17), we were able to find significant GTAs across ancestries (Supplementary Fig. 12; Data S3). However, we opted to limit our in-depth I-snTWAS analysis to EUR and AFR since the MVP had a comparatively small (n = 61,073; 9.28% of total) and heterogeneous^55^ AMR subpopulation which, despite showing good concordance for targeted replication^13^, was less suitable for large-scale GReX scans.

**Fig. 5.**
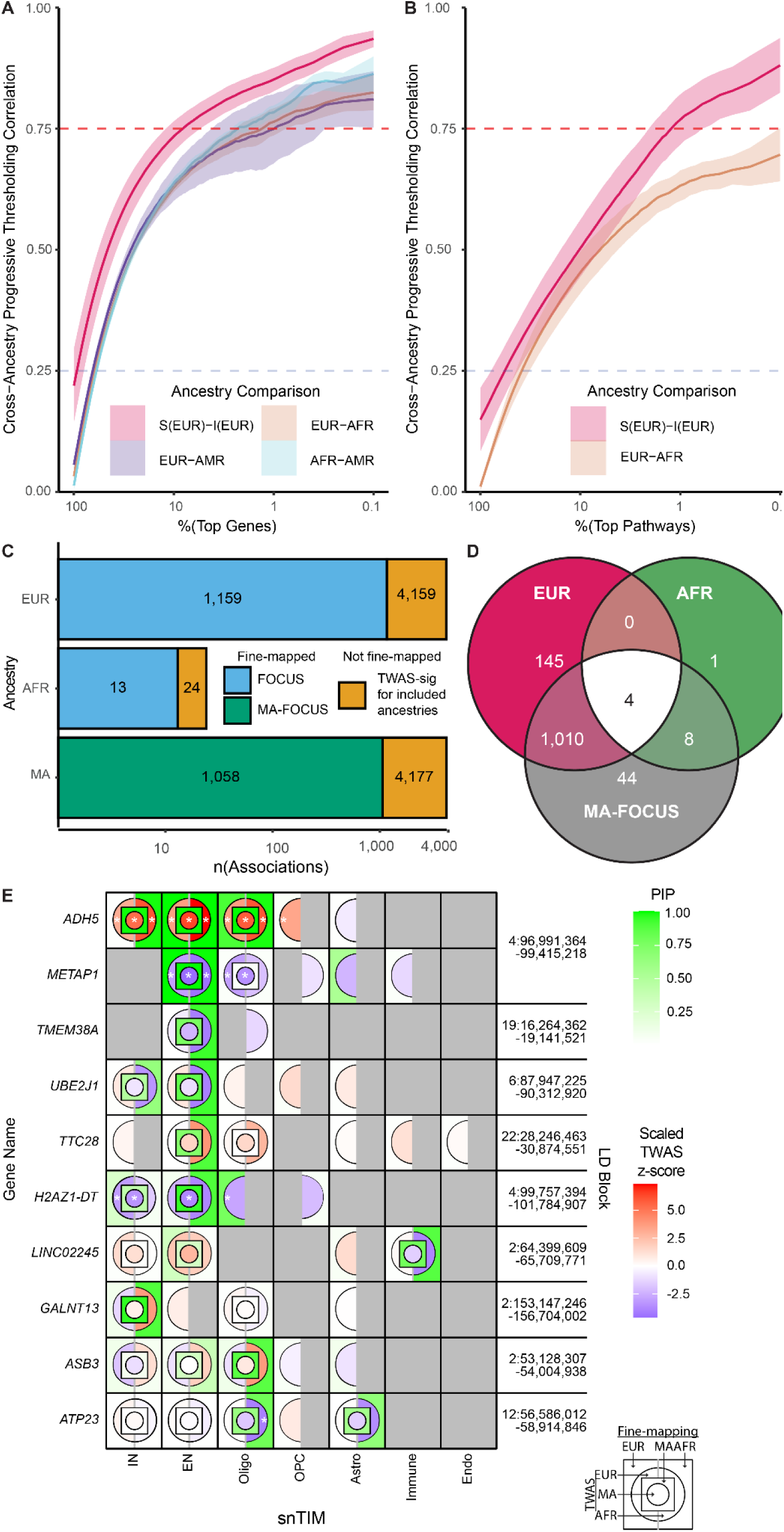
Cross-Ancestry snTWAS identifies similar GTAs across ancestries. **A,** Cross-Ancestry PTCA. Line represents mean correlation between the two comparison groups among the 9 traits considered in I-snTWAS analysis. Shaded area represents the 95% confidence interval. Red and blue dashed horizontal lines denote a 0.75 and 0.25 Pearson’s correlation coefficient (r), respectively. “S(EUR)-I(EUR)” corresponds to the comparison of EUR S-snTWAS against EUR I-snTWAS. EUR, AFR and AMR correspond to their respective I-snTWAS. **B,** Cross-ancestry I-snTWAS pathway-level progressive thresholding correlation analysis (PTCA). Each line graph tracks the cross-disorder association z-score Pearson’s correlation for progressively higher ranked pathway-cell-type combinations. Red and blue dashed horizontal lines denote a 0.75 and 0.25 Pearson’s correlation coefficient (r), respectively. **C,** Overlap of TWAS and fine-mapping in single ancestry and multi-ancestry fine-mapping across the 9 I-snTWAS traits. Annotated numbers in the barplot indicate the total number of associations in each category (i.e. there are 1,159 fine-mapped trait-gene-cell-type combinations in EUR and 4,159 I-snTWAS significant trait-gene-cell-type-combinations). Y-axis indicates the population in which analysis was performed. “MA” indicates the MA-FOCUS bi-ancestral analysis and the union of EUR and AFR I-snTWAS significant trait-gene-cell-type combinations for fine-mapping and TWAS, respectively. **D,** Overlap of fine-mapped trait-gene-cell-type combinations (FOCUS for EUR and AFR; MA-FOCUS for bi-ancestral EUR and AFR). **E,** Bi-ancestral (EUR and AFR) fine-mapping of AUD. The top 10 AUD-associated genes in the AFR I-snTWAS analysis are visualized across class level snTIMs. Z-scores are scaled across all genes within each population.

The high concordance visualized in PTCA analysis (Fig. 5A) highlighted the broad conservation of GReX dysregulation across NPDs and NDDs; however, only 25% (5/20) of the significant GTAs identified in the AFR I-snTWAS were also significant in the EUR I-snTWAS (even after considering all cell-types). Given that replication power was limited for many of our traits in I-snTWAS, we formally assessed the potential benefits of a multi-ancestral approach by comparing ancestry-specific against bi-ancestral TWAS fine-mapping. Despite a similar proportion of significant I-snTWAS associations, bi-ancestral fine-mapping captured a greater proportion of AFR fine-mapped associations (Fig. 5C and Fig. 5D). Furthermore, we showed that these proportions were consistent when looking at GTAs rather than trait-cell-type-gene combinations (Supplementary Fig. 13A and Supplementary Fig. 13B), and that the distribution of bi-ancestral fine-mapping PIPs resembled both EUR and AFR within their respective significant associations (Supplementary Fig. 13C). We then explored potential benefits of utilizing bi-ancestral (EUR and AFR; MA-FOCUS) gene prioritization for LD blocks with multiple GTAs in alcohol use disorder (**AUD**) - the phenotype with the highest number of GTAs in our I-snTWAS (Supplementary Fig. 14). Notably, one of our top fine-mapped associations in AFR, *ATP23*, resided in an LD block with *ARHGEF25*. *ARHGEF25* was the only EUR AUD GTA in this LD block, but utilizing MA-FOCUS shows that *ATP23* (significant in AFR only) was fine-mapped across ancestries (Fig. 5E; Supplementary Fig. 14; Table S37). Taken together, multi-ancestry snTWAS fine-mapping aided in both cell-type specific gene discovery and gene target prioritization.

### NPD/NDD-associated GReX has pleiotropic effects

To explore the pleiotropic effects of cell-type-specific GReX, we performed GReX-PheWAS in MVP on the top 1,101 significant snTWAS genes with all snTIMs (**snGReX-PheWAS**; Data S4). Towards validating our approach, we observed strong phenome-wide conservation of GReX dysregulation across all ancestries as evidenced by the PTCA (Fig. 6A) and association effect size sign concordance (Supplementary Fig. 15) analyses; a finding that replicates and extends our targeted I-snTWAS cross-ancestry analysis. We then limited the scope of our phenome scan to neurological, mental and behavioral, and sensory organ disorders (“focused”) within EUR ancestry to explore pleiotropic effects.

**Fig. 6.**
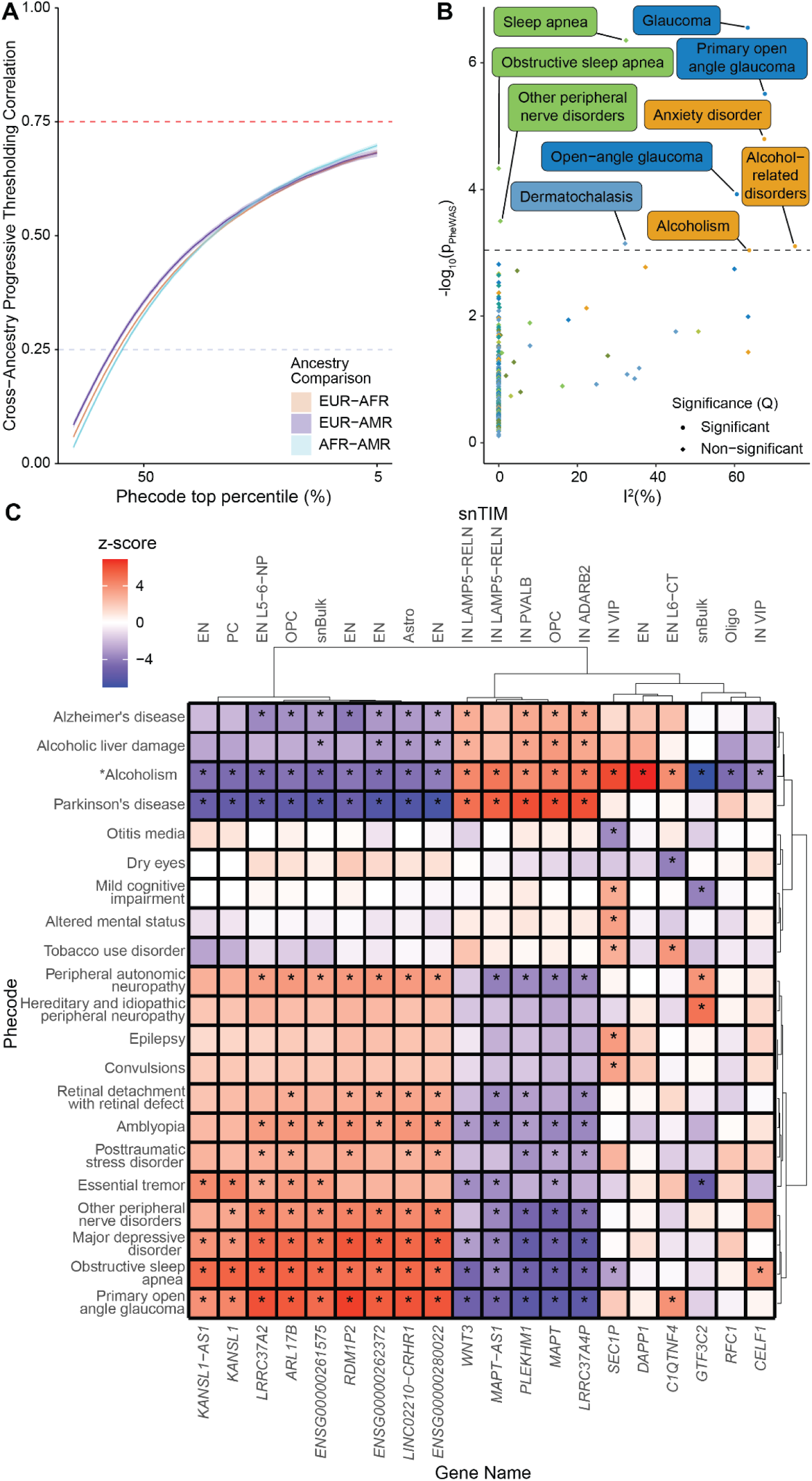
snGReX-PheWAS. **A,** Conservation of cross-ancestry correlation of GReX-PheWAS associations among top trait-gene-cell-type combinations. PheWAS was performed for all genes found to have a significant (Bonferroni-adjusted p-value ≤ 0.05) association in the I-snTWAS analysis spanning all snTIMs and ancestries (EUR, AFR and AMR). For each pairwise ancestry combination, phecodes were ranked by inverse variance weighted meta-analysis utilizing z-scores normalized in each ancestry to adjust for power differences across ancestries; only phecodes with at least 500 cases and 500 controls were considered. The resulting snGReX-PheWAS was then leveraged for three PTCA analyses corresponding to each ancestry pairwise combination among their shared gene-cell-type combinations (n = 1,757, 880 and 681 in EUR-AFR, EUR-AMR, and AFR-AMR, respectively). Consequently, the pairwise Pearson’s correlation coefficient is visualized among decreasing numbers of top phecodes (PTCA). Shaded region around each line represents the 95% confidence interval. Red dashed horizontal line denotes a Pearson’s r of 0.75 and 0.25, respectively. **B,** *CELF1* cell-type-specific effect size heterogeneity across phecodes. Each phecode is represented by the cell-type with the lowest association p value indicated by its correspondent color (<u>Extended</u> <u>Data</u> Fig. 2). To determine significance of effect size heterogeneity (Cochran’s Q), we only consider cell-types with significant associations (FDR^33^ < 0.05). Horizontal dashed line corresponds to the -log_10_(p) value of the weakest significant (FDR< 0.05) association. **C,** Clustering of top relevant PheWAS associations in AUD. PheWAS results are visualized for the top 20 gene-cell-type combinations identified in the AUD S-snTWAS; each gene is represented by the cell-type with the lowest association p value. In addition to replicating the association with AUD (mapped to the “alcoholism” phecode), the top 20 phecodes among relevant phecode categories (including neurological, mental health and sensory organ disorders) ranked by association p value are visualized. Ward’s hierarchical agglomerative clustering was performed with Ward’s criterion preservation.

In the I-snTWAS section (Fig. 3C), we demonstrated for a given trait (AD) the variability across genes of effect size heterogeneity among cell-types (I^2^_cell-type_); here, we present a multi-cell-type GReX-PheWAS to illustrate how the I^2^_cell-type_ for a given gene (*CELF1*) can vary across phenotypes (Fig. 6B; Table S38 for all significant associations within genes selected for PheWAS). *CELF1*, which has no significant GTAs in snBulk, has a wide range of I^2^_cell-type_ values across the relevant phenome where the GTAs are driven by different cellular populations (Fig. 6B; sn-eQTL support in Supplementary Fig. 16). For a gene like *CELF1*, the observed complexity and diversity in snGReX-PheWAS associations underscores our current underappreciation of the pleiotropic effects of genetic variants on gene expression dysregulation across traits in a cell-type-specific manner. Overall, we identify significant heterogeneity among cell-types in PheWAS associations in 57% of genes (579/1,012 EUR PheWAS genes imputable in more than 1 cell-type). Thus, performing snGReX-PheWAS can help us understand the pleiotropic effects of trait-associated GReX dysregulation with cell-type-specificity. For example, performing a focused EUR snGReX-PheWAS for the top 20 AUD GTAs (from the S-snTWAS analysis) (Fig. 6C; Supplementary Fig. 17) validates the associations with “Alcoholism” and uncovers relevant pleiotropy consistent with our shared pathway analysis. Specifically, we identify strong correlation with AD (r = 0.88, p = 4.44 × 10^-7^) and PD (r = 0.75, p = 1.24 × 10^-4^) that span most of the top genes and their respective cell-types. Regarding associations with established AUD comorbidities and complications, we observe strong positive correlation with alcoholic liver damage (r = 0.91, p = 1.72 × 10^-8^), negative correlation with disorders where alcohol is reported for self-medicating, including essential tremor (r = -0.50, p = 2.34 × 10^-2^), post-traumatic stress disorder (**PTSD**) (r = -0.67, p = 1.30 × 10^-3^) and major depressive disorder (r = -0.74, p = 1.65 × 10^-4^) (Fig. 6C).

## Discussion

Prior to the widespread adoption of snRNA-seq approaches, several low-throughput studies investigated genetically regulated gene expression in specific cellular populations of the human brain^7,15–17^. Recently, snRNA-seq^24,56^ has enabled a more systematic characterization of cell-type specific gene expression in the brain; however these studies have been somewhat limited in scale for EUR (192^56^ and 424^24^ donors) and did not model GReX in other ancestries. Here, we present a large-scale brain TWAS that leveraged snTIMs for EUR (n = 920), AFR (n = 321) and AMR (n = 118) ancestries in 12 NPDs and NDDs as well as across the phenome in the MVP cohort (n_EUR_=1,428 phecodes; n_AFR_=1,057; n_AMR_=725)

Across all snTIMs, we were able to both greatly expand the GReX repertoire by imputing 11,266 genes not captured in a similarly sized homogenate TIM (Bulk) from the same region (DLPFC)^13^, and outperform the Bulk TIM in imputation accuracy. Next, in our S-snTWAS for 12 NPDs and NDDs, we established that higher snTIM cellular resolution leads to the discovery of more GTAs with a higher likelihood of them being novel and clinically relevant. Compared to the only published snTWAS in MDD^24^, we noted a 24% power increase among commonly identified gene-cell-type combinations. Our thorough investigation into cell-type-specificity with both probabilistic (based on mash in S-snTWAS) and frequentist (I^2^ in MVP I-snTWAS to overcome limitations in accurate effect size estimation in S-PrediXcan^45^ and TWAS/FUSION^37^) approaches revealed that the most unique signal among brain cell-types in NPDs and NDDs stems from the Class-immune, which is unsurprising given their distinct erythromyeloid origin^57^. In addition, disease-relevant pathways were preferentially, and in some cases uniquely, captured in biologically relevant cell-types.

Beyond single-trait analyses, snTWAS also aids in uncovering cell-type-specific cross-disorder biology. Specifically, among 12 NPDs and NDDs, we observed increased likelihood of shared GTAs and pathway dysregulation in 68% and 21% of trait pairwise comparisons, respectively, and uncovered biology that would otherwise go undetected in homogenate TWAS. For example, despite identifying no common pathways in snBulk TWAS, AD and PD shared 49 pathways in microglia; a finding supported by local genetic correlation analysis^52^ and their common heritability enrichment for microglia^7,58^. Finally, in our snGReX-PheWAS, we were able to validate the S-snTWAS findings, explore the variability of GReX cell-type heterogeneity across traits, and uncover cell-type specific associations with genetically correlated traits, comorbid conditions and trait-associated complications. For cross ancestry analyses, snTIM performance metrics for EUR, AFR and AMR expectedly reflected their respective training sample sizes. Despite their lower power, AFR and AMR snTIMs captured an additional 2,379 and 1,558 genes that cannot be reliably imputed by EUR snTIMs. Interestingly, on average, the level at which a gene’s expression is under genetic regulation is consistent across ancestries and cell-types. Similarly, between ancestries, we report strong correlation and concordance among the top ranked GTAs for both snTWAS and GReX-PheWAS. In addition, non-EUR snTWAS uncover ancestry-specific GTAs and aid in the multi-ancestry fine-mapping of GTAs. Most of the literature focuses on the poor portability of EUR TIMs for imputing GReX for other ancestries^53,54^. However, here we demonstrate that ancestry-matched experimental designs are very effective for cross-ancestry NPD/NDD studies even with limited sample sizes, paralleling prior diverse ancestry blood monocyte TWASs^59–61^.

Overall, our study builds on existing literature to highlight the importance of cell-type specificity for identifying genetically driven actionable gene expression changes for translational applications. Compared with bulk approaches, there seems to be no downside at the gene level in transitioning to snRNA-seq beyond cost and instances where capturing extranuclear RNA might prove desirable. However, for optimal model performance, in addition to the number of donors, future experimental designs should also consider optimizing the number of nuclei captured per specimen. In addition, standard snRNA-seq approaches do not estimate isoform abundance which can capture more heritability than genes^62^. Inclusion of isoform information would further improve resolution and, in so doing, increase our understanding of the cell-type specific nature of disease.

In conclusion, our study demonstrates that incorporating cell-type resolution and ancestry specificity into TWAS frameworks substantially enhances gene discovery and provides deeper insights into the biological underpinnings of neuropsychiatric and neurodegenerative disorders. By moving beyond bulk tissue analyses, we pave the way for more targeted therapeutic interventions that consider the intricate cellular landscape of the human brain.

## Methods

### Training of snTIMs

#### PsychAD Cohort

Molecular profiling efforts of the PsychAD consortium include snRNA-seq from more than 6 million nuclei from the DLPFC of 1,494 unique donors^25^. As previously described in the Capstone paper^22^, we utilize Quadratic Discriminant Analysis (**QDA**) using the 1000 Genomes Project^63,64^ reference to divide the full cohort into five discrete superpopulations based on ancestry: European (**EUR**), African (**AFR**), Ad-Mixed American (**AMR**), East Asian (**EAS**), and South Asian (**SAS**). QDA identifies 1,359 genotyped individuals (920 EUR, 321 AFR and 118 AMR)^22^ who underwent snRNA-seq-based gene expression profiling; data from the remaining individuals (EAS and SAS) were not considered for TIM building in our analyses due to lack of sufficient power. Furthermore, PsychAD is composed of 3 “subcohorts” (MSSM, HBCC, RADC), of which RADC is predominantly AFR (Table S39). We utilized snRNA-seq data from all eight cell-type classes, and 23 subclasses (excluding 4 subclasses that are identical to classes, and Subclass-EN NF, which had too few cells and individuals for snTIM training). Finally, we note that we utilized release 2.5 of PsychAD.

#### Genotype and gene expression quality control for TIM training

##### Generation of an ancestry-specific common variant reference list

The merged PsychAD genotype dataset consisting of PsychAD-MSSM SNP array, CommonMind SNP array, Rush Alzheimer’s Disease Center (RADC) WGS, ADSP WGS samples was prepared as described previously^25^. Towards harmonizing SNP utilization across the training (merged PsychAD genotype dataset) and target cohorts (GWAS for S-TWAS and TOPMED-imputed SNP arrays for MVP), we constructed ancestry-specific common variant reference panels. First, we queried all variants in the TOPMED-imputed^65–68^ Million Veteran Program’s (**MVP**)^65^ genotype release 4 and retained non-ambiguous biallelic SNPs with rsID annotation included in MVP individuals that passed the following filters (see below for variant-level and sample-level quality control (**QC**)): population-specific minor allele frequency (**MAF**) greater than or equal to 0.01, population-specific genotype missingness less than or equal to 0.05, and TOPMED imputation R^2^ greater than or equal to 0.6. The resulting ancestry-specific SNPs were retained if they also had a matched superpopulation-specific MAF greater than or equal to 0.01 in 1000 Genomes^64,63^. The resulting ancestry-specific common variant reference list comprises approximately 4.5, 7.5 and 5.1 million SNPs in EUR, AFR and AMR, respectively.

##### Variant level filtering in the training cohort (PsychAD)

Variant-level filtering on the genotypes used for the TIM training requires that all the following variant inclusion criteria were met: SNPs exist in the ancestry-specific SNP list described above, MAF ≥ 0.01, based on ancestry-specific information from NCBI Allele Frequency Aggregator (**ALFA**)^69^ (EUR: SAMN10492695; AFR: SAMN10492698; AMR: SAMN10492700), or an in-cohort MAF ≥ 0.01 when ALFA information was not available or the variant does not pass the ALFA filter, in-sample minor allele count (**MAC**) of 5 or greater, and a Hardy-Weinberg equilibrium p-value of 10^-6^ or greater, and do not fall within areas of high linkage disequilibrium (**LD**), such as the Major Histocompatibility Complex (Table S40). Finally, missing information for SNP predictors in existing TIMs were replaced with double the in-sample MAF (representing the in-sample average genotype), rounded to the nearest integer.

##### Gene expression QC

First, we limited genes to those within our expression annotation, Ensembl 104^70,71^. Second, for each “subcohort” within psychAD (see “PsychAD Cohort”), we independently scaled gene expression, performed PEER^72^ to identify and adjust for hidden factors driving gene expression differences, and quantile normalized the resulting residualized gene expression. We utilized fastQTL^73^ to determine the number of PEER factors (ranging from 5 to 50) that yield optimal genetic signal by identifying the point at which the number of significant eQTLs (FDR^33^-adjusted p-value ≤ 0.05) is closest to 95% of the maximum number of significant eQTLs found across any assessed number of PEER factors. Finally, we combined residualized gene expression across the 3 “subcohorts”, and repeated scaling, PEER factor optimization, and quantile normalization of the combined cohort to reduce the impact of the batch effect (Supplementary Fig. 18).

##### Training of cell-type specific PrediXcan models

We used PrediXcan^74^ to create per-cell-type TIMs in each of three populations (EUR, AFR, and AMR) using the QCed genotypes and gene expression. Imputable genes are considered passing cross-validation R^2^ (R^2^_CV_) ≥ 0.01, p_CV_ ≤ 0.05 and SNPs in model > 0. R^2^_CV_ and p_CV_ values are prediction performance R^2^ and prediction performance p-value from the “PredictDB” software^75^. To compare gene imputation models across individual TIMs or ancestries, we utilize R^2^_CV_, a proxy of variance in gene expression explained by genetic variants serving as predictors.

We performed linear regression to assess the extent to which major snTIM metadata predictors (sample size and median number of nuclei contributing to the pseudobulk expression from every individual) influence snTIM performance (proxied by number of confidently imputed genes). We report the adjusted R^2^ as a measure of the variation explained in snTIM performance for each predictor; adjusted R^2^ was estimated by the stats package^76,77^.

We compared snTIMs against a DLPFC^26^ EpiXcan^9^ TIM^27^ (referred to as “Bulk”). We compared whether genes were more strongly imputed in psychAD snTIMs or Bulk using a sign test. To more accurately calculate the sign test p-value, we utilized Ramanujan’s factorial approximation^78^.

### Abundance analysis of imputable genes

We hypothesized that genes uniquely identified by snBulk versus other snTIMs were lower in abundance. To investigate this, we subsetted imputable genes into those uniquely identified by snBulk (904) and those identified by at least one other snTIM (19,285). We calculated the mean number of transcripts for every gene amongst all EUR individuals with available expression data for the gene. To establish the statistical significance of the difference in mean transcript count, we used the Kruskal-Wallis test.

### Summary-level TWAS using summary statistics

We selected 12 well-powered NPD/NDDs GWAS summary statistics^79–90^ (Table S17) for our summary-level TWAS (**S-TWAS**) analysis. We utilized MungeSumstats^91^ to standardize GWAS format and update rsID annotations, and, where possible, we use 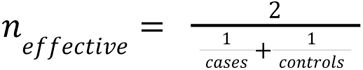 as sample size. To ensure maximum utilization of TIM SNPs, we imputed missing variants in GWAS summary statistics based on established methods^92^ (Supplementary Fig. 19). Ancestry-specific imputation LD panels were built on the 1000 Genomes Project with plink^93^ and the following parameters: --ld-window-kb 1000000 --ld-window 1000 --maf 0.01 --ld-window-r2 0. We employed run_imputez^92,94^ using a maxWindowSize of 200. To validate imputed GWAS summary statistics, we randomly sampled 1,000 SNPs per chromosome common to the GWAS and LD reference panel, and correlated real and imputed z-scores (Supplementary Fig. 19). Only SNPs with a GWAS imputation r^2^ ≥ 0.7 were retained. Finally, we performed summary-level TWAS (**S-TWAS**) for our TIMs (<u>Data S1</u>) using S-PrediXcan^45^. Significant gene-cell-type combinations were determined using the FDR^33^-adjusted p-value cutoff of 0.05. FDR was calculated across all gene-cell-type combinations for each trait.

We validated S-snTWAS associations using two external data sets, FACS-MG and Zeng-2024^24^. We utilized the average increase of the *χ*^2^ statistic, as described previously^9^, to determine the increase in power by using psychAD snTIMs over the data described in Zeng-2024.

### Individual-level TWAS and GReX-PheWAS in MVP

#### Million Veteran Program Genotype Quality Control

The MVP’s data core team handles genotyping, SNP imputation, and initial quality control of genotypes. For this study, we utilized the MVP Release 4, which includes genotypes from 662,681 individuals. DNA was extracted from whole blood and genotyping was performed with the MVP 1.0 custom Axiom Array^65^. The MVP data core called genotypes using APT version 2.11.3 and MVP 1.0 array library r6. Genotyping was validated using three plates of 1000 Genome samples. Rigorous genotype quality control such as plate normalization was used to improve the accuracy of genotype calling. Genotype imputation was performed using the TOPMED^66^ reference and ancestral principal components were generated using EIGENSOFT v.6^95,96^. We then performed sample-and variant-level QC on genotypes. Our QC pipeline is based on previously established methods for other genetic analyses and optimized for maximizing input information for GReX estimation^97^. We grouped samples by ancestry based on HARE^98^ (a method that integrates and harmonizes self-identified ancestry and genetic ancestry) into three ancestries (438,582 EUR; 112,346 AFR; 48,726 AMR). Using only autosomal variants, we filtered variants for MAF (≥ 0.01), Hardy-Weinberg equilibrium p-value (1×10^-6^), and removed high disequilibrium regions. We then filtered for excess heterozygosity (≤ 4 standard deviations from the mean), ambiguous sex (using both plink’s check-sex option, as well as sex chromosome aneuploidies assignments derived from plate intensity measurements), and relatedness (KING filter = 0.0884)^93,99^. Finally, we limited variants to biallelic SNPs for individual imputation of GReX.

#### MVP phenotypes

Phenotypes from ICD-9/10 codes were transformed into phecodes using Phecode Map v.1.2^100,101^. Moreover, individuals with at least 2 phecodes for a given trait were considered a case, whereas individuals with fewer than 2 phecodes were considered a control for both I-snTWAS and GReX-PheWAS analyses. For the I-snTWAS analysis only traits with at least 2% case prevalence in EUR, AFR and AMR were utilized for power considerations (Table S17), and AD was mapped to the phecode for “Delirium dementia and amnestic disorders”, 290, due to the way AD is routinely coded in the VA medical system^102^. For the GReX-PheWAS analysis, traits with at least 500 cases and 500 controls were considered.

#### GReX imputation

We estimated GReX in the MVP on an individual-level using TIM-derived SNP predictor weights. Missing genotypes were replaced with double the in-sample MAF (representing the in-sample average genotype), rounded to the nearest integer.

#### I-snTWAS association analysis

Logistic regression analysis was performed while adjusting for sex, age, and top ten ancestry principal components. Due to the comparatively lower power of the MVP (vs. GWAS), multiple test correction with FDR^33^ (≤ 0.05) was performed within each snTIM rather than across relevant snTIMs. For comparison of I-snTWAS with S-snTWAS we also utilized GWAS summary statistics for Anxiety^103^ and PTSD^88^ (Table S17; Fig. 4C).

#### Gene selection for GReX-PheWAS

Due to computational resource limitations, PheWAS analysis was performed on top ranked genes from the S-snTWAS and I-snTWAS. Genes were selected via bonferroni-corrected p-value (≤ 0.05; bonferroni-correction applied across all cell-types for S-snTWAS analysis and within each cell-type for I-snTWAS analysis, for power considerations). S-snTWAS genes were considered across all cell-types in EUR. I-snTWAS genes were considered across all cell-types and ancestries. This resulted in a total of 13,456 PheWASs(approximately 5.39% of all imputable gene-cell-type combinations; Table S8) across 1,101 unique genes.

#### GReX-PheWAS association analysis

As in I-snTWAS, logistic regression analysis was performed while adjusting for sex, age, and top ten ancestral principal components. Associations with FDR^33^-corrected two-sided p-value ≤ 0.05 were considered significant.

### Enrichment for clinically relevant genes

To validate GTAs of the S-TWAS we performed gene set enrichment analysis (**GSEA**) for genes with entries in the “neurologicCentralNervousSystem” and “neurologicBehavioralPsychiatricManifestations” columns of the Clinical Synopsis tables of the Online Mendelian Inheritance in Man (**OMIM**) database^38^. For our analysis, we only considered gene-specific OMIM entries and not entries corresponding to large loci (e.g. regions spanning multiple genes), resulting in a final set of 1,949 genes. Query TWAS genes (nominal p-value ≤ 0.01; only protein-coding genes) were tested for enrichment against this gene set using a one-sided Fisher’s exact test followed by Benjamin-Hochberg FDR^33^ multiple testing correction.

### Enrichment of Novel Genes

We qualified a gene-trait association as “novel” if the association was not present in Bulk DLPFC S-TWAS (FDR^33^-adjusted p-value < 0.05; see above) or MAGMA analysis (FDR-adjusted p-value < 0.05) performed using the MAGMA^40^ SNP2GENE function on the FUMA web portal^39,40^. GWASs were prepared as above and then lifted over to GRCh37 using liftover within MungeSumstats^91,104^ as required by FUMA^91,104^. FUMA default parameters were used, except for the following: “genetype” was set to “all”; MHC region was custom defined to “28477797-33448354” (to match the definition used in other analyses, accounting for genome build differences); window was set to 50kb (up-and down-stream). The union (∪) of FDR-significant S-TWAS and MAGMA hits was used to define our “known” list of gene-trait associations. Significant GTAs from the S-snTWAS not within this list were defined as “novel”. To test whether novel gene-trait associations were enriched in snTIMs, we separated significant gene-trait associations into 2 categories: significant in snBulk, and significant only in snTWAS (either class or subclass level; FDR-significant across all cellular populations). We used Fisher’s exact test to obtain odds ratios and bonferroni-corrected p-values for the enrichment of novel gene-trait associations in cell-type specific TIMs.

### TWAS Pathway Enrichment Analysis

We used JEPEGMIX2-P^47^ to perform LD-aware competitive pathway enrichment analysis. For this analysis we used GRCh37-aligned GWAS summary statistics (prepared as above in “Enrichment of Novel Genes”), snTIMs converted into JEPEGMIX2-P compatible annotation files. Additionally, we incorporated biological pathways from Gene Ontology (Gene Ontology 2015), accessed through MSigDB 5.1. JEPEGMIX2-P is designed to perform pathway enrichment analysis while accounting for LD structures among genetic variants. For this study, the software was enhanced to conduct competitive analyses using the CAMERA^105^ gene set test procedure, allowing for a more refined understanding of gene-pathway associations; the publicly available binary executable file was updated to include this enhancement. The derived p-values were FDR^33^-corrected among all pathways across all population-specific TIMs, and an 0.05 FDR threshold was used to determine significance. To obtain more conservative estimates of the number of significant pathways per cell-type while maintaining specificity, we removed all significant pathways that were “parents” of other significant pathways (referred as pathway pruning in the main text).

### mash Analysis

We implemented multivariate adaptive shrinkage (**mash**^43^) using the mashr package in R. Due to differences in the imputability of genes across snTIMs at the subclass level, GReX matrices are sparse; thus, we limited analysis to class-level snTIMs. Moreover, we excluded EUR Classes “Endo” and “Mural”, which confidently impute a much smaller number of genes in comparison to other class-level snTIMs. Then, we performed mash on z-scores set as effect size and standard errors set to 1; we didn’t use the reported association effect sizes due to limitations in accurate effect size estimation in S-PrediXcan^45^ and other TWAS methods^45^. We filled missing values (where we do not have a model for a given gene in a given cell-type), with a z-score of 0 and an arbitrarily high standard error (1 × 10^10^). We ran mash using data-driven covariances as recommended by the mashr authors. The mash analysis was used to obtain the probabilities of an effect (GTA) existing in a given cell-type (e.g. Extended Data Fig. 5D) and to assess cell-type-specificity in S-snTWAS (Fig. 3A and Fig. 3B). For the latter, we first limited the analysis to all S-snTWAS-significant GTAs within the 6 cell-types utilized in the mash analysis above, and further restricted GTAs to the ones having at least 1 but less than 6 (all) cell-types with a significant association (local false sign rate ≥ 0.95; due to bayesian statistics, not all S-snTWAS significant GTAs are necessarily significant in mash results).

### GTA Heterogeneity Analysis

I-snTWAS and GReX-PheWAS heterogeneity statistics were calculated for gene-trait association effect sizes in R using the metafor package^106^. Heterogeneity^46^ I^2^ across cell-types (I^2^_cell-type_) was calculated for genes imputable in at least two cell-type TIMs.

### Targeted sn-eQTL Analysis

To validate heterogeneity found in the same gene in different cell-types, we calculated sn-eQTLs among the SNPs chosen in TIMs. We created a SNP superset of SNPs utilized in each gene-cell-type imputation model, converted genotypes to dosage format, and performed linear regression of SNP dosage against residualized gene expression (see “Gene expression QC”).

### TWAS fine-mapping

To address potential horizontal pleiotropic effects and account for LD among SNPs utilized in snTIMs, we utilized the fine-mapping of causal gene sets (**FOCUS**)^42^. FOCUS models the marginal TWAS z-scores as a multivariate Gaussian distribution given the estimated eQTL effect size and the SNP correlation and utilizes a Bayesian approach to calculate the marginal posterior inclusion probability (**PIP**) for each gene, indicating its likelihood of being causal in a specific TWAS risk region. Due to internal GWAS imputation in FOCUS, we utilized post-munging GWAS summary statistics from our pipeline prior to missing SNP imputation. PrediXcan-based snTIMs were converted to FOCUS-compatible format after retaining only imputable genes. FOCUS was applied both individually to each snTIM to assess effects within each cell-type, and jointly (multi-cell-type) for further prioritization. Unless otherwise specified, we considered TWAS-significant associations with PIP ≥ 0.5 to be fine-mapped.

We evaluated whether fine-mapped genes increase in class and subclass aggregates by assessing the total number of unique fine-mapped genes and loci in each snTIM versus the respective aggregate. Because our primary analyses utilize FOCUS applied to individual snTIMs, we repeated the analysis using multi-cell-type fine-mapping. We also applied the Kruskal-Wallis test to compare genes per loci in aggregates versus individual snTIMs.

To perform bi-ancestral fine-mapping for EUR and AFR I-snTWAS in MVP, we employed multi-ancestry FOCUS (**MA-FOCUS**^107^). We utilized TOPMED^66^-imputed genotypes in MVP so no additional imputation was needed for SNP predictors in the snTIMs, individual-level SNP missingness was handled as above. To visualize the top AFR fine-mapped associations and their concordance with EUR fine-mapped associations and MA-FOCUS, we plotted the scaled I-snTWAS z-scores within each cell-type and ancestry (for meta-analysis, we performed inverse variance weighted meta-analysis using I-snTWAS effect size and standard error and scaled the resulting z-score). We restricted the analysis to class-level snTIMs and prioritized the top ten associations by ranking each gene by the maximum scaled AFR I-snTWAS z-score × AFR fine-mapping PIP across all considered cell-types.

### Genetic Correlation Analysis

We performed bivariate heritability analysis using LD Score Regression^108,109^ to assess the genetic correlation between the aforementioned 12 NPD/NDDs GWAS summary statistics. The datasets were munged to match the HapMap3 SNP allelic information and the LD weights were pre-calculated on the 1000 Genomes European dataset^110^.

### snGReX-PheWAS Clustering

To explore the pleiotropic effects on multiple phenotypes of top snTWAS GTAs, we performed clustering of snGReX-PheWAS results. For every snTWAS, we extracted the top 20 significant genes in association with the target phecode (Table S17). We extracted only the top associated gene-cell-type combination for each gene to prevent clustering due to homogeneity among gene-cell-type combinations driven by the same gene. We then extracted the top 20 phecodes associated with these gene-cell-type combinations in either brain categories (mental disorder, neurological, sense organs), or all categories. Ward’s hierarchical agglomerative clustering^111^ was performed on the association z-scores with an implementation that preserves Ward’s criterion^112^ (ward.D2 method in R).

### Progressive Thresholding Correlation Analysis

To assess and visualize the concordance of z-scores between two datasets (e.g. TWAS or summary statistics from competitive pathway enrichment analysis) at increasing significance thresholds, we performed the following analysis which we term “progressive thresholding correlation analysis” (**PTCA**). First, we scaled z-scores among all values in each dataset (using R’s base scale function, to normalize the values with a mean of 0 and a standard deviation of 1). Next, we matched identifiers (e.g. trait-cell-type-gene or trait-cell-type-pathway) between the two datasets and restricted the final dataset to elements common to both datasets. Then, we performed a fixed-effect inverse variance weighted meta-analysis using the dataset’s z-scores (standard error is assumed to be 1), and sorted the table based on the meta-analysis p-value (increasing order). Finally, we measured the correlation between ordered and scaled z-scores in a step-wise restrictive manner (assuming step size = 10 and the dataset consists of 1,000 common elements, we assessed the correlation between the top 1,000 elements, followed by the top 990 elements, etc.).

## Data availability

All results are included either in the main text or provided in Supplementary Tables or Data.

## Code availability

This project utilized publicly available code as described in the Reporting Summary.

## Acknowledgements

This research is based on data from the Million Veteran Program, Office of Research and Development, Veterans Health Administration, and was supported by award I01BX004189. This publication does not represent the views of the Department of Veteran Affairs or the United States Government. We thank the participants of the Million Veteran Program, the scientists, clinicians and supportive staff involved in the construction of this biobank, and the scientific computing staff for the expertise that they provided. We thank the computational resources and staff expertise provided by the Scientific Computing at the Icahn School of Medicine at Mount Sinai. This study was also supported by the National Institutes of Health (NIH), Bethesda, MD under award numbers R01AG067025 (PR), R01AG082185 (PR), K08MH122911 (GV), R01AG078657 (GV), BX004189 (PR), R01AG065582 (PR), R01AG067025 (PR), R01MH125246 (PR) and T32MH087004 (KT). Human tissues were obtained from the NIH NeuroBioBank at the Mount Sinai Brain Bank (MSSM; supported by NIMH-75N95019C00049), the Rush Alzheimer’s Disease Center (RADC; funding: P30AG10161, P30AG72975, R01AG15819, R01AG17917, R01AG22018, U01AG46152, and U01AG61356), and NIMH-IRP Human Brain Collection Core (HBCC, project # ZIC MH002903). This work was supported in part through the computational and data resources and staff expertise provided by Scientific Computing and Data at the Icahn School of Medicine at Mount Sinai and supported by the Clinical and Translational Science Award (CTSA) grant UL1TR004419 from the National Center for Advancing Translational Sciences.

## Ethics declarations

**Competing interests.** The authors declare no competing interests.

## Author information

These authors contributed equally: Georgios Voloudakis, Panos Roussos

## Contributions

Conceptualization and study design: SV, GV, PR. Data contribution or analysis tools: CD, PFNU, DB, DM, RK, CC, BA, BZ, AH, CC, MA, ZS, SA, KT, TB, PA, DB, SM, VH, KG, JB, DL, JFF, GEH, GV, PR. SV, ZW, MA, FT, GV performed the analyses. SV, JFF, GV, PR wrote the manuscript with input from all authors.

---

**Extended Data Fig. 1.**
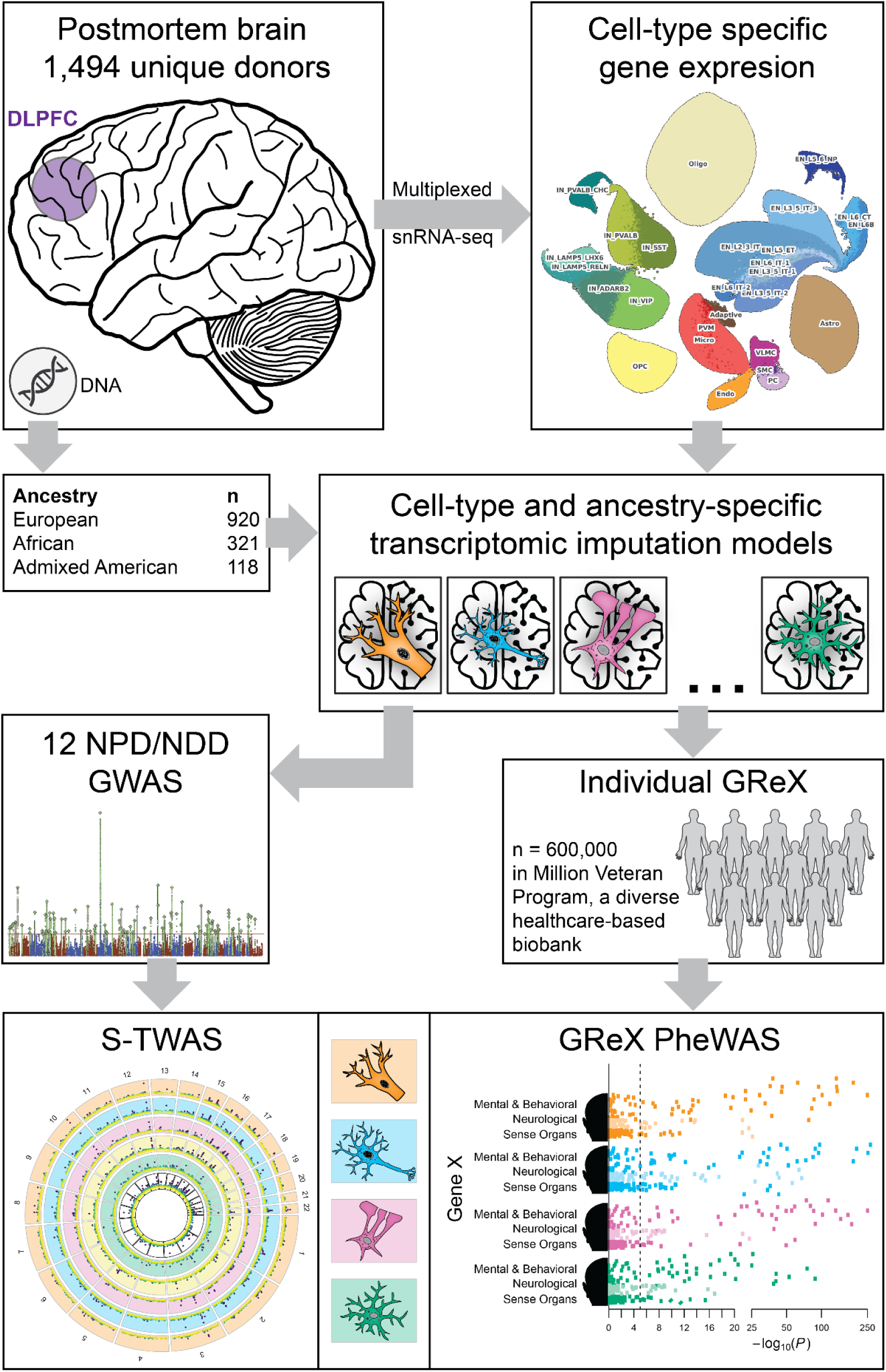
Graphical Abstract. A diagram depicting the overall study design.

**Extended Data Fig. 2.**
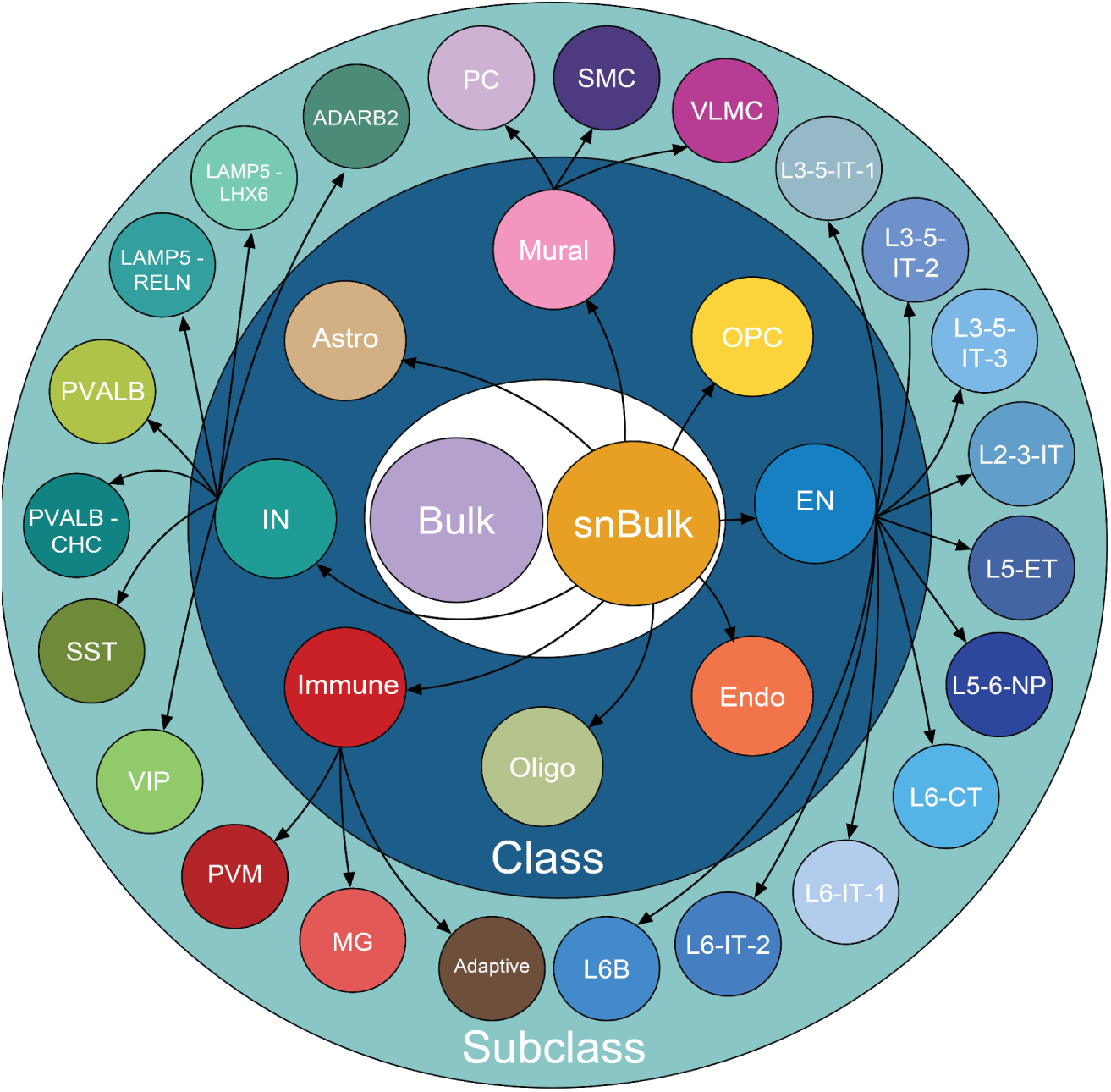
PsychAD Cell-type hierarchy chart. A hierarchy chart of the cell-types considered in this study. Cell-types are split into 3 levels depending on level of resolution. (*) Class-OPC, Astro, Endo, and Oligo are not further subdivided at the subclass level and are included both in the class and subclass aggregate analyses. Cell-type abbreviations are expanded in Table S1.

**Extended Data Fig. 3.**
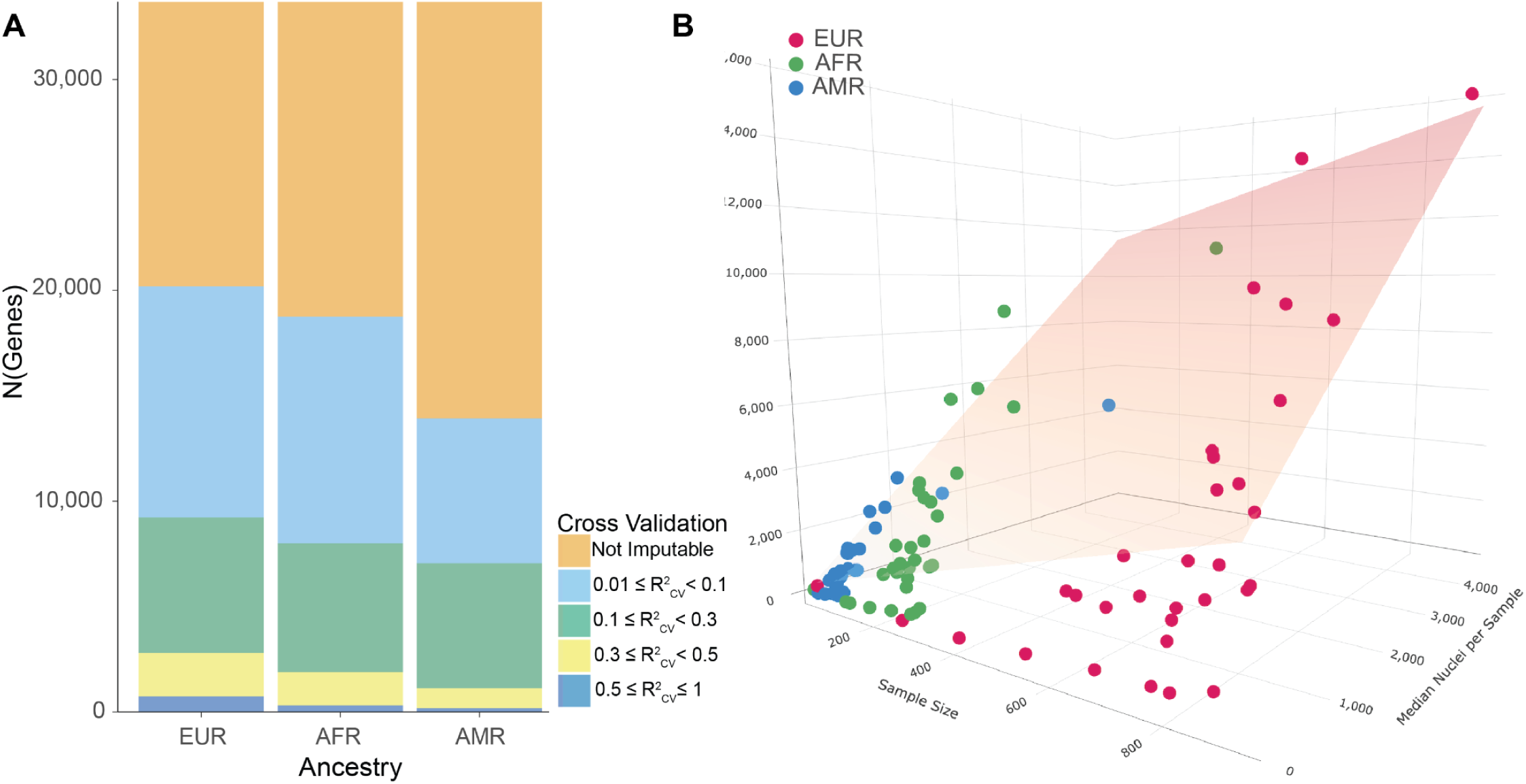
Multi-ancestry snTIM training. **A,** Gene imputability among snTIMs. The “Not Imputable” category comprises gene models with R^2^_CV_ < 0.01 or p_CV_ > 0.05. The remaining categories correspond to imputable genes with progressively greater gene expression variance explained (R^2^_CV_) by SNPs in the model. The model for each gene with the greatest R^2^_CV_ amongst all gene models (in snBulk, Class, and Subclass level snTIMs) is visualized. **B,** Major predictors of number of imputable genes for snTIMs. X-axis corresponds to the number of donors (“Sample Size”); Y-axis corresponds to the number of imputable genes; Z-axis is median number of nuclei contributing to the pseudobulk expression from every individual (“Median Nuclei per Sample”). The plane corresponds to the multiple linear regression model: N of Imputable Genes = 4.53 × “Sample Size” + 2.49 × “Median Nuclei per Sample” + 164.49; p = 2.63 × 10^-22^; adjusted R^2^ = 0.657; “Sample Size” and “Median Nuclei per Sample” predictors are significant (Table S9).

**Extended Data Fig. 4.**
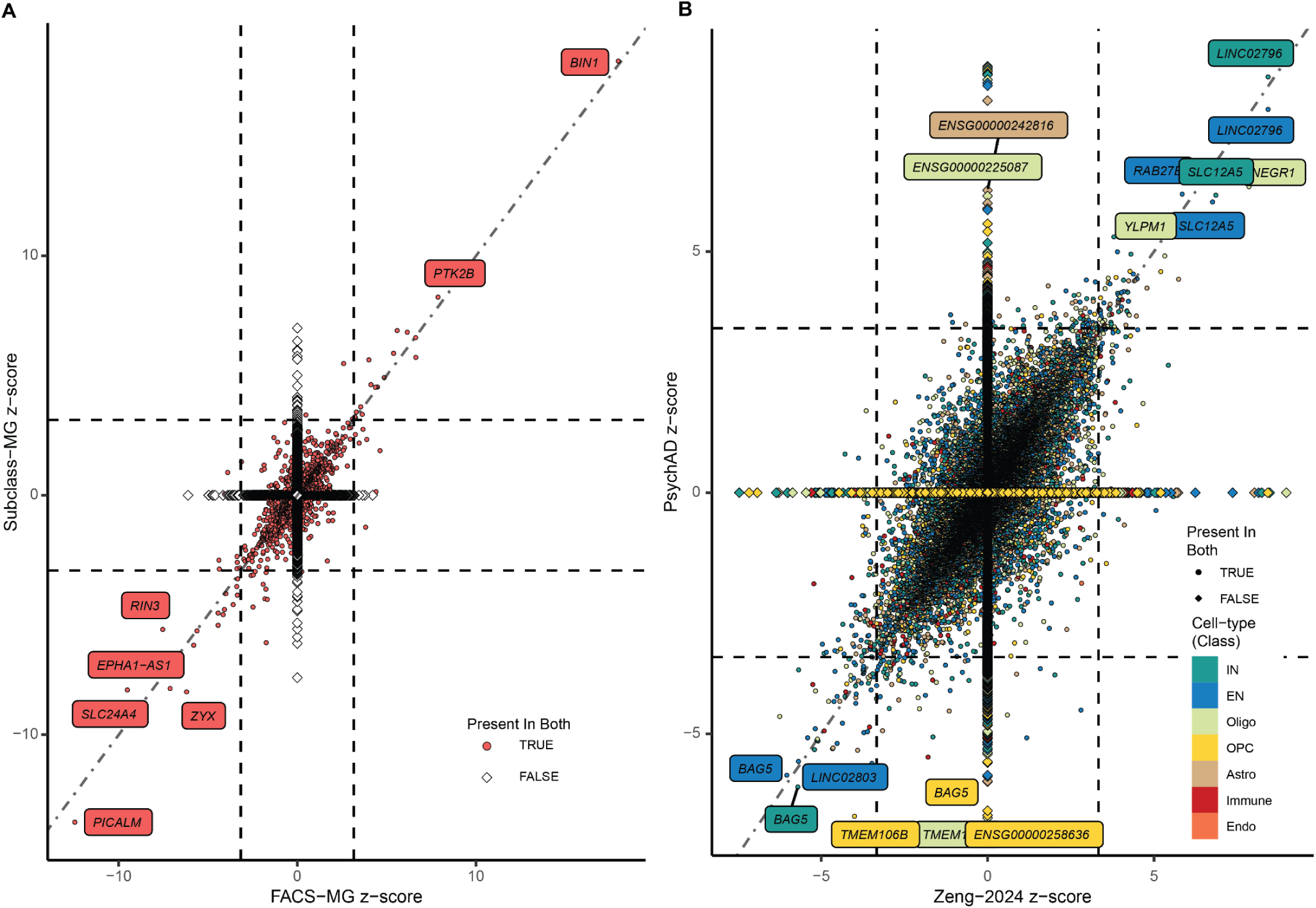
S-snTWAS validation. **A,** AD TWAS z-score comparison between FACS-MG and PsychAD Subclass-MG. Genes imputable in both TIMs are marked as points, and genes imputable in only one dataset are marked as diamonds. We observe a Pearson’s correlation coefficient of 0.84 (p = 2.25×10^-263^). **B,** Major depressive disorder (MDD) TWAS comparison between Zeng-2024 snTIMs and PsychAD snTIMs. Genes imputable in both TIMs are marked as points, and genes imputable in only one dataset are marked as diamonds. We observe a Pearson’s correlation coefficient of 0.757 (p=6.84×10^-3,217^) and a jaccard similarity index of 0.325.

**Extended Data Fig. 5.**
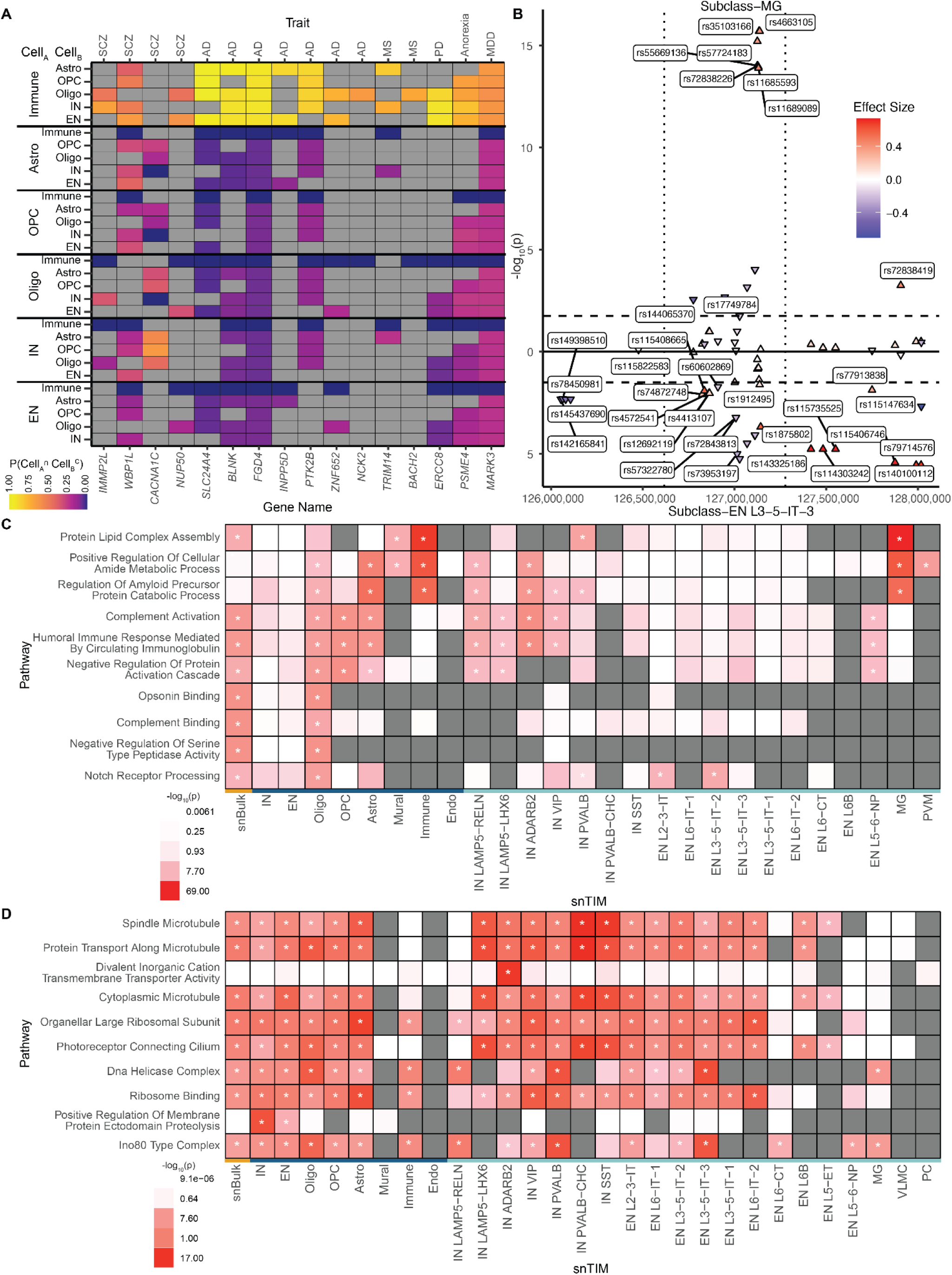
Cell-type specific genes, SNPs and pathways in snTWAS. **A,** A heatmap depicting the combinatorial probability that an effect exists in cell-type A and not in cell-type B at the class level. Only results with a 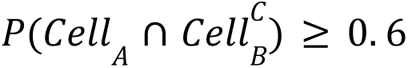 in at least one comparison were retained for visualization comprising 16 gene-trait combinations. Cell-type probabilities are estimated with mash. Cells are colored gray when the analysis is not applicable. **B,** sn-eQTLs for *BIN1* across 2 cell-types. Cell-types were chosen based on the differing association of cell-type specific *BIN1* GReX with AD. Labeled points indicate SNPs in which there is a significant (FDR^33^ ≤ 0.05; threshold for significance is indicated by horizontal dashed lines) effect in one cell-type but not the other. No SNPs contained significant effects in opposite directions for the two cell-types. Plotted SNPs were selected using a superset of SNP predictors in both *BIN1* snTIMs. Upward arrow indicates positive effect size and downward arrow indicates negative effect size. The vertical dotted line divides the LD blocks (utilizing EUR LD blocks as previously described)^113^. **C & D,** Top 10 pathways enriched in AD and SCZ. Enriched pathways for AD (**C**) and SCZ (**D**) were prioritized by camera p-value. For visualization purposes, hierarchical pruning was performed to retain distinct pathway signals (see Methods), and only up to 2 significant pathways were contributed from each cell-type to increase cell-type-specific representation.

