## Supplementary Information for "Single-nucleus transcriptome-wide association study of human brain disorders"

### SUPPLEMENTARY FIGURES

#### Supplementary Fig. 1

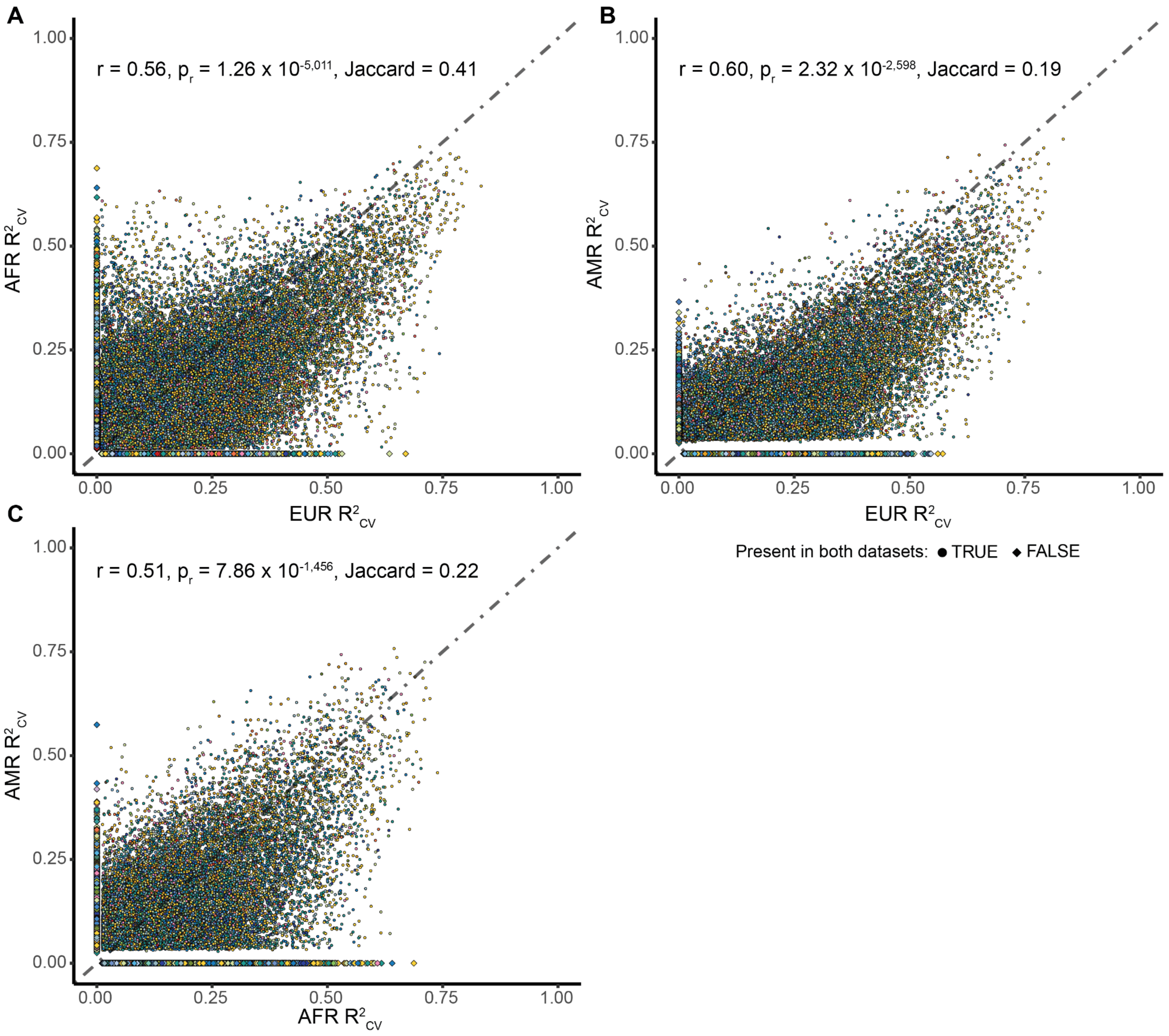

**Supplementary Fig. 1 | Comparison of gene-cell-type prediction performance across ancestry-specific snTIMs.** Color of each point represents the cell-type (see legend). Comparisons are for EUR vs AFR (A), EUR vs AMR (B) and AFR vs AMR (C) snTIMs. **A.** 3,819 genes are imputed in EUR and not AFR snTIMs, and vice versa for 2,378 genes. **B.** 7,824 genes are imputed in EUR and not AMR snTIMs, and vice versa for 1,558 genes. **C.** 6,795 genes are imputed in AFR and not AMR snTIMs, and vice versa for 1,970 genes. r: Spearman's correlation coefficient.

Supplementary Fig. 2

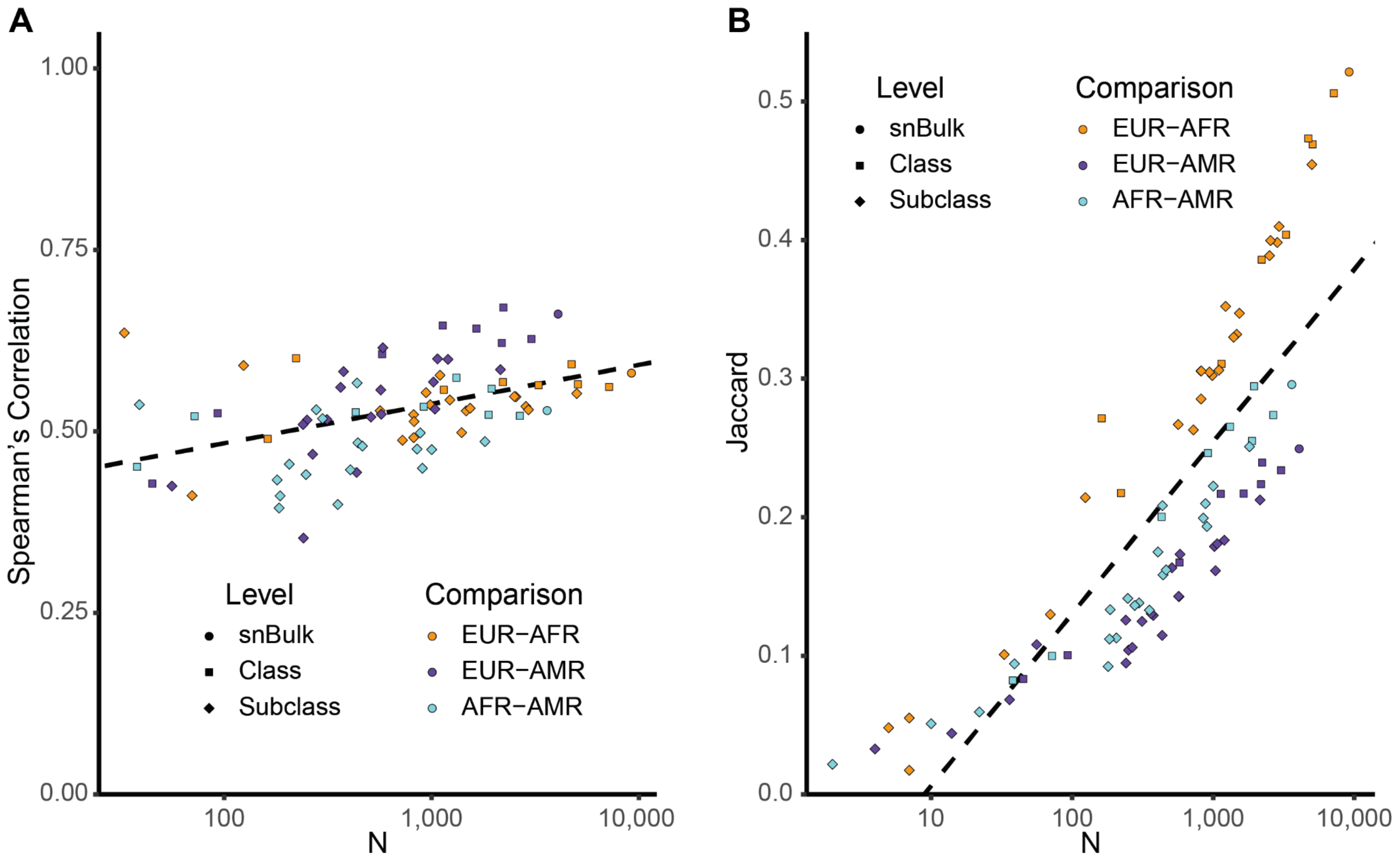

**Supplementary Fig. 2 | Spearman's correlation (A) and Jaccard similarity (B) of cross-ancestry prediction performance at different cell-type levels.** Each point represents a pairwise analysis (Spearman's correlation for A and Jaccard index for B, respectively) of $R^2_{CV}$ across all shared imputable genes ("N") between ancestries for the same snTIM. Applicable pairwise ancestry comparisons include EUR-AFR, EUR-AMR and AFR-AMR colored orange, purple and light-blue, respectively. Point shape denotes snTIM level including snBulk, Class and Subclass. Dashed lines represent linear regression lines. **A.** $\log_{10}(N)$ is positively associated ($p = 1.01\times10^{-5}$) with Spearman's correlation in the linear regression analysis ($p = 1.01\times10^{-5}$, adjusted $R^2 = 0.21$). **B.** $\log_{10}(N)$ is positively associated ($p = 1.91\times10^{-24}$) with the Jaccard index in a linear regression model ($p = 3.81\times10^{-24}$, adjusted $R^2 = 0.679$).

### Supplementary Fig. 3

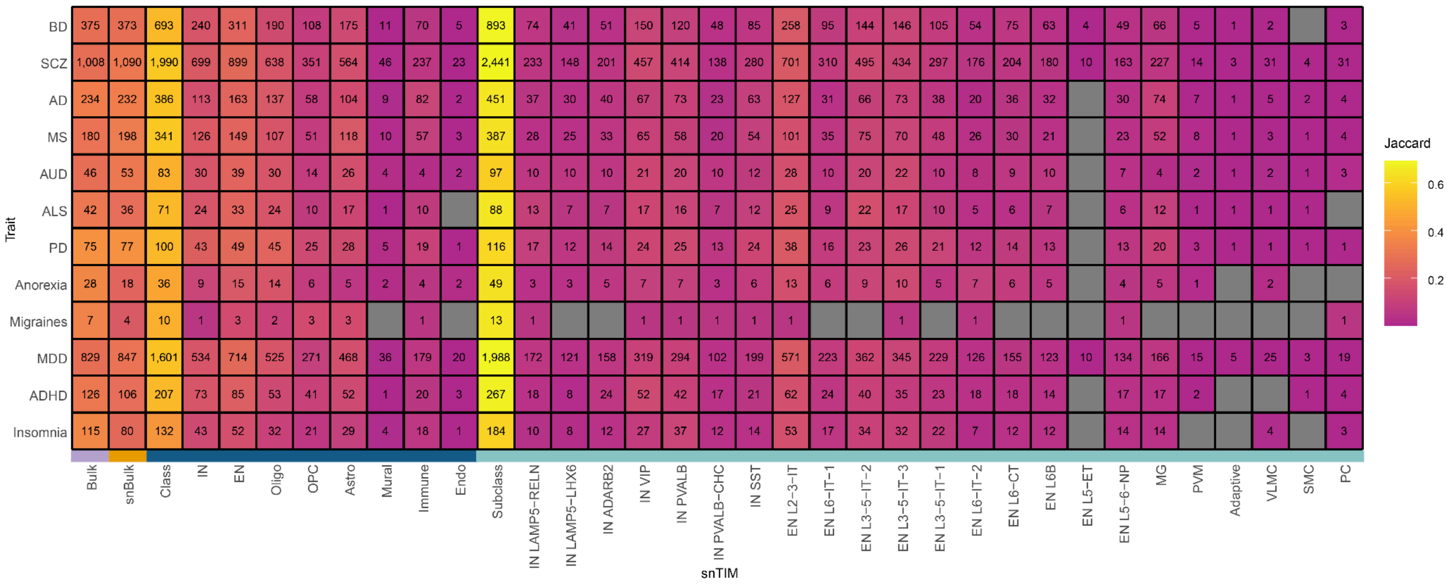

**Supplementary Fig. 3 | Expanded heatmap of S-TWAS significant gene-trait associations by cell-type hierarchy level.** The number within each cell represents the number of FDR[33]-significant gene-trait associations (GTAs) within each snTIM and level aggregate. FDR-correction for class and subclass aggregate categories is performed across all participating models for that hierarchy level. Color denotes the Jaccard index ($\frac{|x \cap all|}{|x \cup all|}$) of the specific cell ($x$) against the union of all FDR-significant trait-associated genes across Bulk, snBulk, class and subclass. S-TWAS analysis is limited to EUR ancestry.

### Supplementary Fig. 4

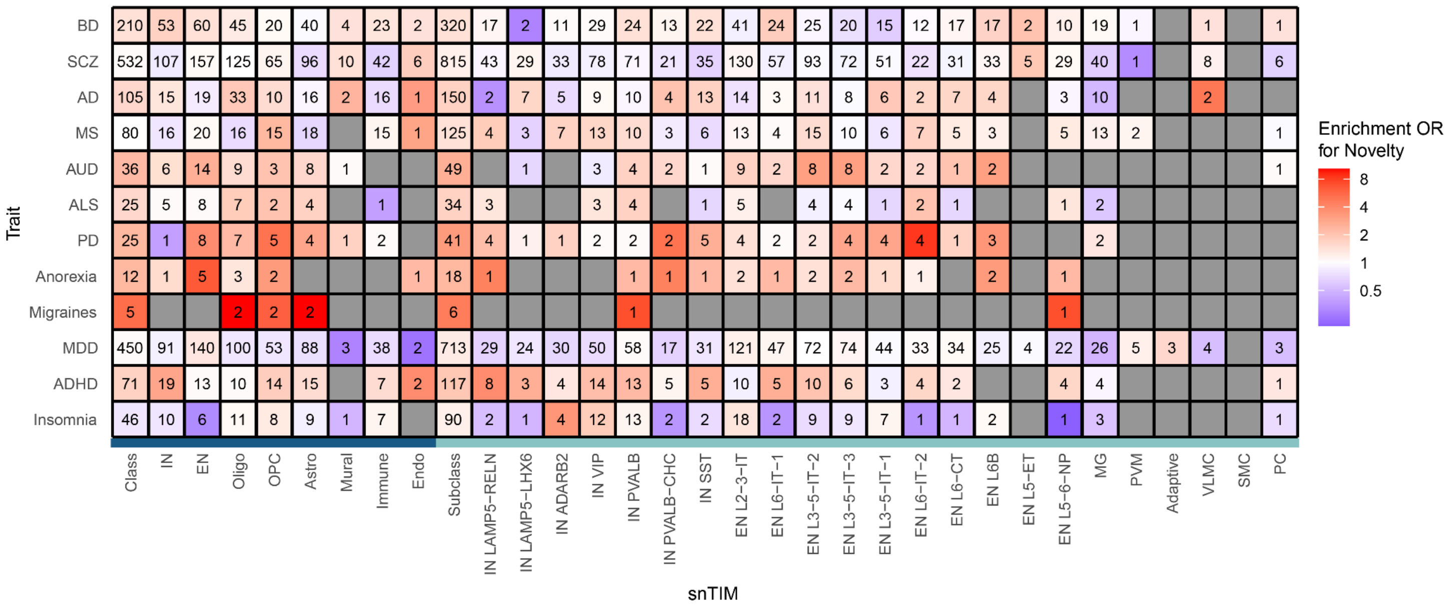

**Supplementary Fig. 4 | Heatmap of novel GTAs.** Number in each cell indicates the number of novel GTAs (as in Fig. 2C) found in each snTIM and level aggregates. Color of each cell indicates the effect size of the fisher's exact test comparing each snTIM to snBulk.

### Supplementary Fig. 5

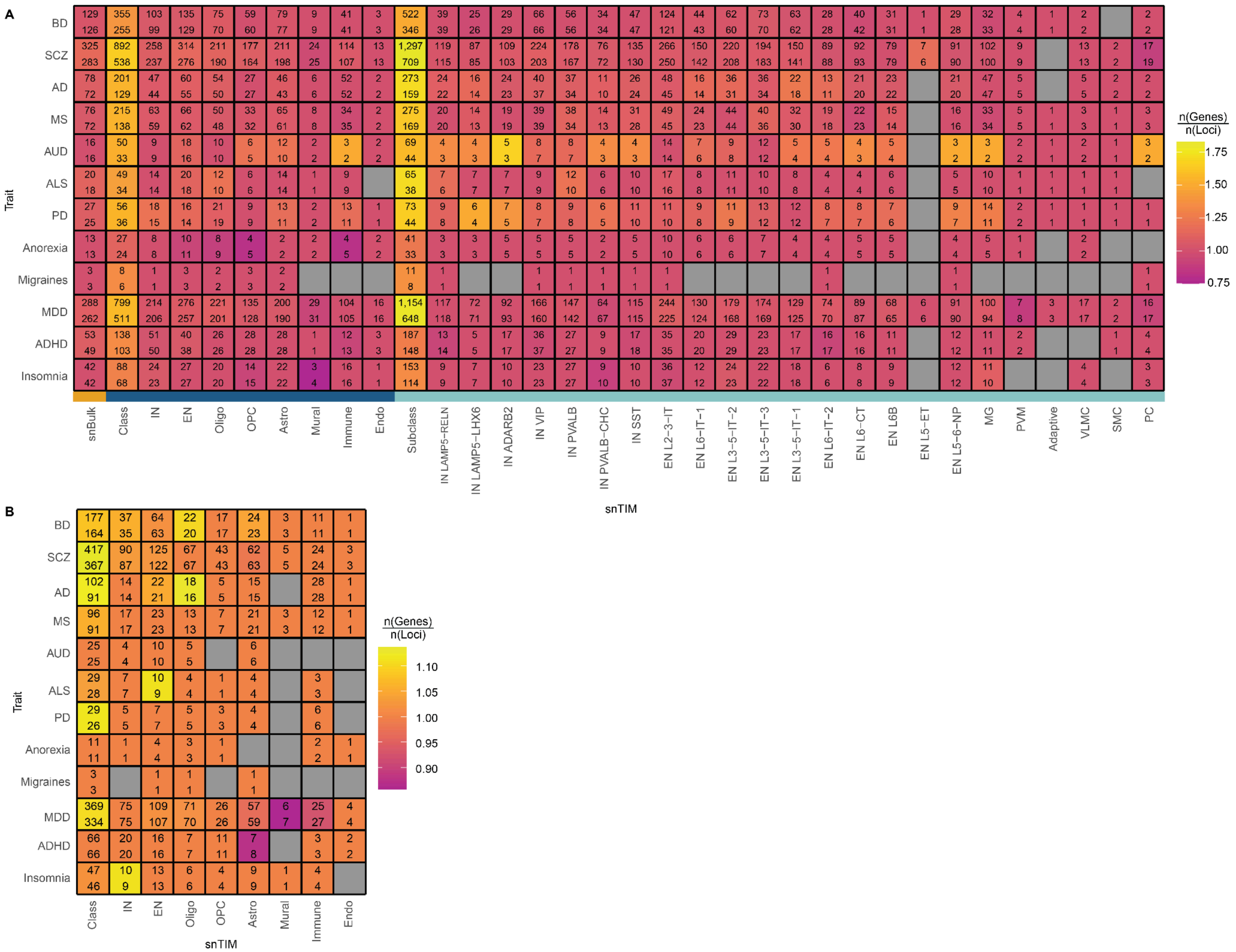

**Supplementary Fig. 5 | Expanded gene fine-mapping heatmaps. A,** Expanded heatmap of S-snTWAS individual-cell-type gene fine-mapping. The numbers at the top and bottom of each cell indicate the count of fine-mapped genes and loci, respectively. Color indicates the genes per locus for every trait-snTIM combination. Fine-mapping was performed with FOCUS for each cell-type. FOCUS performs analyses at the LD block level rather than the standard 1 mega base pair window from the transcription start site which means that dysregulated GReX for a given gene-trait combination may be driven by more than one locus. In our cell-type-specific snTWAS FOCUS analysis, in most loci one gene was fine-mapped (PIP ≥ 0.5), with the exception of a few loci prioritizing more than one gene. Interestingly, in a few instances (e.g. MDD PVM), a gene was fine-mapped in two different LD blocks. **B,** Expanded heatmap of S-snTWAS multiple-cell-type gene fine-mapping. The numbers at the top and bottom of each cell indicate the count of fine-mapped genes and loci, respectively. Color indicates the genes per locus for every trait-snTIM combination. Fine-mapping was performed with FOCUS while jointly considering all cell-types at the class level.

### Supplementary Fig. 6

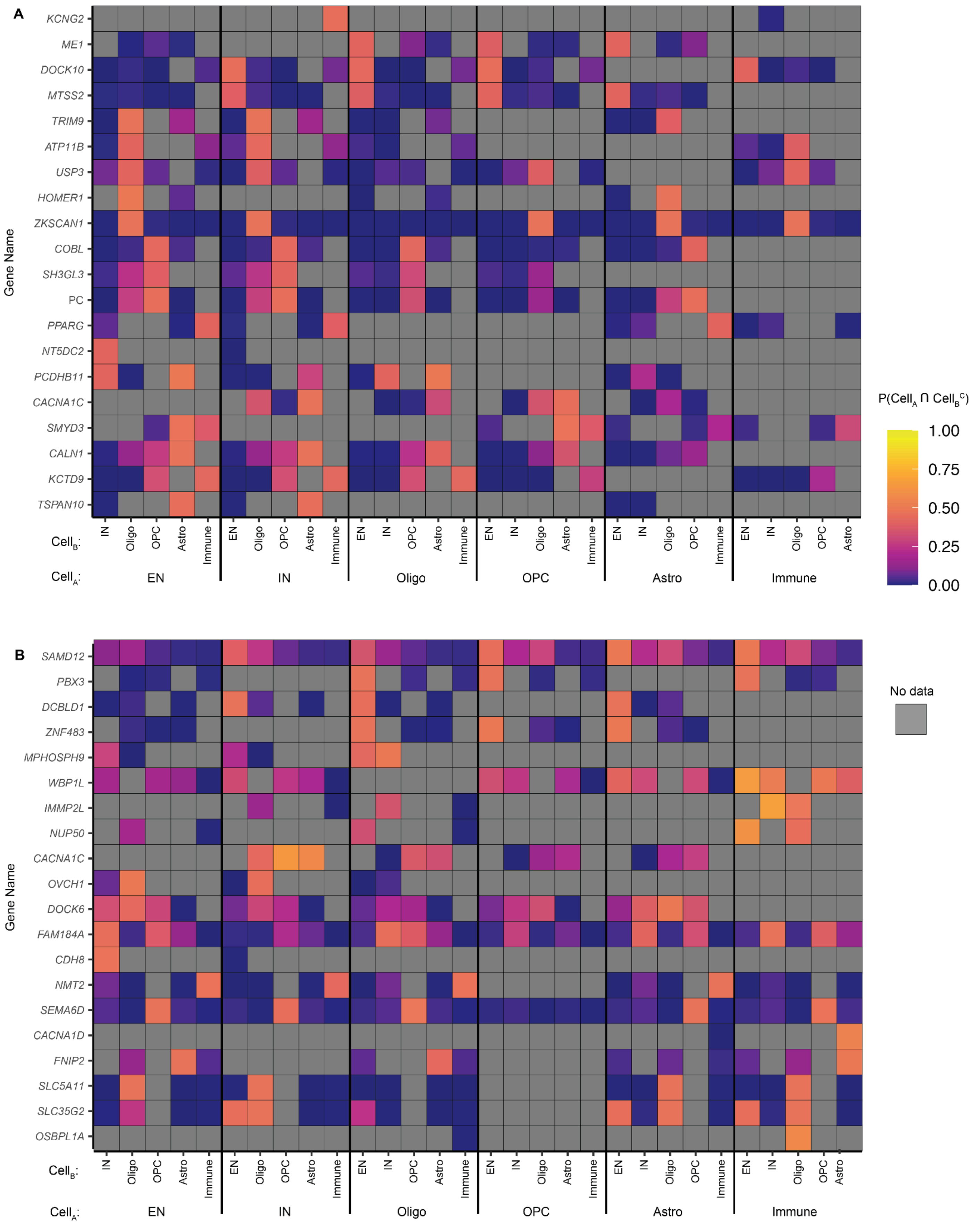

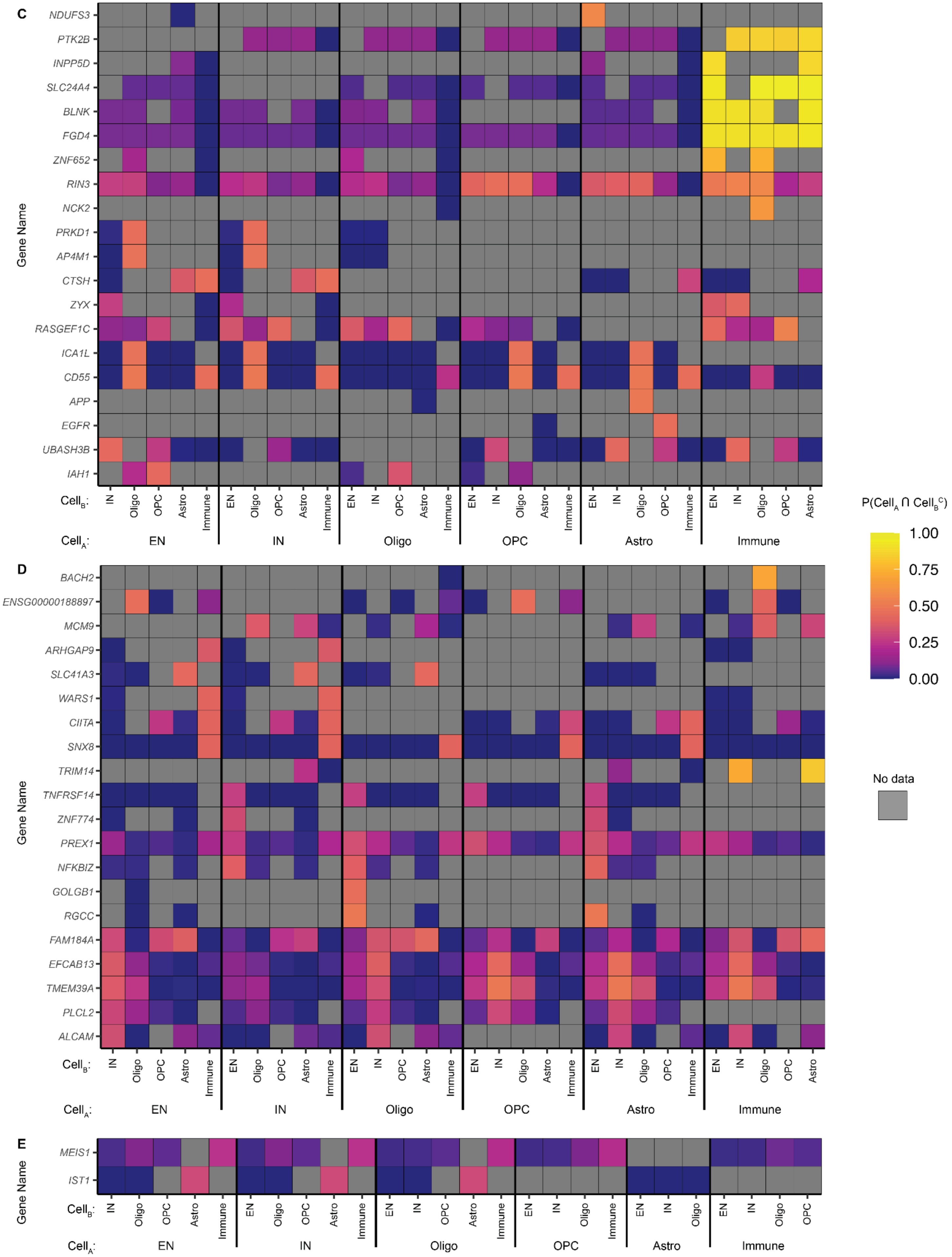
C
NDUFS3
PTK2B
INPP5D
SLC24A4
BLNK
FGD4
ZNF652
RIN3
NCK2
PRKD1
AP4M1
CTSH
ZYX
RASGEF1C
ICA1L
CD55
APP
EGFR
UBASH3B
IAH1
Gene Name
Cell_B:
Cell_A:
EN
IN
Oligo
OPC
Astro
Immune
D
BACH2
ENSG00000188897
MCM9
ARHGAP9
SLC41A3
WARS1
CIITA
SNX8
TRIM14
TNFRSF14
ZNF774
PREX1
NFKBIZ
GOLGB1
RGCC
FAM184A
EFCAB13
TMEM39A
PLCL2
ALCAM
E
MEIS1
IST1
P(Cell_A ∩ Cell_B^C)
1.00
0.75
0.50
0.25
0.00
No data

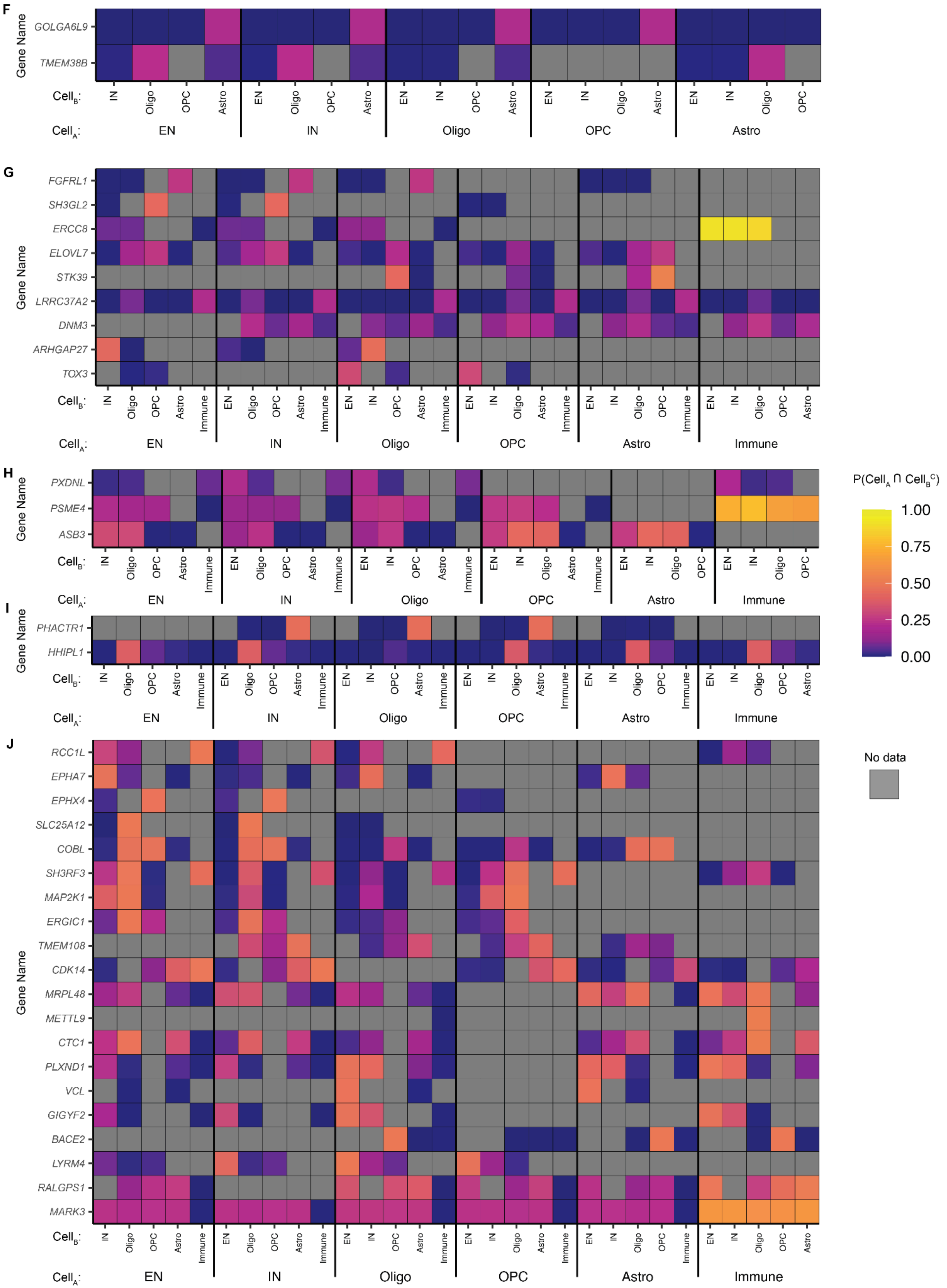

F
G
H
I
J
Gene Name
GOLGA6L9
TMEM38B
FGFRL1
SH3GL2
ERCC8
ELOVL7
STK39
LRRC37A2
DNM3
ARHGAP27
TOX3
PXDNL
PSME4
ASB3
PHACTR1
HHIPL1
RCC1L
EPHA7
EPHX4
SLC25A12
COBL
SH3RF3
MAP2K1
ERGIC1
TMEM108
CDK14
MRPL48
METTL9
CTC1
PLXND1
VCL
GIGYF2
BACE2
LYRM4
RALGPS1
MARK3
Cell$_B$:
Cell$_A$:
EN
IN
Oligo
OPC
Astro
Immune
P(Cell$_A$ ∩ Cell$_B^C$)
1.00
0.75
0.50
0.25
0.00
No data

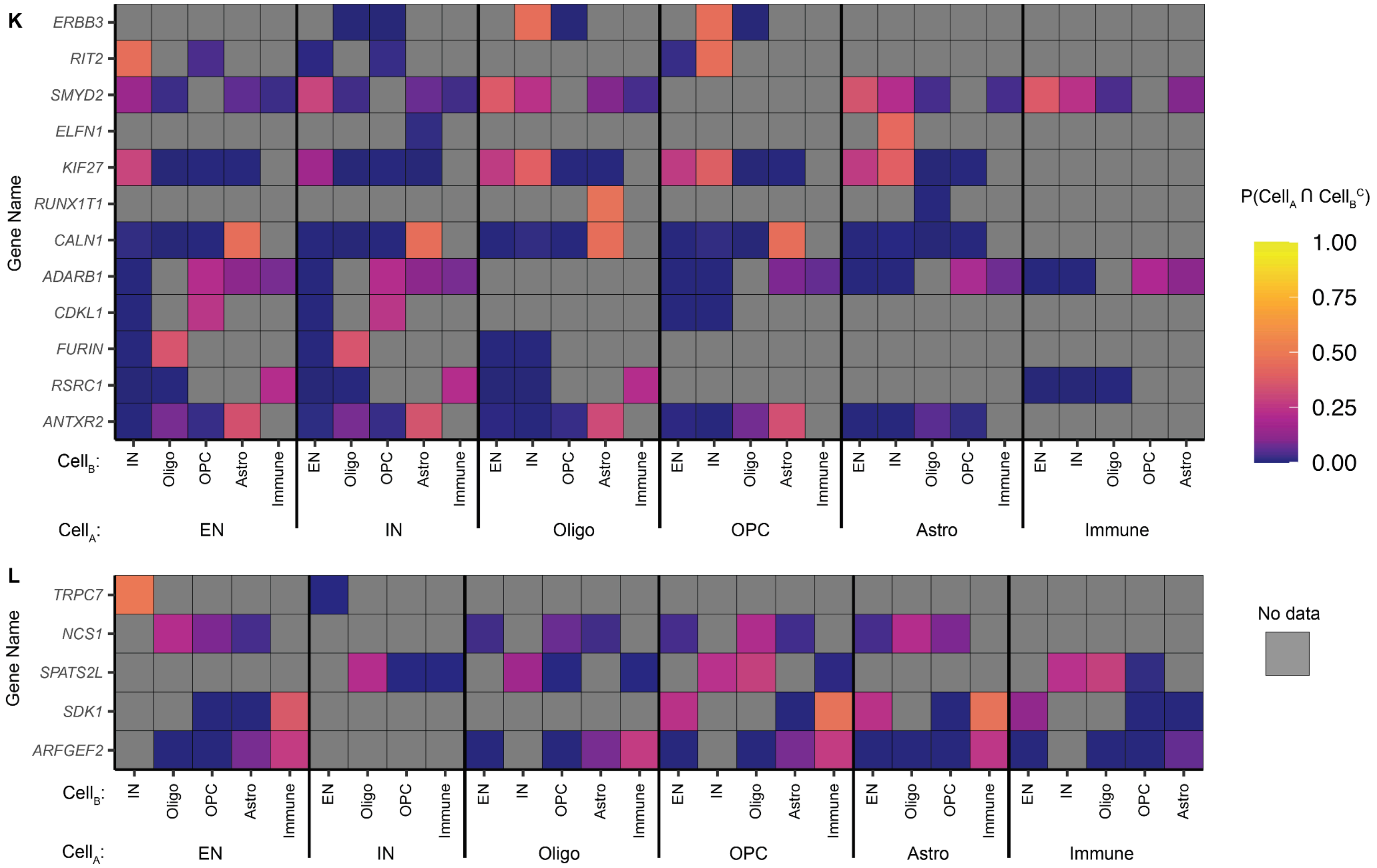

**Supplementary Fig. 6 | Multivariate adaptive shrinkage (mash) analysis in S-snTWAS traits.** Heatmaps depicting the combinatorial probability that an effect exists in cell-type A and not in cell-type B at the class level. Only results with a $P(Cell_A \cap Cell_B^C) \geq 0.2$ in at least one comparison were retained for visualization. The top 20 results (ordered by the maximum row-wise combinatorial probability) are displayed for BD (A), SCZ (B), AD (C), MS (D), AUD (E), ALS (F), PD (G), Anorexia (H), Migraines (I), MDD (J), ADHD (K), Insomnia (L). Cell type probabilities are estimated with mash. Cells are colored gray when the analysis is not applicable.

### Supplementary Fig. 7

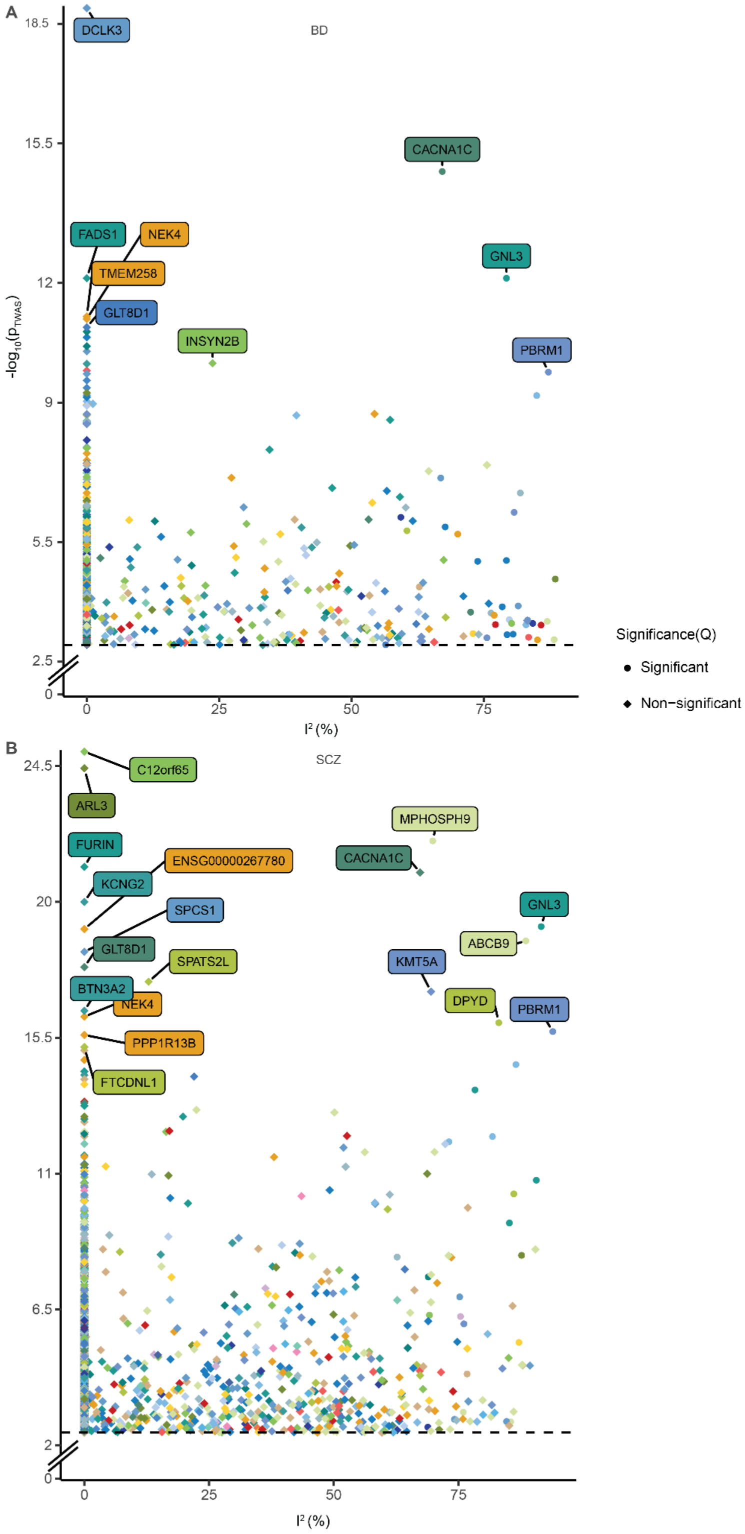

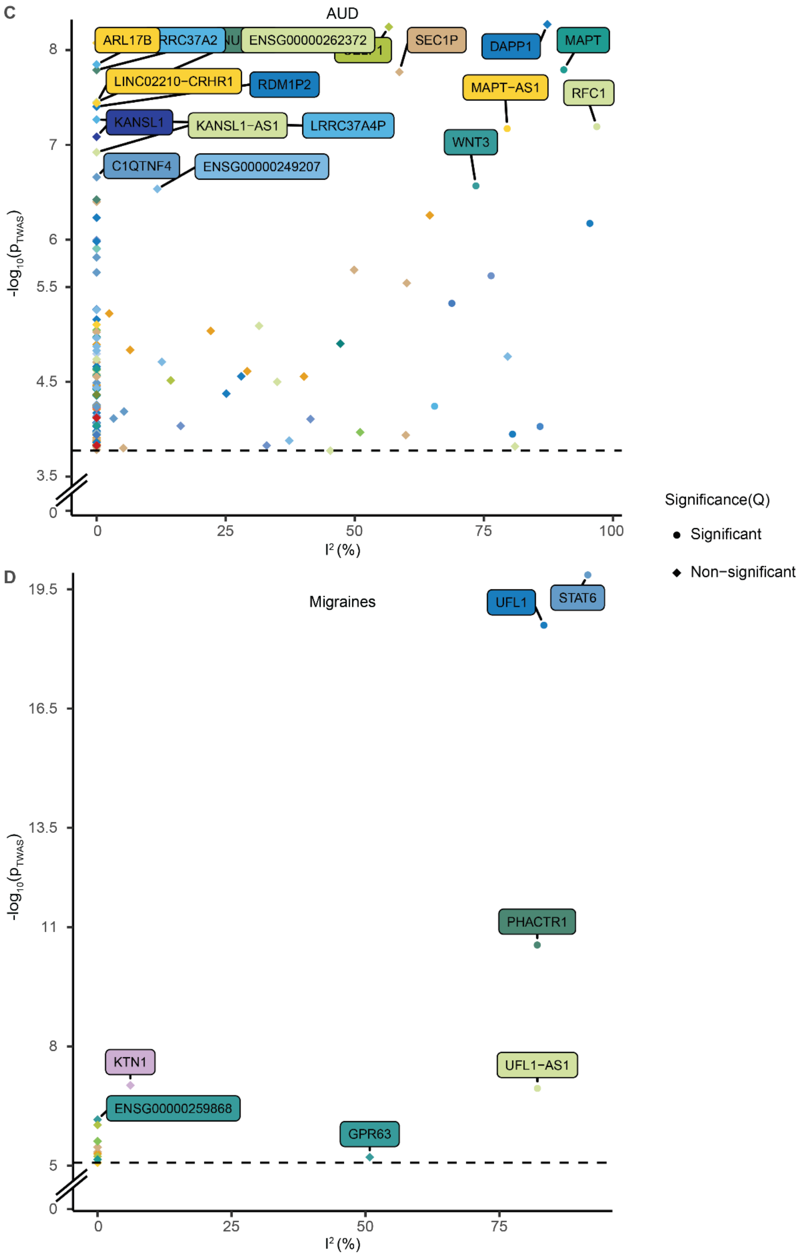
C
AUD
ARL17B
RRC37A2
ENSG00000262372
SEC1P
DAPP1
MAPT
LINC02210−CRHR1
RDM1P2
MAPT−AS1
RFC1
KANSL1
KANSL1−AS1
LRRC37A4P
WNT3
C1QTNF4
ENSG00000249207
$-\log_{10}(p_{TWAS})$
$I^2$ (%)
Significance(Q)
Significant
Non−significant
D
Migraines
UFL1
STAT6
PHACTR1
KTN1
UFL1−AS1
ENSG00000259868
GPR63
$-\log_{10}(p_{TWAS})$
$I^2$ (%)

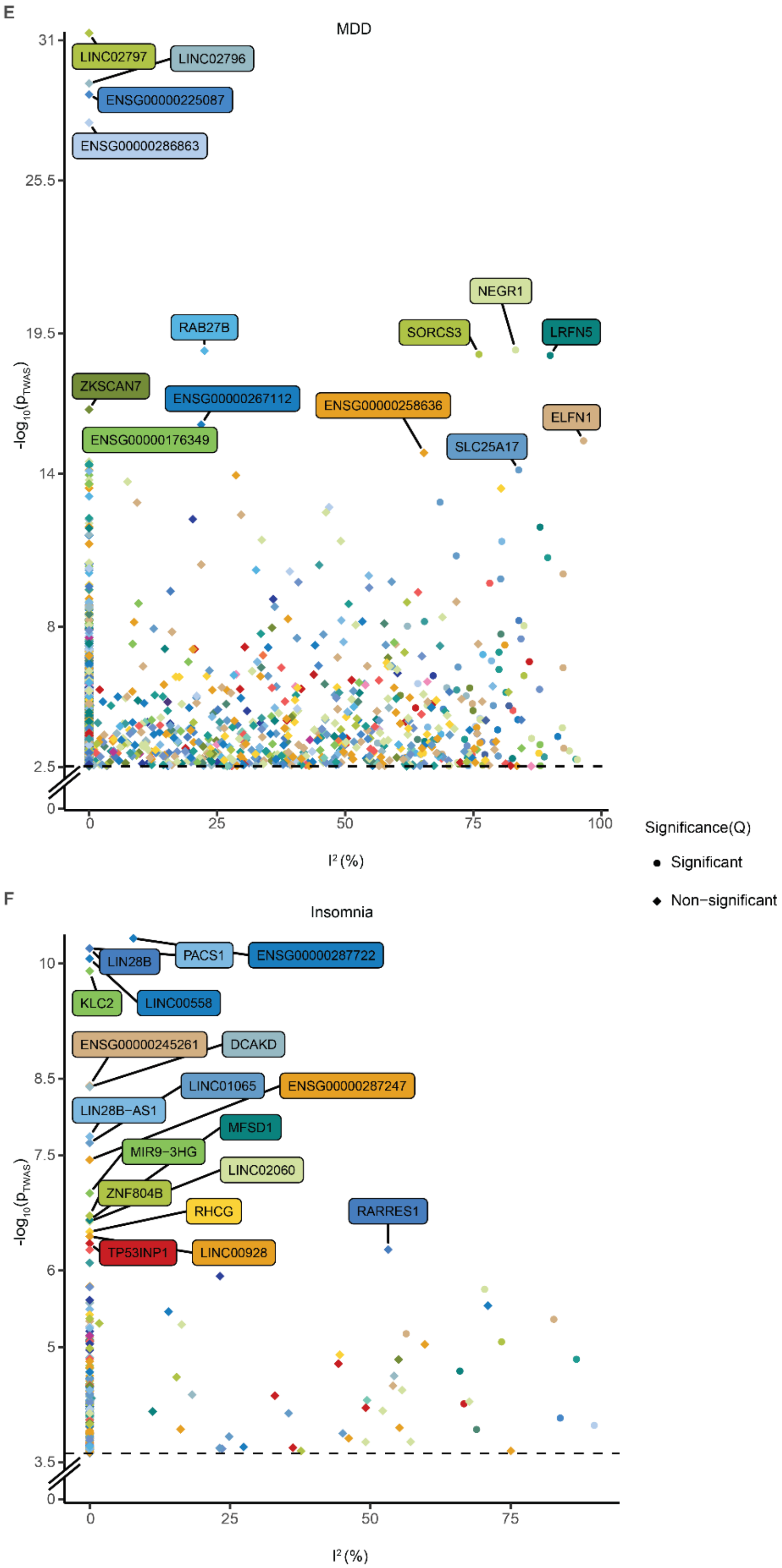

E
MDD
31
25.5
19.5
14
8
2.5
0
-log10(pTWAS)
0
25
50
75
100
I² (%)
LINC02797
LINC02796
ENSG00000225087
ENSG00000286863
NEGR1
SORCS3
LRFN5
RAB27B
ZKSCAN7
ENSG00000267112
ENSG00000258636
ELFN1
ENSG00000176349
SLC25A17
Significance(Q)
Significant
Non−significant
F
Insomnia
10
8.5
7.5
6
5
3.5
0
-log10(pTWAS)
0
25
50
75
I² (%)
LIN28B
PACS1
ENSG00000287722
KLC2
LINC00558
ENSG00000245261
DCAKD
LINC01065
ENSG00000287247
LIN28B-AS1
MFSD1
MIR9-3HG
LINC02060
ZNF804B
RHCG
RARRES1
TP53INP1
LINC00928

**Supplementary Fig. 7 | Gene-trait association effect size heterogeneity scatterplots**. Gene association effect size heterogeneity ($I^2$) across cell types for BD (**A**), SCZ (**B**), AUD (**C**), Migraines (**D**), MDD (**E**), Insomnia (**F**). Significance denotes the presence of different effects of individual cell types in the analysis (Cochran's Q test). Only genes with significant GTAs in S-snTWAS analysis are visualized. Color corresponds to cell-type (Extended Data Fig. 2). Only traits with a high enough sample size in the MVP (see Methods) were evaluated for heterogeneity, due to need for effect size estimates. AD is visualized in Fig. 3C. ADHD, ALS, Anorexia, MS, and PD are excluded due to few cases in the MVP. PTSD and Anxiety are excluded due to few significant S-snTWAS associations.

Supplementary Fig. 8

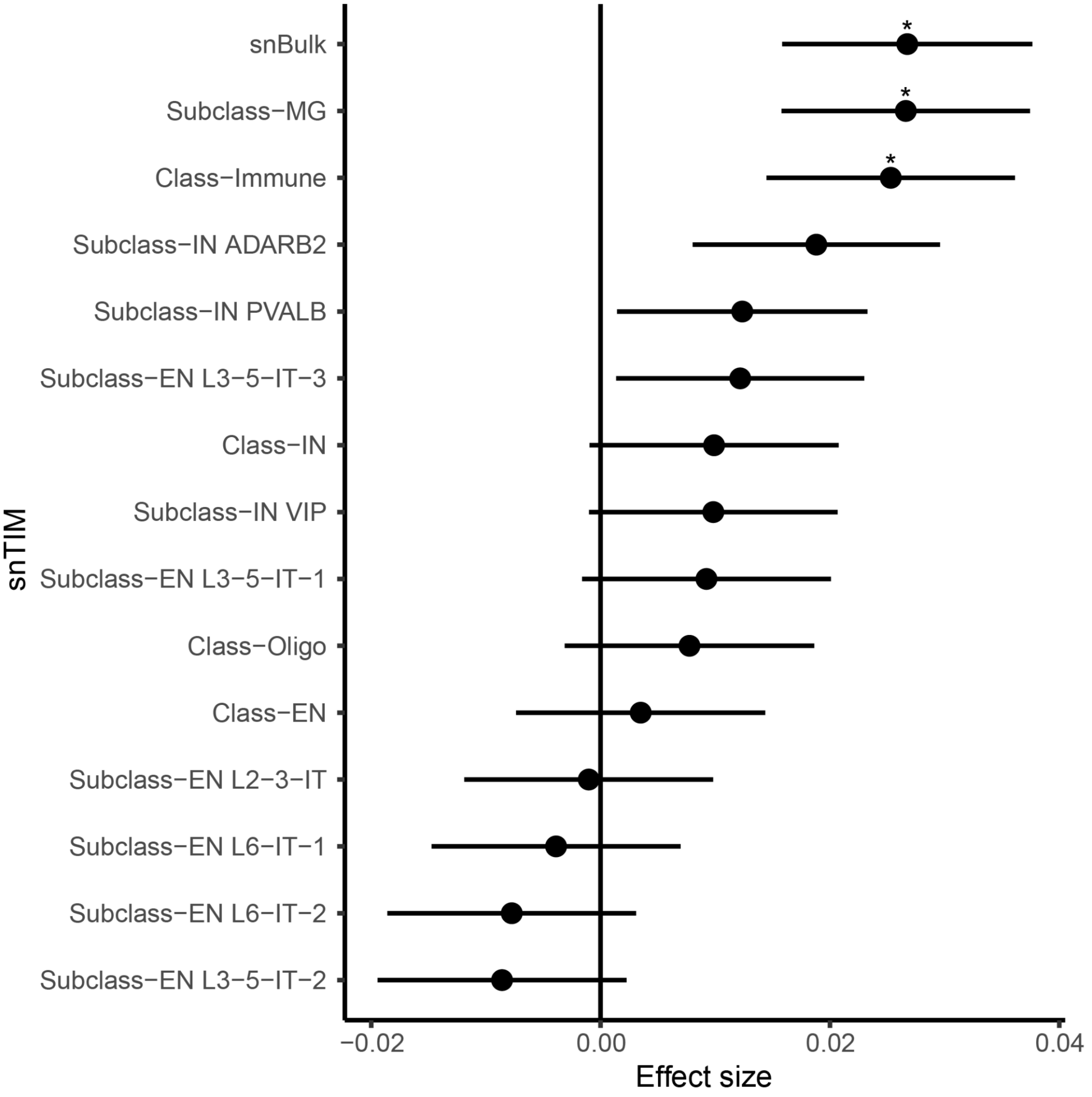

**Supplementary Fig. 8 | GTA:*BIN1*-AD Forest Plot.** Significant associations are marked by an asterisk. Horizontal bars denote the 95% confidence interval.

### Supplementary Fig. 9

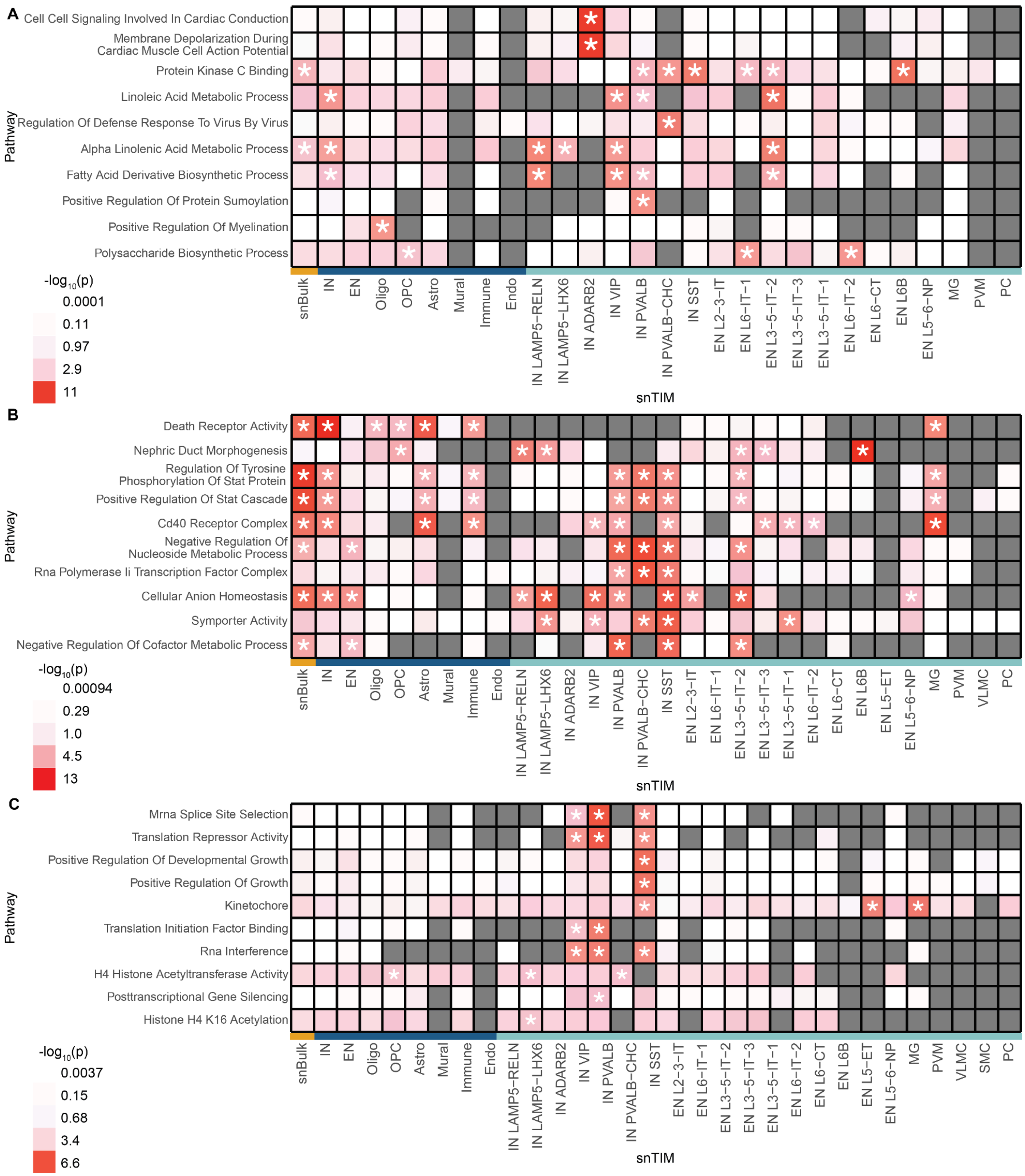

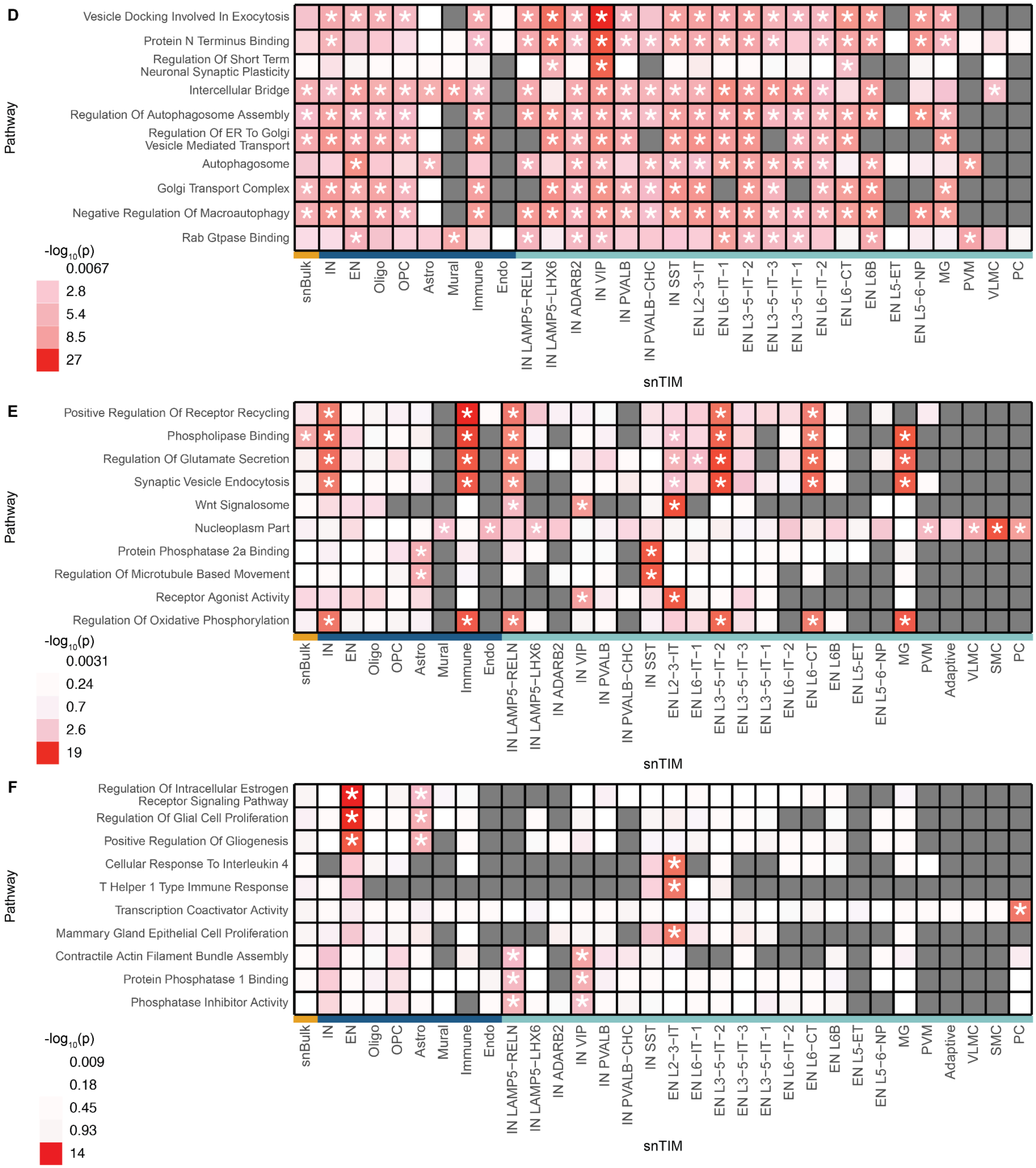
D
Vesicle Docking Involved In Exocytosis
Protein N Terminus Binding
Regulation Of Short Term Neuronal Synaptic Plasticity
Intercellular Bridge
Regulation Of Autophagosome Assembly
Regulation Of ER To Golgi Vesicle Mediated Transport
Autophagosome
Golgi Transport Complex
Negative Regulation Of Macroautophagy
Rab Gtpase Binding
Pathway
-log10(p)
0.0067
2.8
5.4
8.5
27
snBulk
IN
EN
Oligo
OPC
Astro
Mural
Immune
Endo
IN LAMP5−RELN
IN LAMP5−LHX6
IN ADARB2
IN VIP
IN PVALB
IN PVALB−CHC
IN SST
EN L2−3−IT
EN L6−IT−1
EN L3−5−IT−2
EN L3−5−IT−3
EN L3−5−IT−1
EN L6−IT−2
EN L6−CT
EN L6B
EN L5−ET
EN L5−6−NP
MG
PVM
VLMC
PC
snTIM
E
Positive Regulation Of Receptor Recycling
Phospholipase Binding
Regulation Of Glutamate Secretion
Synaptic Vesicle Endocytosis
Wnt Signalosome
Nucleoplasm Part
Protein Phosphatase 2a Binding
Regulation Of Microtubule Based Movement
Receptor Agonist Activity
Regulation Of Oxidative Phosphorylation
Pathway
-log10(p)
0.0031
0.24
0.7
2.6
19
snBulk
IN
EN
Oligo
OPC
Astro
Mural
Immune
Endo
IN LAMP5−RELN
IN LAMP5−LHX6
IN ADARB2
IN VIP
IN PVALB
IN PVALB−CHC
IN SST
EN L2−3−IT
EN L6−IT−1
EN L3−5−IT−2
EN L3−5−IT−3
EN L3−5−IT−1
EN L6−IT−2
EN L6−CT
EN L6B
EN L5−ET
EN L5−6−NP
MG
PVM
Adaptive
VLMC
SMC
PC
snTIM
F
Regulation Of Intracellular Estrogen Receptor Signaling Pathway
Regulation Of Glial Cell Proliferation
Positive Regulation Of Gliogenesis
Cellular Response To Interleukin 4
T Helper 1 Type Immune Response
Transcription Coactivator Activity
Mammary Gland Epithelial Cell Proliferation
Contractile Actin Filament Bundle Assembly
Protein Phosphatase 1 Binding
Phosphatase Inhibitor Activity
Pathway
-log10(p)
0.009
0.18
0.45
0.93
14
snBulk
IN
EN
Oligo
OPC
Astro
Mural
Immune
Endo
IN LAMP5−RELN
IN LAMP5−LHX6
IN ADARB2
IN VIP
IN PVALB
IN PVALB−CHC
IN SST
EN L2−3−IT
EN L6−IT−1
EN L3−5−IT−2
EN L3−5−IT−3
EN L3−5−IT−1
EN L6−IT−2
EN L6−CT
EN L6B
EN L5−ET
EN L5−6−NP
MG
PVM
Adaptive
VLMC
SMC
PC
snTIM

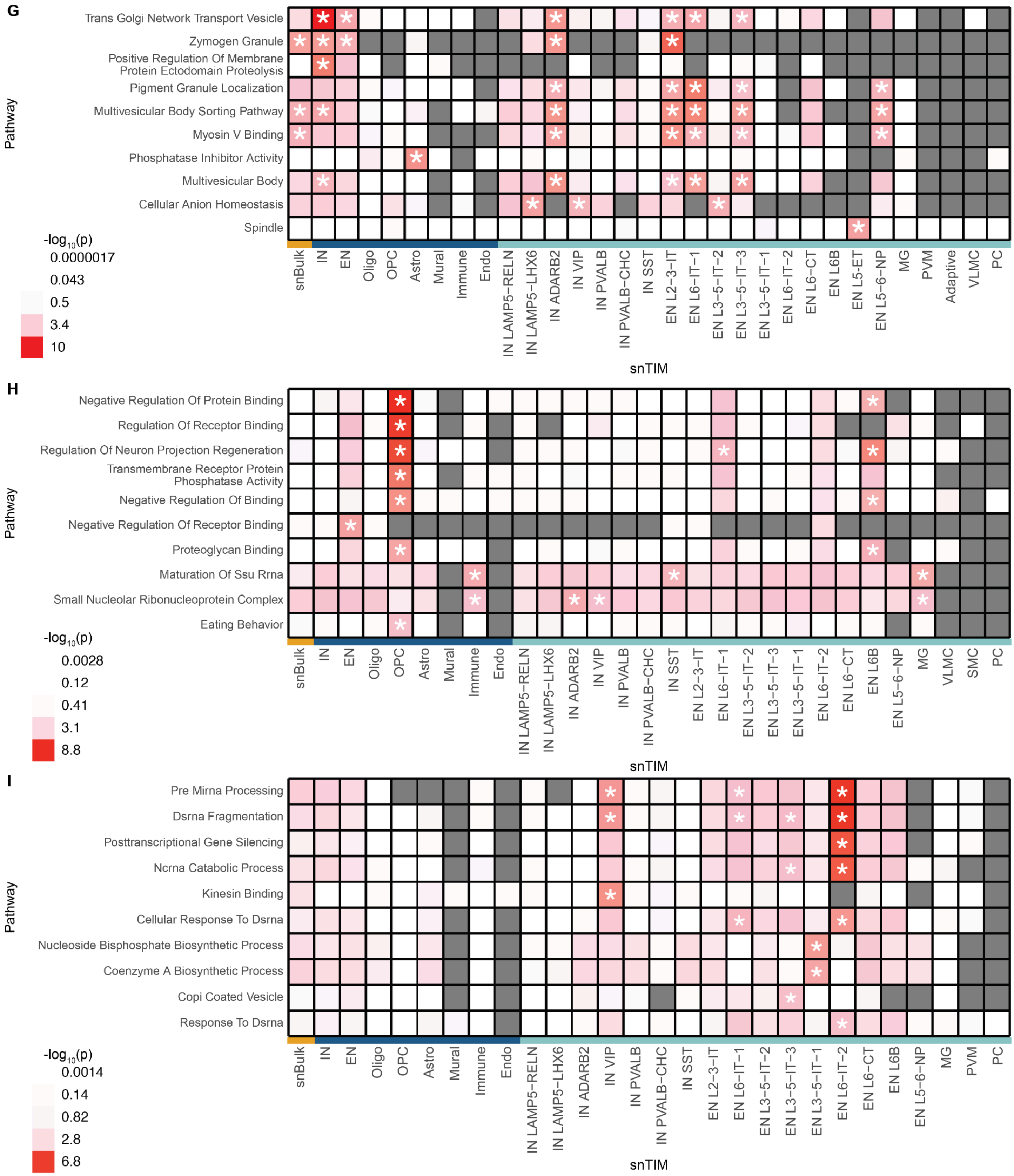

**Supplementary Fig. 9 | Top 10 pathways enriched within all S-snTWAS traits.** Pathways were prioritized by p-value. Hierarchical pruning was performed to deprioritize pathways (see Methods). For each cell-type, the top 2 significant pathways after hierarchical pruning (if any) were prioritized.

We describe the following traits in each panel: BD (**A**), MS (**B**), AUD (**C**), ALS (**D**), PD (**E**), Migraines (**F**), MDD (**G**), ADHD (**H**), Insomnia (**I**). AD and SCZ are described within Extended Data Fig. 5B.

### Supplementary Fig. 10

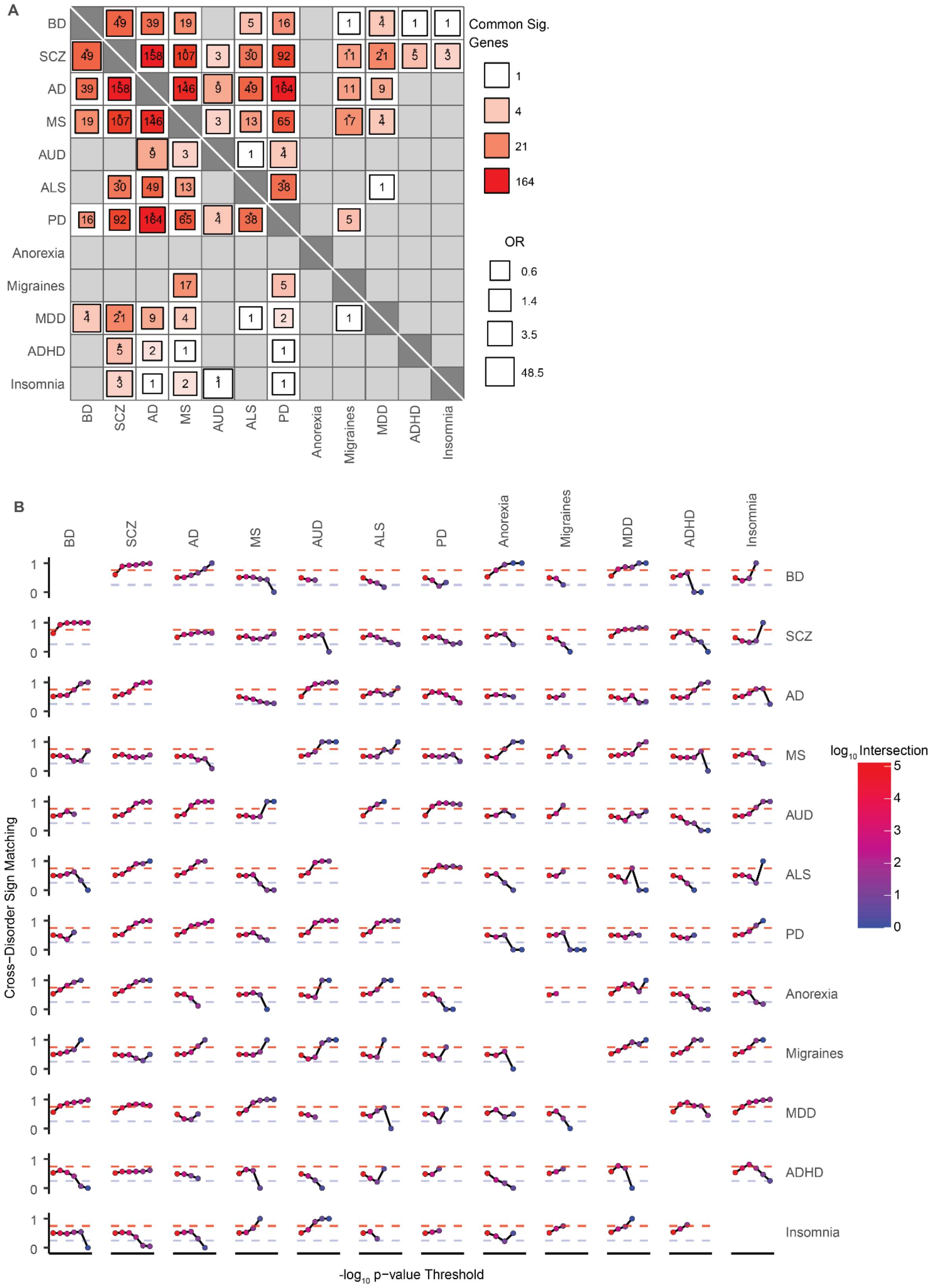

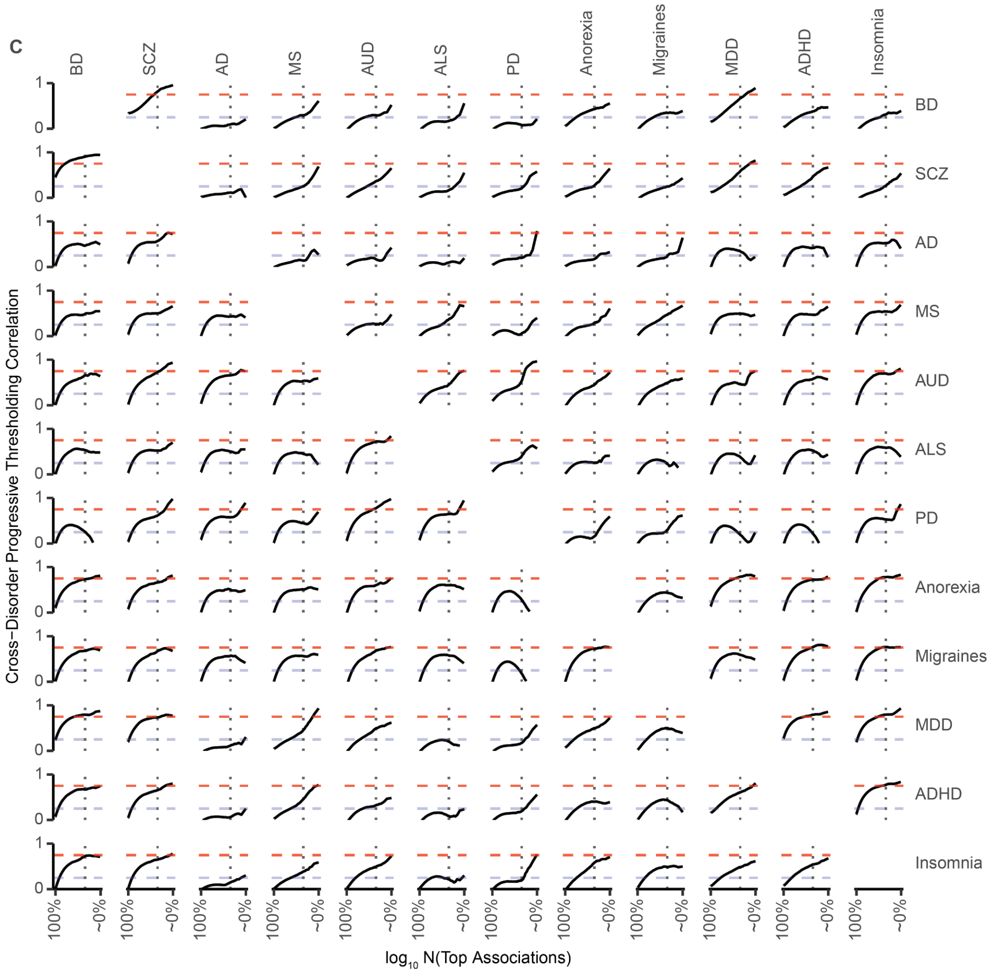

**Supplementary Fig. 10 | Cross-disorder concordance. A,** Fisher's exact test Heatmap of cross-disorder significant gene sharing. *Lower-Left Triangle*: The number in each cell describes the number of shared significant genes (*lower-left triangle*) or pathways (*upper-right triangle*) between each pair of disorders. Fisher's exact test p-values were FDR[33]-corrected, and significant values are annotated by asterisks. **B,** Cross-disorder association concordance. Each point represents the percent of concordant gene-cell-type (*Lower-Left Triangle*) or pathway-cell-type (*Upper-Right Triangle*) combinations (same association z-score sign) under the appropriate p-value threshold in both disorders. Each progressive point restricts the p-value threshold 10-fold (i.e. $p \leq 1$, $p \leq 1 \times 10^{-1}$, $p \leq 1 \times 10^{-2}$, …). Red and blue dashed horizontal lines denote a 0.75 (75%) and 0.25 (25%) sign concordance, respectively. **C,** snTWAS cross-disorder progressive thresholding correlation analysis (PTCA). Each line graph tracks the cross-disorder association z-score Pearson's correlation for progressively higher ranked (see Methods) gene-cell-type (*Lower-Left Triangle*) or pathway-cell-type (*Upper-Right Triangle*) combinations. Red and blue dashed horizontal lines denote a 0.75 and 0.25 Pearson's correlation coefficient (r), respectively. Vertical dotted lines denote top 1% of gene-cell-type and pathway-cell-type combinations for the lower-left and upper-right triangles, respectively.

Supplementary Fig. 11

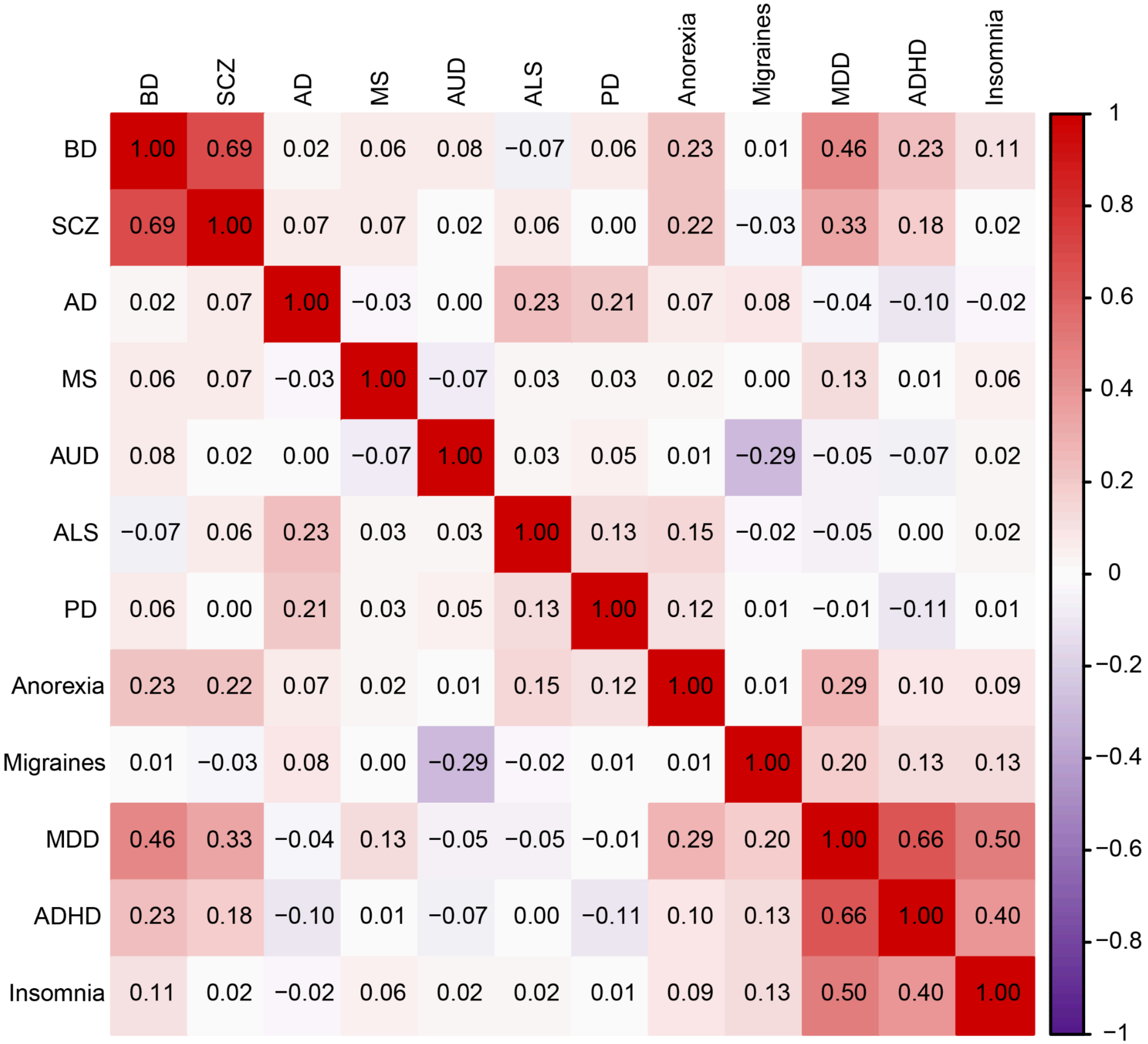

**Supplementary Fig. 11 | LD Score Regression bivariate heritability analysis.** Heatmap values range from -1 to 1 to indicate heritability correlation. Red indicates positive correlation, violet indicates negative correlation.

### Supplementary Fig. 12

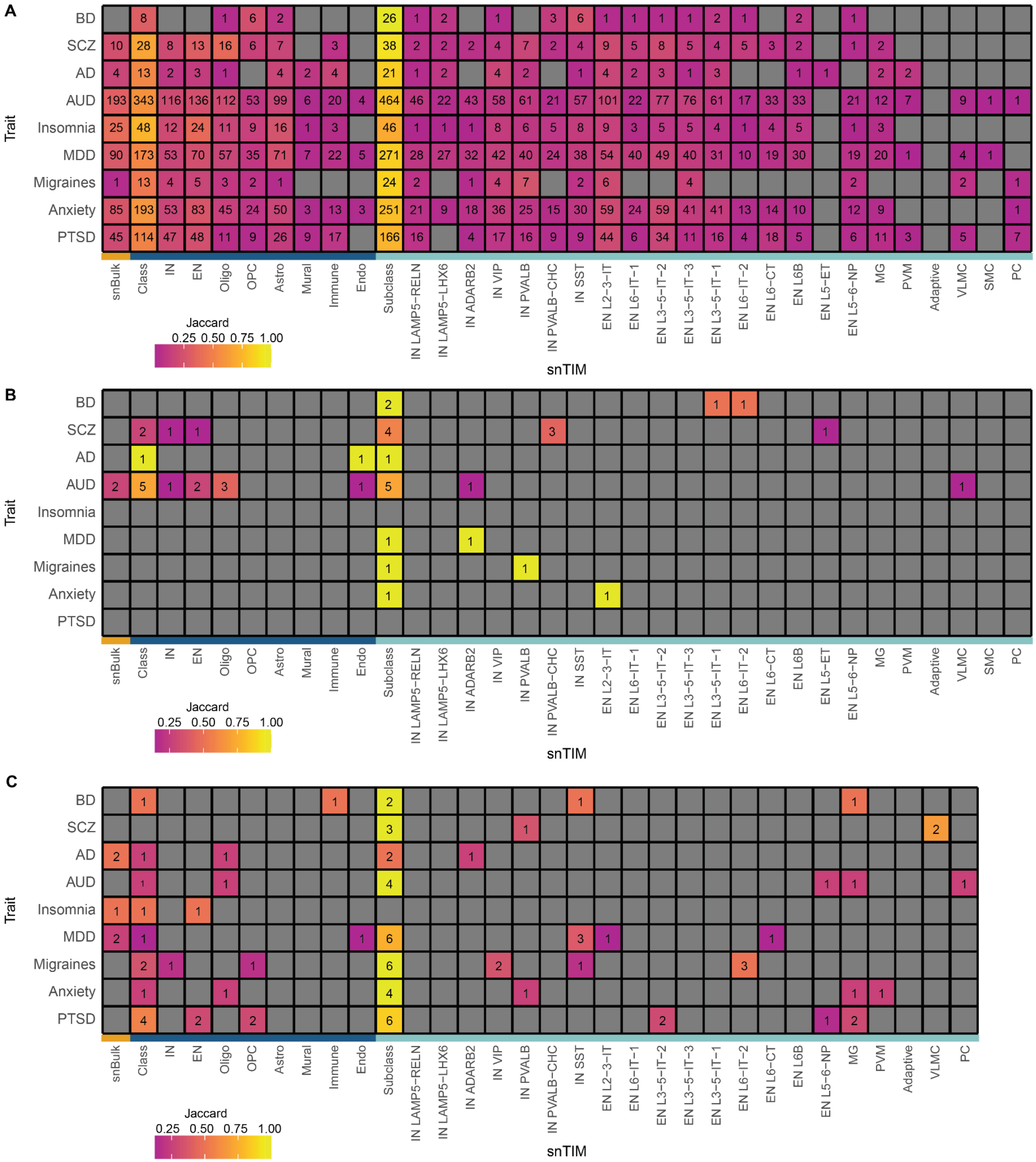

**Supplementary Fig. 12 | I-snTWAS GTA Heatmap for EUR (A), AFR (B) and AMR (C) ancestries.** The number within each cell represents the number of FDR[33]-significant gene-trait associations for the respective snTIM. “Class” and “Subclass” represent the number of FDR-significant genes among all class- and subclass-level snTIMs, respectively.

Supplementary Fig. 13

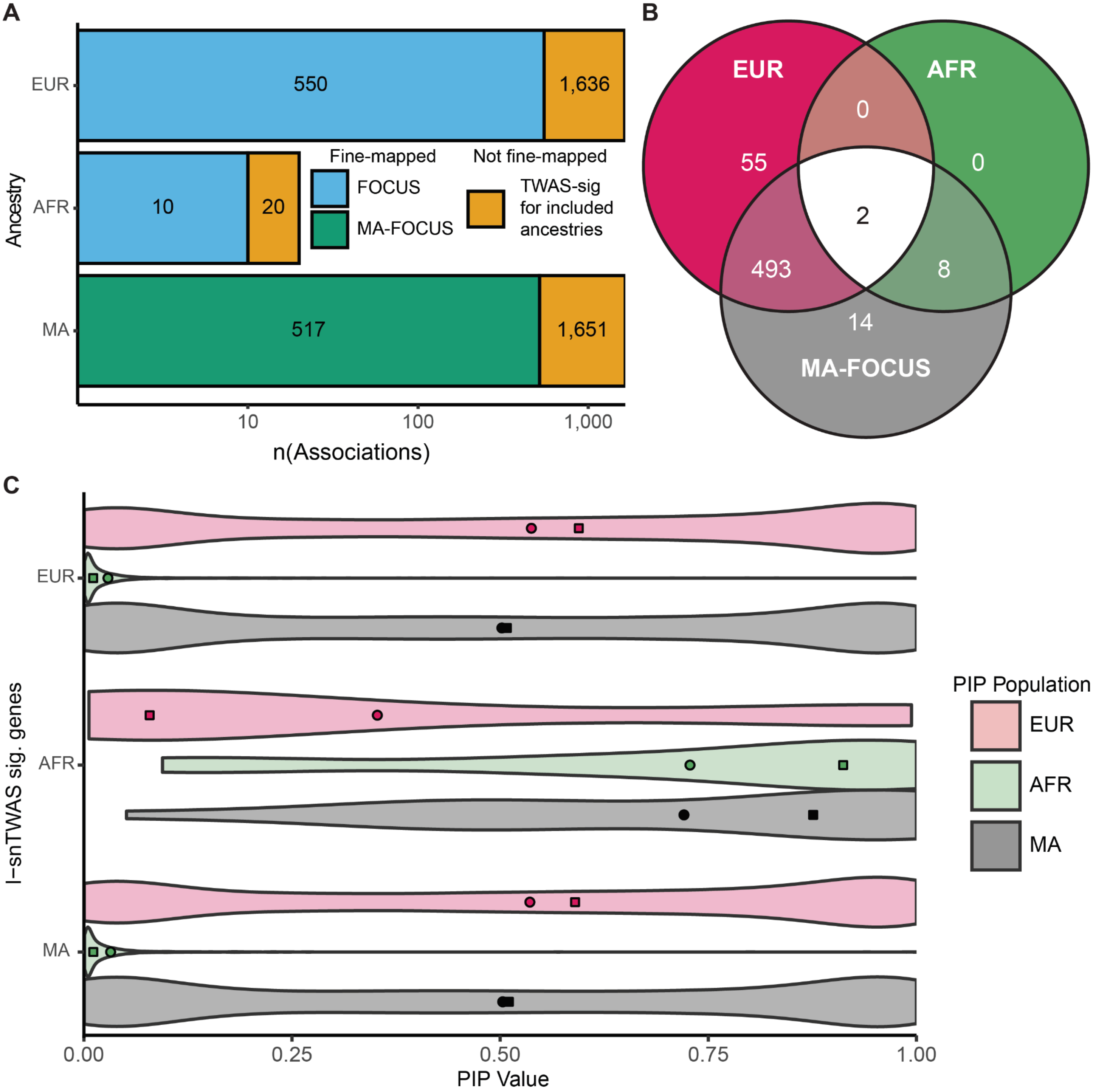

**Supplementary Fig. 13 | Overlap of cross-ancestry fine-mapping. A,** Overlap of TWAS and fine-mapping in single ancestry and multi-ancestry fine-mapping across the 9 I-snTWAS traits. Annotated numbers in the barplot indicate the total number of GTAs in each category (i.e. there are 550 fine-mapped GTAs in EUR and 1,636 I-snTWAS significant GTAs). Y-axis indicates the population in which analysis was performed. “MA” indicates the MA-FOCUS bi-ancestral analysis and the union of EUR and AFR I-snTWAS significant trait-gene-cell-type combinations for fine-mapping and TWAS, respectively. **B,** Overlap of fine-mapped GTAs (FOCUS for EUR and AFR; MA-FOCUS for bi-ancestral EUR and AFR). **C,** Distribution of PIP values (FOCUS for EUR and AFR, MA-FOCUS for bi-ancestral EUR and AFR) amongst **I**-snTWAS significant trait-gene-cell-type combinations for each ancestry ("MA" indicates the union of EUR and AFR **I**-snTWAS significant trait-gene-cell-type combinations). Square points indicate the median values of each distribution and circular points indicate the mean values of each distribution.

Supplementary Fig. 14

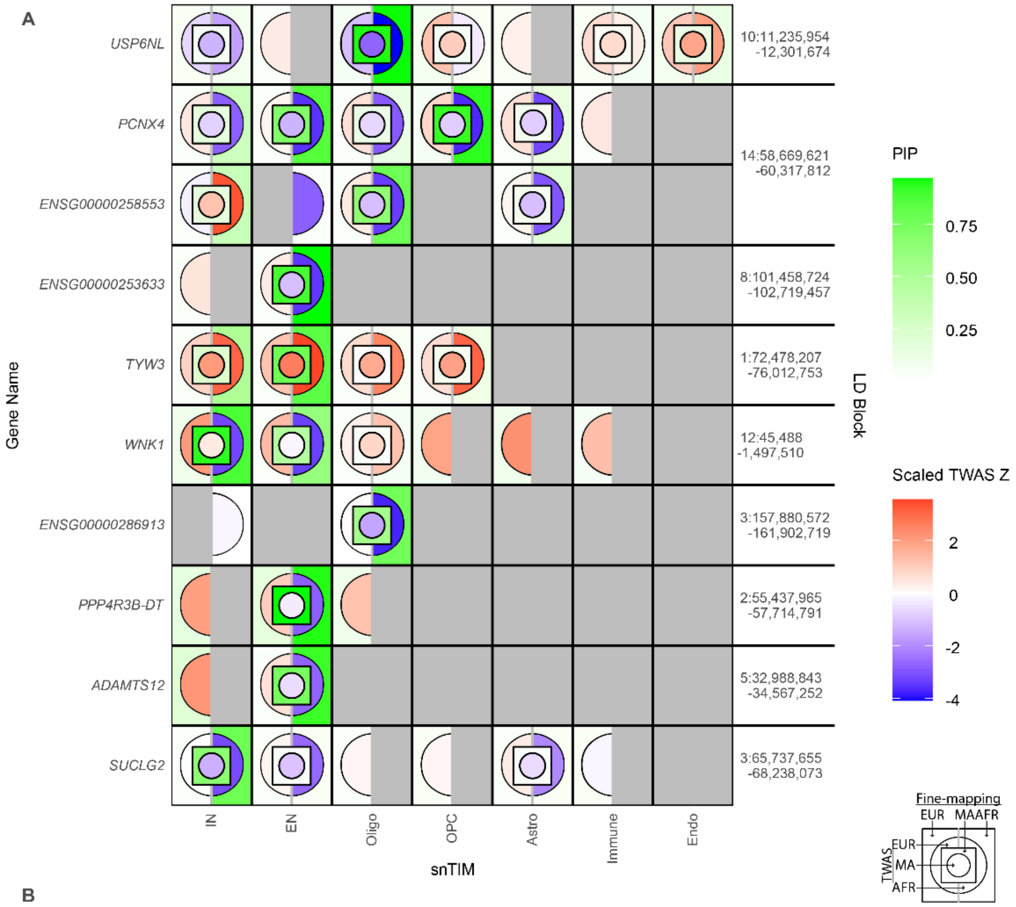

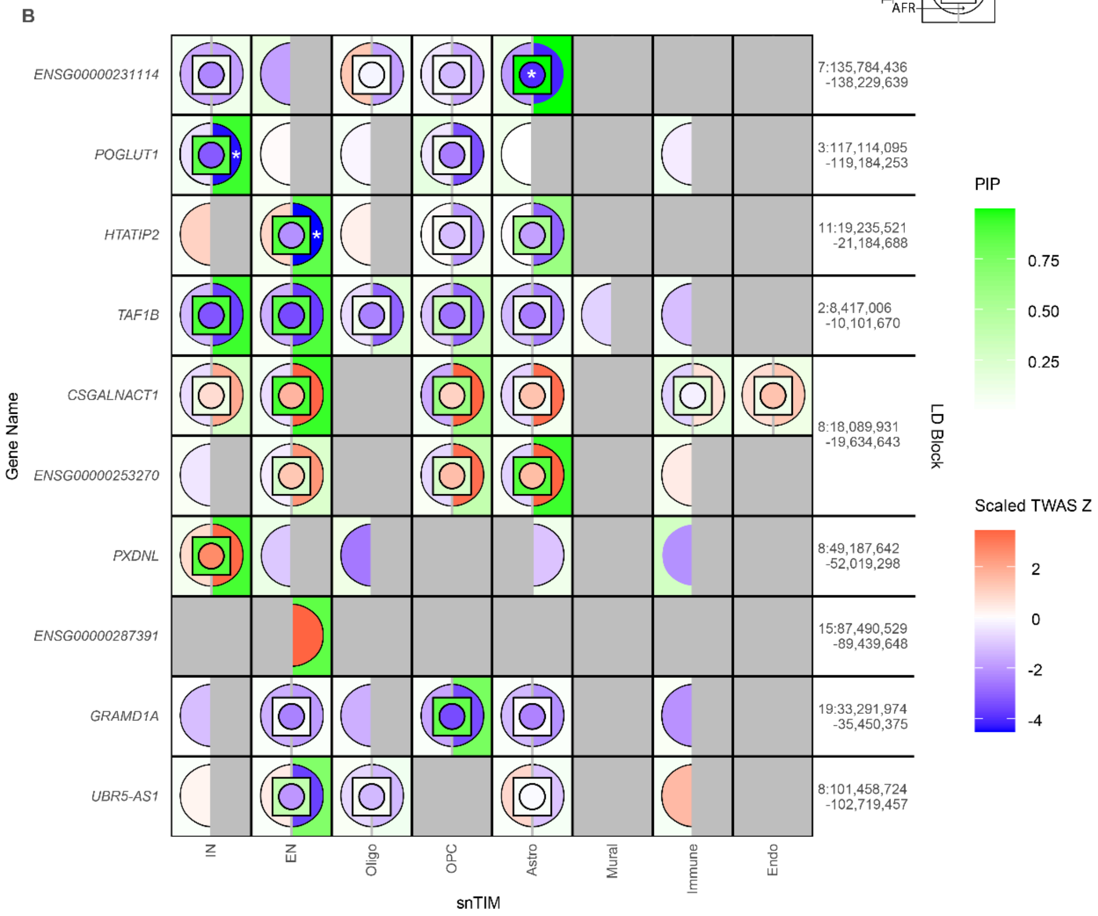

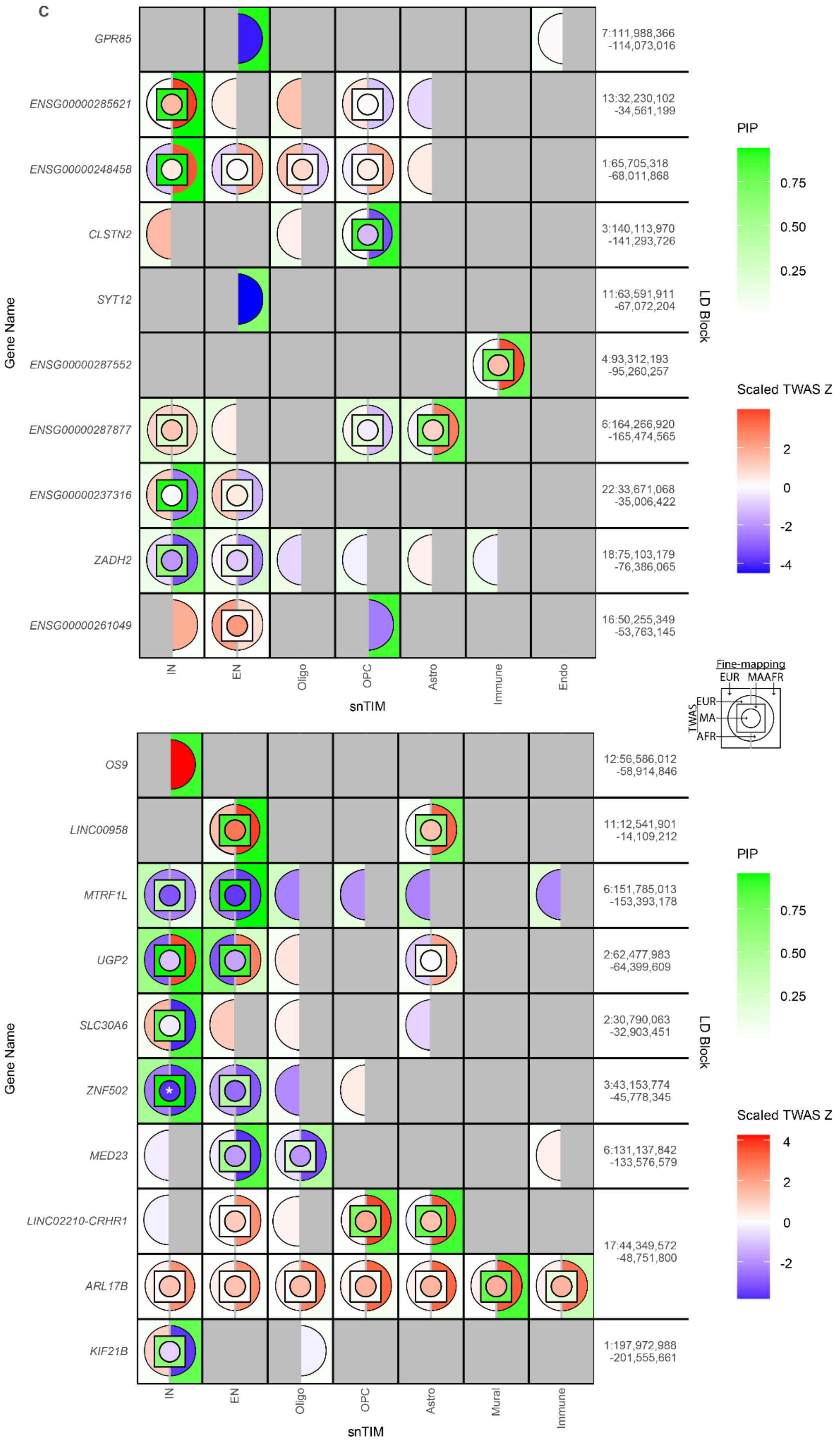
C
GPR85
ENSG00000285621
ENSG00000248458
CLSTN2
SYT12
ENSG00000287552
ENSG00000287877
ENSG00000237316
ZADH2
ENSG00000261049
Gene Name
7:111,988,366 -114,073,016
13:32,230,102 -34,561,199
1:65,705,318 -68,011,868
3:140,113,970 -141,293,726
11:63,591,911 -67,072,204
4:93,312,193 -95,260,257
6:164,266,920 -165,474,565
22:33,671,068 -35,006,422
18:75,103,179 -76,386,065
16:50,255,349 -53,763,145
LD Block
IN
EN
Oligo
OPC
Astro
Immune
Endo
snTIM
PIP
0.75
0.50
0.25
Scaled TWAS Z
2
0
-2
-4
Fine-mapping
EUR
MA
AFR
TWAS
EUR
MA
AFR
OS9
LINC00958
MTRF1L
UGP2
SLC30A6
ZNF502
MED23
LINC02210-CRHR1
ARL17B
KIF21B
Gene Name
12:56,586,012 -58,914,846
11:12,541,901 -14,109,212
6:151,785,013 -153,393,178
2:62,477,983 -64,399,609
2:30,790,063 -32,903,451
3:43,153,774 -45,778,345
6:131,137,842 -133,576,579
17:44,349,572 -48,751,800
1:197,972,988 -201,555,661
LD Block
IN
EN
Oligo
OPC
Astro
Mural
Immune
snTIM
PIP
0.75
0.50
0.25
Scaled TWAS Z
4
2
0
-2

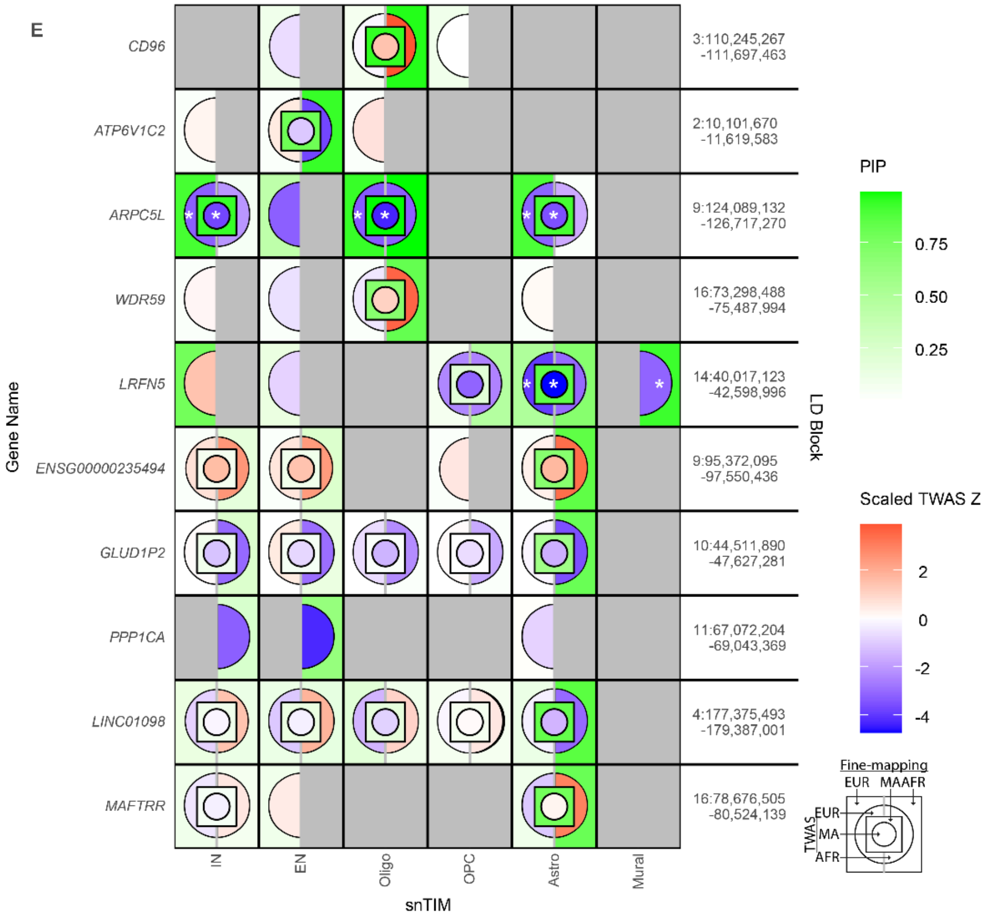
E
CD96
ATP6V1C2
ARPC5L
WDR59
LRFN5
ENSG00000235494
GLUD1P2
PPP1CA
LINC01098
MAFTRR
Gene Name
3:110,245,267 -111,697,463
2:10,101,670 -11,619,583
9:124,089,132 -126,717,270
16:73,298,488 -75,487,994
14:40,017,123 -42,598,996
9:95,372,095 -97,550,436
10:44,511,890 -47,627,281
11:67,072,204 -69,043,369
4:177,375,493 -179,387,001
16:78,676,505 -80,524,139
LD Block
IN
EN
Oligo
OPC
Astro
Mural
snTIM
PIP
0.75
0.50
0.25
Scaled TWAS Z
2
0
-2
-4
Fine-mapping
EUR
MA
AFR
TWAS
EUR
MA
AFR

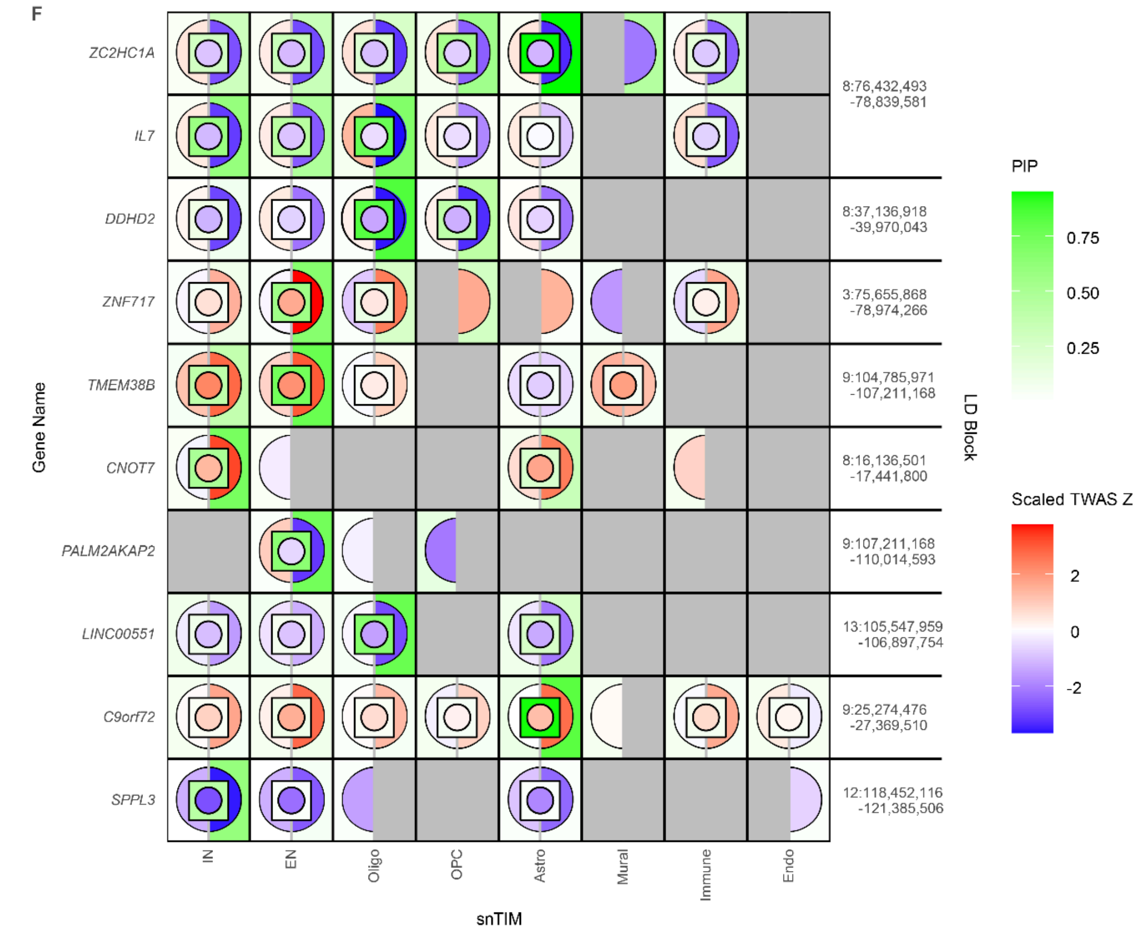
F
ZC2HC1A
IL7
DDHD2
ZNF717
TMEM38B
CNOT7
PALM2AKAP2
LINC00551
C9orf72
SPPL3
Gene Name
8:76,432,493 -78,839,581
8:37,136,918 -39,970,043
3:75,655,868 -78,974,266
9:104,785,971 -107,211,168
8:16,136,501 -17,441,800
9:107,211,168 -110,014,593
13:105,547,959 -106,897,754
9:25,274,476 -27,369,510
12:118,452,116 -121,385,506
LD Block
IN
EN
Oligo
OPC
Astro
Mural
Immune
Endo
snTIM
PIP
0.75
0.50
0.25
Scaled TWAS Z
2
0
-2

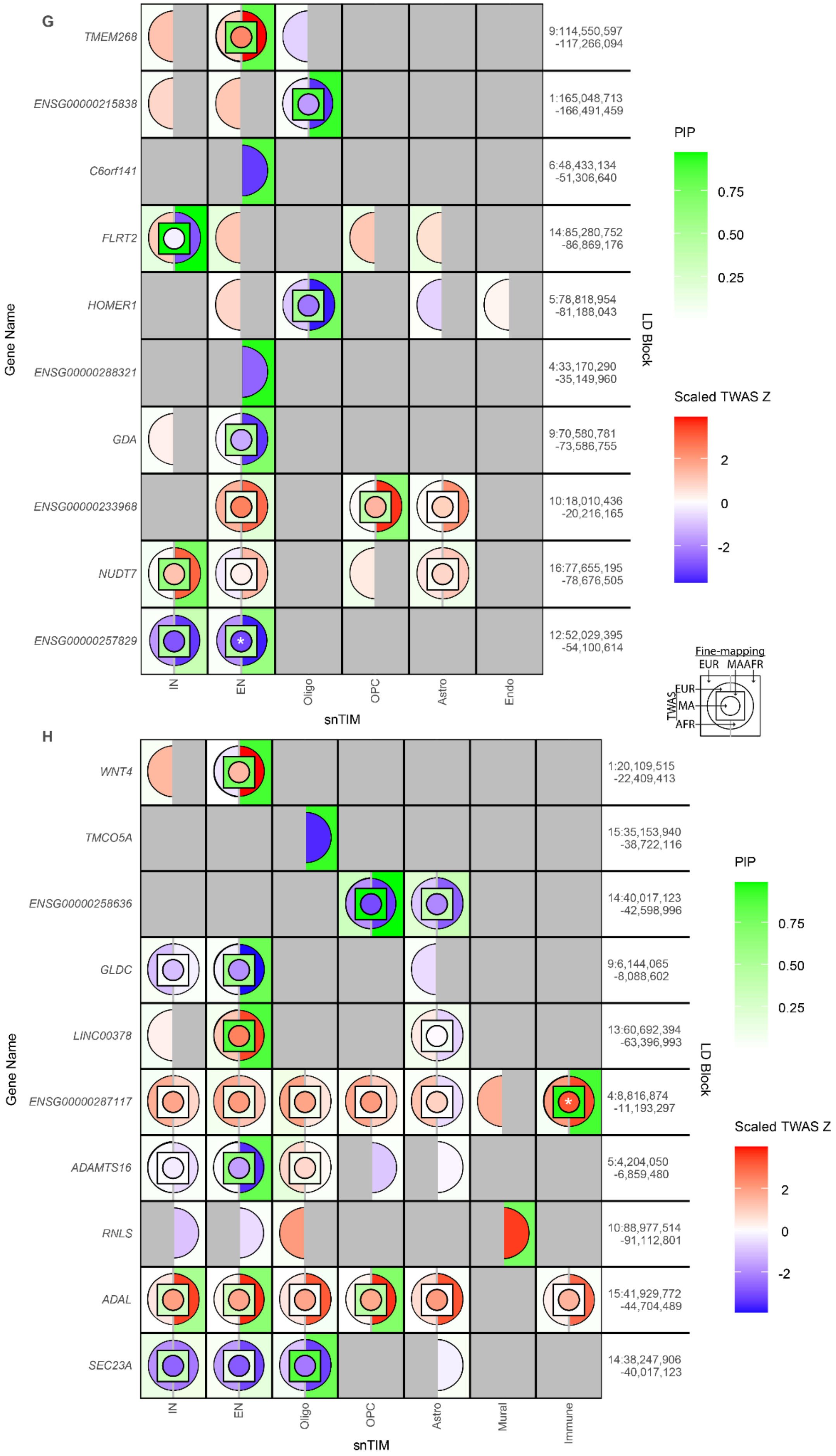
G
Gene Name
TMEM268
ENSG00000215838
C6orf141
FLRT2
HOMER1
ENSG00000288321
GDA
ENSG00000233968
NUDT7
ENSG00000257829
IN
EN
Oligo
OPC
Astro
Endo
snTIM
9:114,550,597 -117,266,094
1:165,048,713 -166,491,459
6:48,433,134 -51,306,640
14:85,280,752 -86,869,176
5:78,818,954 -81,188,043
4:33,170,290 -35,149,960
9:70,580,781 -73,586,755
10:18,010,436 -20,216,165
16:77,655,195 -78,676,505
12:52,029,395 -54,100,614
LD Block
PIP
0.75
0.50
0.25
Scaled TWAS Z
2
0
-2
Fine-mapping
EUR MA AFR
TWAS
EUR
MA
AFR
H
Gene Name
WNT4
TMCO5A
ENSG00000258636
GLDC
LINC00378
ENSG00000287117
ADAMTS16
RNLS
ADAL
SEC23A
IN
EN
Oligo
OPC
Astro
Mural
Immune
snTIM
1:20,109,515 -22,409,413
15:35,153,940 -38,722,116
14:40,017,123 -42,598,996
9:6,144,065 -8,088,602
13:60,692,394 -63,396,993
4:8,816,874 -11,193,297
5:4,204,050 -6,859,480
10:88,977,514 -91,112,801
15:41,929,772 -44,704,489
14:38,247,906 -40,017,123
LD Block
PIP
0.75
0.50
0.25
Scaled TWAS Z
2
0
-2

**Supplementary Fig. 14 | MAFOCUS plots for NPD/NDDs.** Bi-ancestral (EUR and AFR) fine-mapping of BD (**A**), SCZ (**B**), AD (**C**), Insomnia (**D**), MDD (**E**), Migraines (**F**), Anxiety (**G**), PTSD (**H**). The top 10 trait-associated genes in the AFR I-snTWAS analysis are visualized across class level snTIMs. Z-scores are scaled across all genes within each population. AUD is visualized in Fig. 5E.

Supplementary Fig. 15

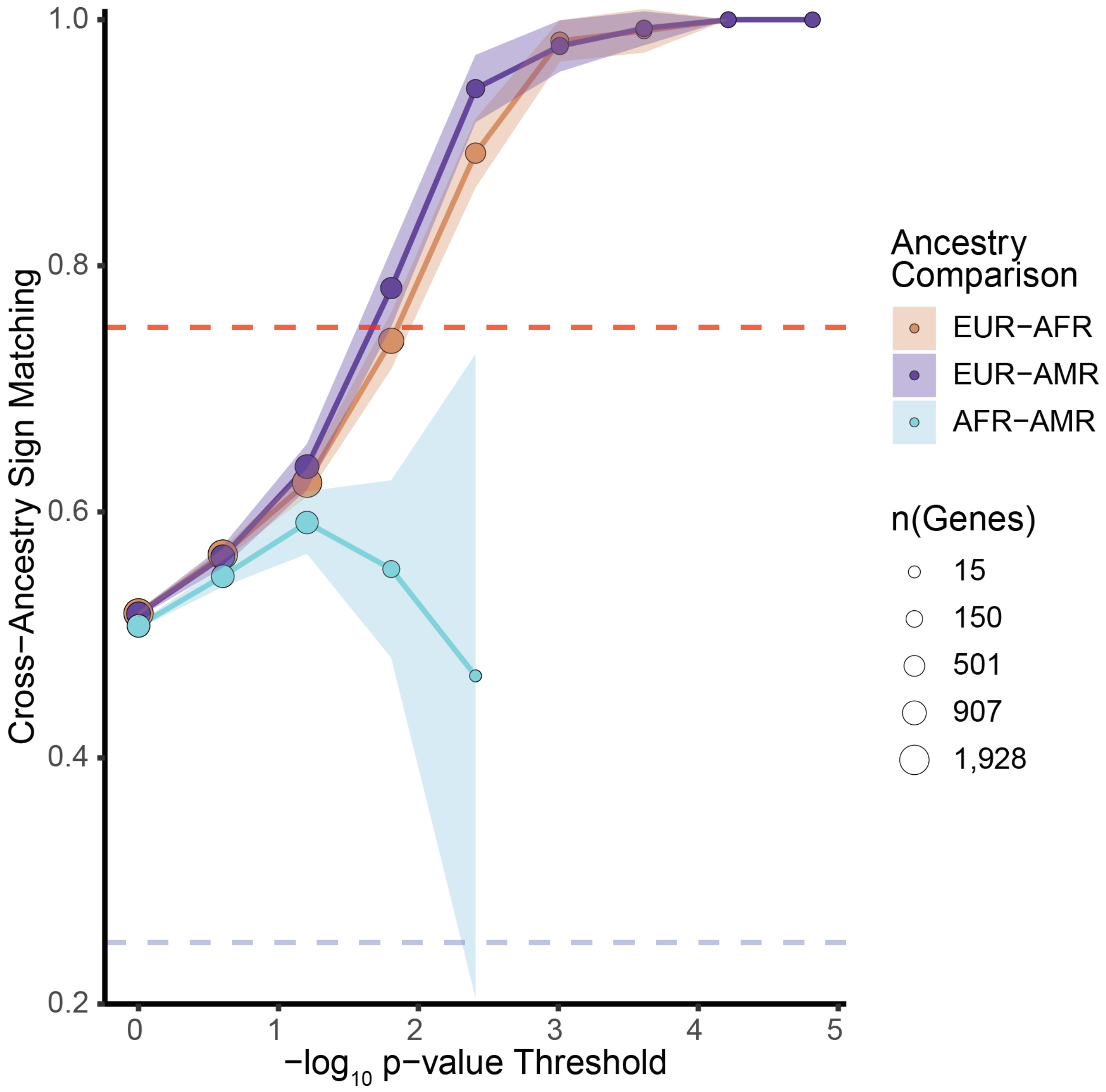

**Supplementary Fig. 15 | Conservation of cross-ancestry concordance of GReX-PheWAS associations among top trait-gene-cell-type combinations.** PheWAS was performed for all genes found to have a significant (Bonferroni-adjusted p-value ≤ 0.05) association in the I-snTWAS analysis spanning all snTIMs and ancestries (EUR, AFR and AMR). For each pairwise ancestry combination, trait-specific PheWAS associations were binned (x-axis) based on the maximum p-value of the shared gene-cell-type combinations (n=1,757, 880 and 681 in EUR-AFR, EUR-AMR, and AFR-AMR, respectively); only phecodes with at least 500 cases and 500 controls were considered. Consequently, the percentage of associations with concordant effect size signs for each ancestry pairwise combination is visualized among increasing p-value significance thresholds. Shaded region around each line represents the 95% confidence interval. Size of the point represents the number of gene-cell-type combinations at that threshold. To visualize a specific p-value threshold, we require a minimum number of observations corresponding to 2% of the applicable gene-cell-type combinations (36, 18 and 14 for EUR-AFR, EUR-AMR and AFR-AMR, respectively). Please note that we are underpowered for the AFR-AMR comparison.

Supplementary Fig. 16

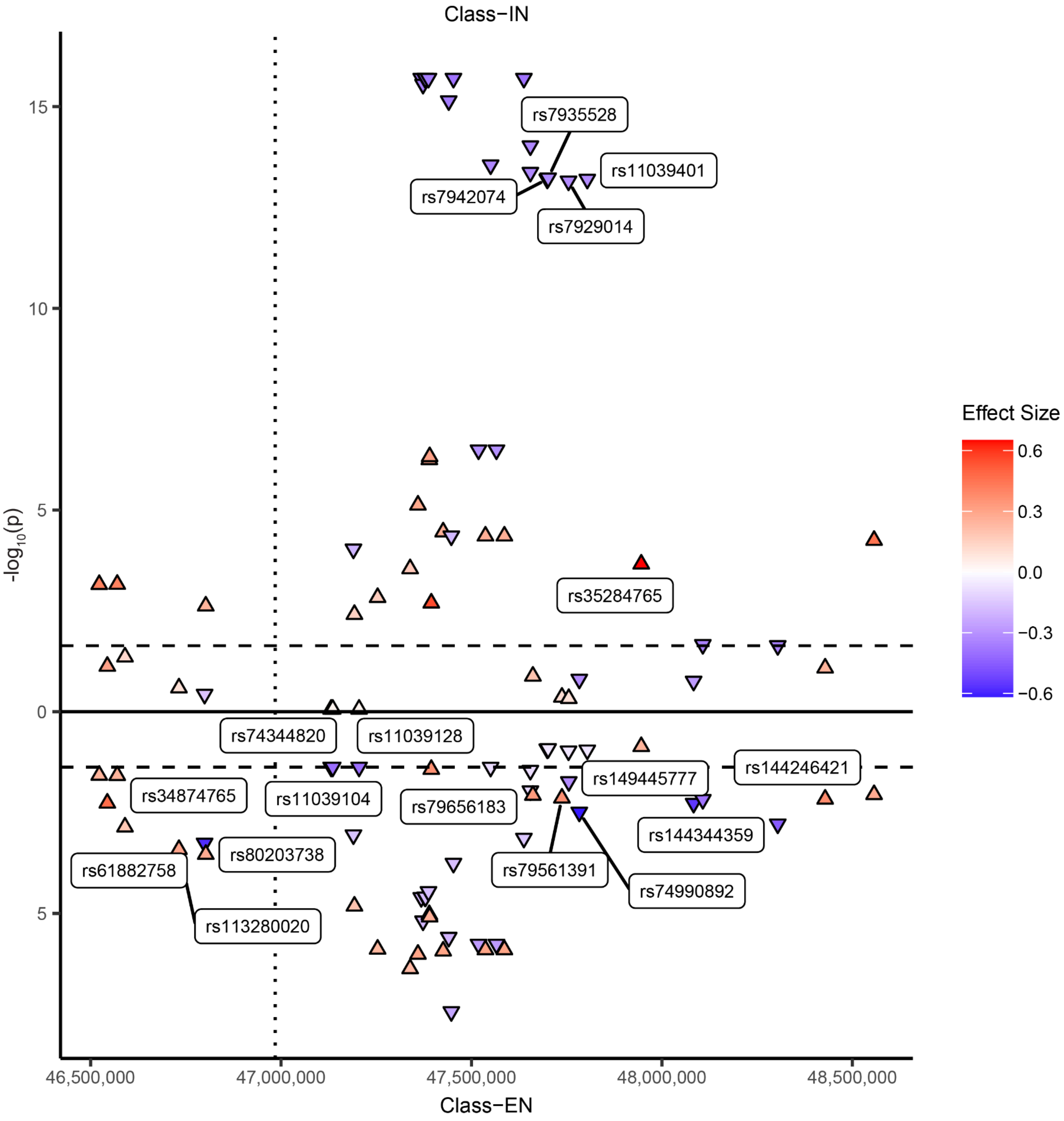

**Supplementary Fig. 16 | *CELF1* sn-eQTLs for inhibitory and excitatory neurons.** Sn-eQTLs for *CELF1* across 2 cell-types. Cell-types were chosen based on the differing association of cell-type specific *CELF1* GReX with various phenotypes. Labeled points indicate SNPs in which there is a significant (FDR[33] ≤ 0.05; threshold for significance is indicated by horizontal dashed lines) effect in one cell-type but not the other. No SNPs contained significant effects in opposite directions for the two cell-types. Plotted SNPs were selected using a superset of SNP predictors in both *CELF1* snTIMs. Upward arrow indicates positive effect size and downward arrow indicates negative effect size. The vertical dotted line divides the LD blocks (utilizing EUR LD blocks as previously described[113]).

### Supplementary Fig. 17

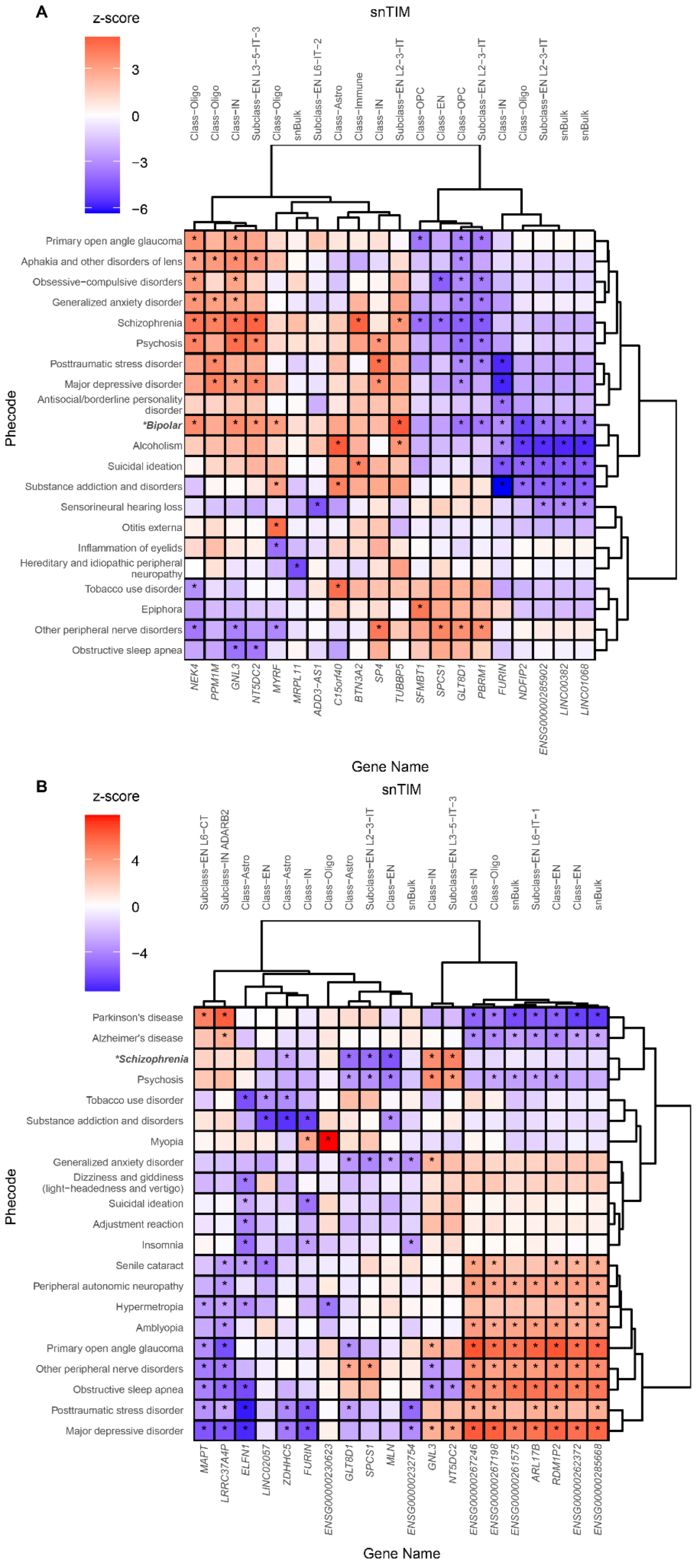

C

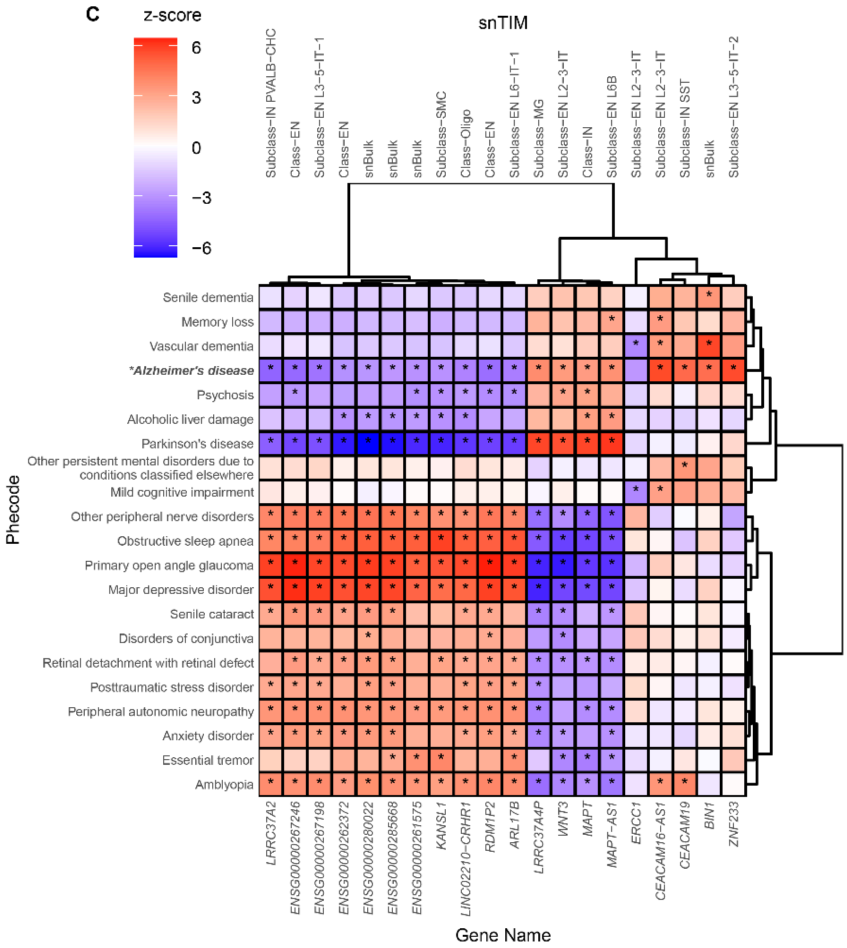
z-score
6
3
0
−3
−6
snTIM
Subclass−IN PVALB-CHC
Class−EN
Subclass−EN L3−5−IT−1
Class−EN
snBulk
snBulk
snBulk
Subclass−SMC
Class−Oligo
Class−EN
Subclass−EN L6−IT−1
Subclass−MG
Subclass−EN L2−3−IT
Class−IN
Subclass−EN L6B
Subclass−EN L2−3−IT
Subclass−EN L2−3−IT
Subclass−IN SST
snBulk
Subclass−EN L3−5−IT−2
Senile dementia
Memory loss
Vascular dementia
*Alzheimer's disease
Psychosis
Alcoholic liver damage
Parkinson's disease
Other persistent mental disorders due to conditions classified elsewhere
Mild cognitive impairment
Other peripheral nerve disorders
Obstructive sleep apnea
Primary open angle glaucoma
Major depressive disorder
Senile cataract
Disorders of conjunctiva
Retinal detachment with retinal defect
Posttraumatic stress disorder
Peripheral autonomic neuropathy
Anxiety disorder
Essential tremor
Amblyopia
Phecode
LRRC37A2
ENSG00000267246
ENSG00000267198
ENSG00000262372
ENSG00000280022
ENSG00000285668
ENSG00000261575
KANSL1
LINC02210−CRHR1
RDM1P2
ARL17B
LRRC37A4P
WNT3
MAPT
MAPT−AS1
ERCC1
CEACAM16−AS1
CEACAM19
BIN1
ZNF233
Gene Name

D

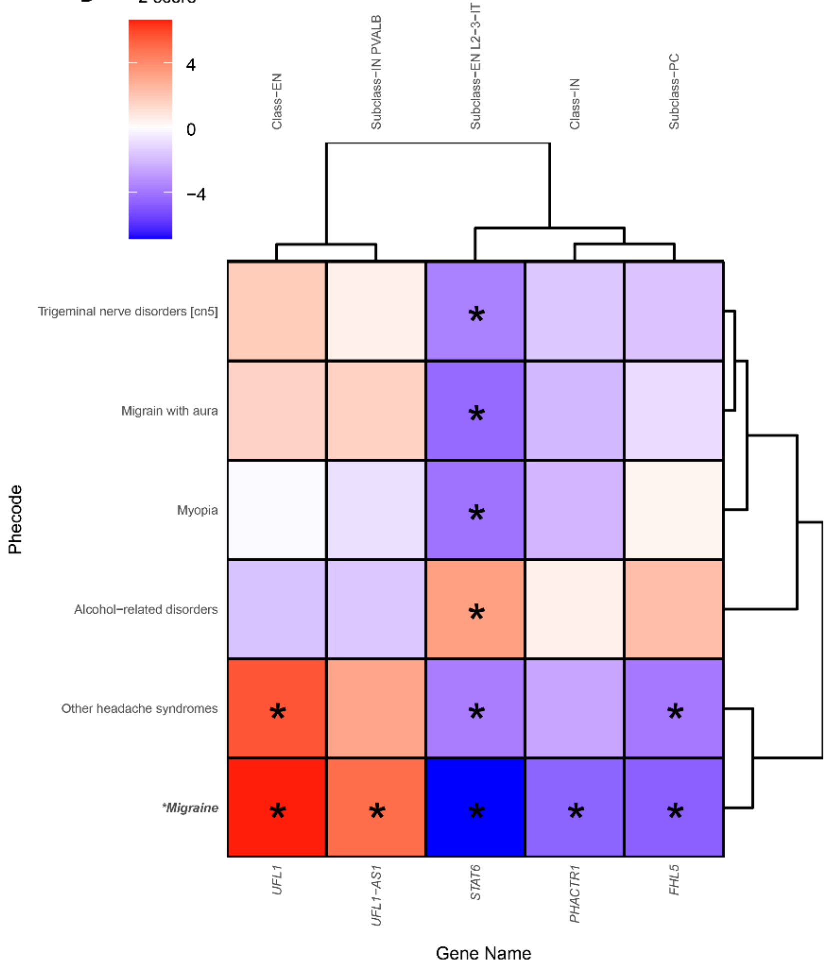
z-score
4
0
−4
snTIM
Class−EN
Subclass−IN PVALB
Subclass−EN L2−3−IT
Class−IN
Subclass−PC
Trigeminal nerve disorders [cn5]
Migrain with aura
Myopia
Alcohol−related disorders
Other headache syndromes
*Migraine
Phecode
UFL1
UFL1−AS1
STAT6
PHACTR1
FHL5
Gene Name

E

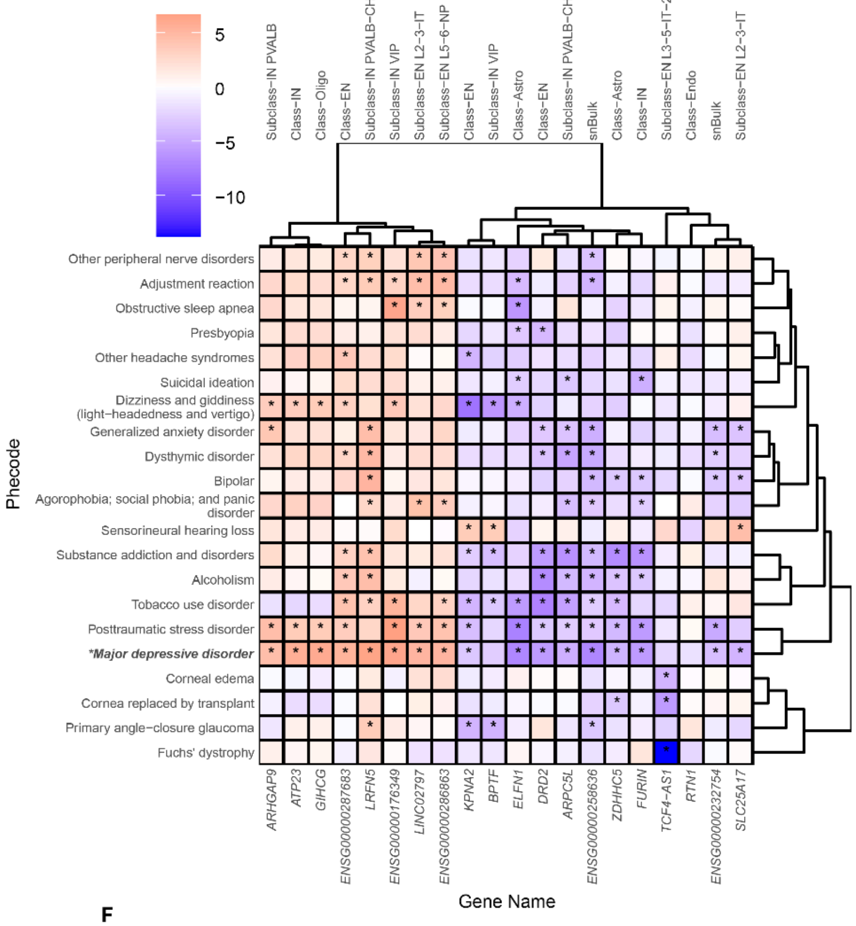

z-score
5
0
−5
−10
snTIM
Subclass−IN PVALB
Class−IN
Class−Oligo
Class−EN
Subclass−IN PVALB−CHC
Subclass−IN VIP
Subclass−EN L2−3−IT
Subclass−EN L5−6−NP
Class−EN
Subclass−IN VIP
Class−Astro
Class−EN
Subclass−IN PVALB−CHC
snBulk
Class−Astro
Class−IN
Subclass−EN L3−5−IT−2
Class−Endo
snBulk
Subclass−EN L2−3−IT
Phecode
Other peripheral nerve disorders
Adjustment reaction
Obstructive sleep apnea
Presbyopia
Other headache syndromes
Suicidal ideation
Dizziness and giddiness (light−headedness and vertigo)
Generalized anxiety disorder
Dysthymic disorder
Bipolar
Agorophobia; social phobia; and panic disorder
Sensorineural hearing loss
Substance addiction and disorders
Alcoholism
Tobacco use disorder
Posttraumatic stress disorder
*Major depressive disorder
Corneal edema
Cornea replaced by transplant
Primary angle−closure glaucoma
Fuchs' dystrophy
ARHGAP9
ATP23
GIHCG
ENSG00000287683
LRFN5
ENSG00000176349
LINC02797
ENSG00000286863
KPNA2
BPTF
ELFN1
DRD2
ARPC5L
ENSG00000258636
ZDHHC5
FURIN
TCF4−AS1
RTN1
ENSG00000232754
SLC25A17
Gene Name

F

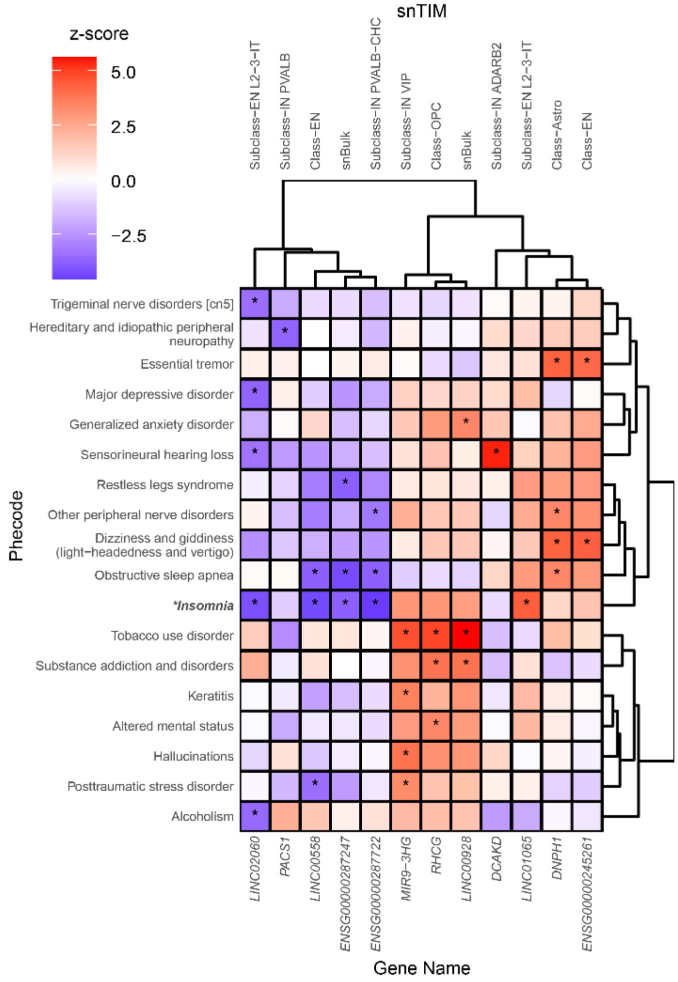

z-score
5.0
2.5
0.0
−2.5
snTIM
Subclass−EN L2−3−IT
Subclass−IN PVALB
Class−EN
snBulk
Subclass−IN PVALB−CHC
Subclass−IN VIP
Class−OPC
snBulk
Subclass−IN ADARB2
Subclass−EN L2−3−IT
Class−Astro
Class−EN
Phecode
Trigeminal nerve disorders [cn5]
Hereditary and idiopathic peripheral neuropathy
Essential tremor
Major depressive disorder
Generalized anxiety disorder
Sensorineural hearing loss
Restless legs syndrome
Other peripheral nerve disorders
Dizziness and giddiness (light−headedness and vertigo)
Obstructive sleep apnea
*Insomnia
Tobacco use disorder
Substance addiction and disorders
Keratitis
Altered mental status
Hallucinations
Posttraumatic stress disorder
Alcoholism
LINC02060
PACS1
LINC00558
ENSG00000287247
ENSG00000287722
MIR9−3HG
RHCG
LINC00928
DCAKD
LINC01065
DNPH1
ENSG00000245261
Gene Name

**Supplementary Fig. 17 | Clustering of top PheWAS associations in S-snTWAS.** PheWAS for the top 20 gene-cell-type combinations ranked from the S-snTWAS for each trait; each gene is represented by the cell-type with the lowest association p-value. In addition to each trait (mapped to the bolded italicized phecode), the top 20 (if at least 20 distinct phecodes are significant amongst the op 20 gene-cell-type combinations) phecodes among all phecode categories ranked by association p value are visualized. Ward's hierarchical agglomerative clustering was performed with Ward's criterion preservation. We describe the following traits in each panel: BD (**A**), SCZ (**B**), AD (**C**), Migraines (**D**), MDD (**E**), Insomnia (**F**). AUD is described within fig. 6C. ADHD, ALS, Anorexia, MS, and PD are excluded due to few cases in the MVP. PTSD and Anxiety are excluded due to few significant S-snTWAS associations.

Supplementary Fig. 18

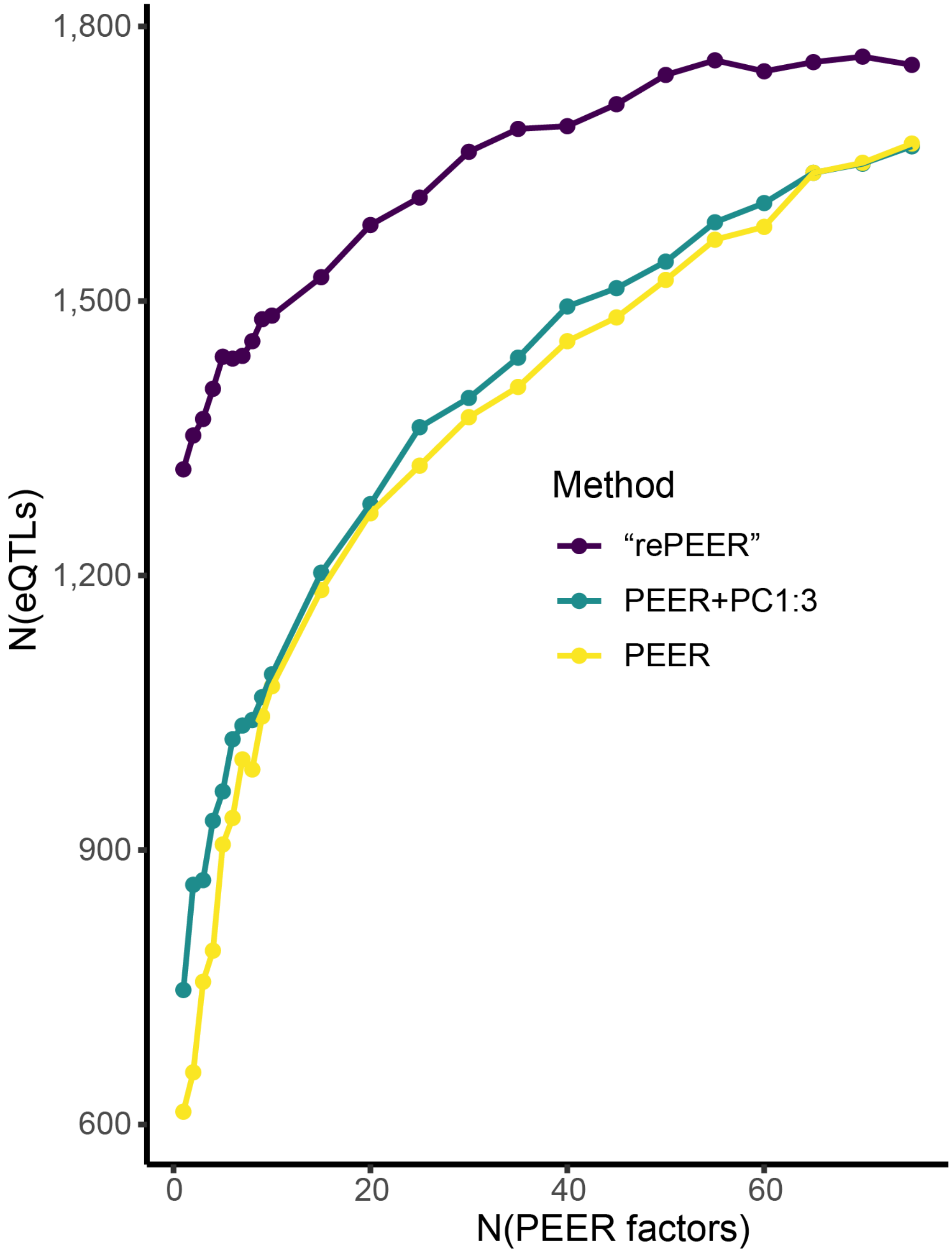

**Supplementary Fig. 18 | Comparison of batch effect correction methods.** To assess the impact of different batch effect correction methods on overall power, we utilized the FACS-MG cohort. "rePEER" refers to the method to perform PEER on each batch separately and again altogether after aggregation of all PEER-residualized expression data. "PEER" refers to the method to just perform PEER on all expression data aggregated across all batches. "PEER+PC1:3" refers to the method to perform PEER on all expression data aggregated across all batches with genotyping PCs 1, 2, and 3 supplied to the PEER algorithm. Each point represents the number of significant eQTLs

(FDR[33]-adjusted p-value ≤ 0.05) found at each selected number of PEER factors used in each method (using the second round of PEER for the “rePEER” method).

Supplementary Fig. 19

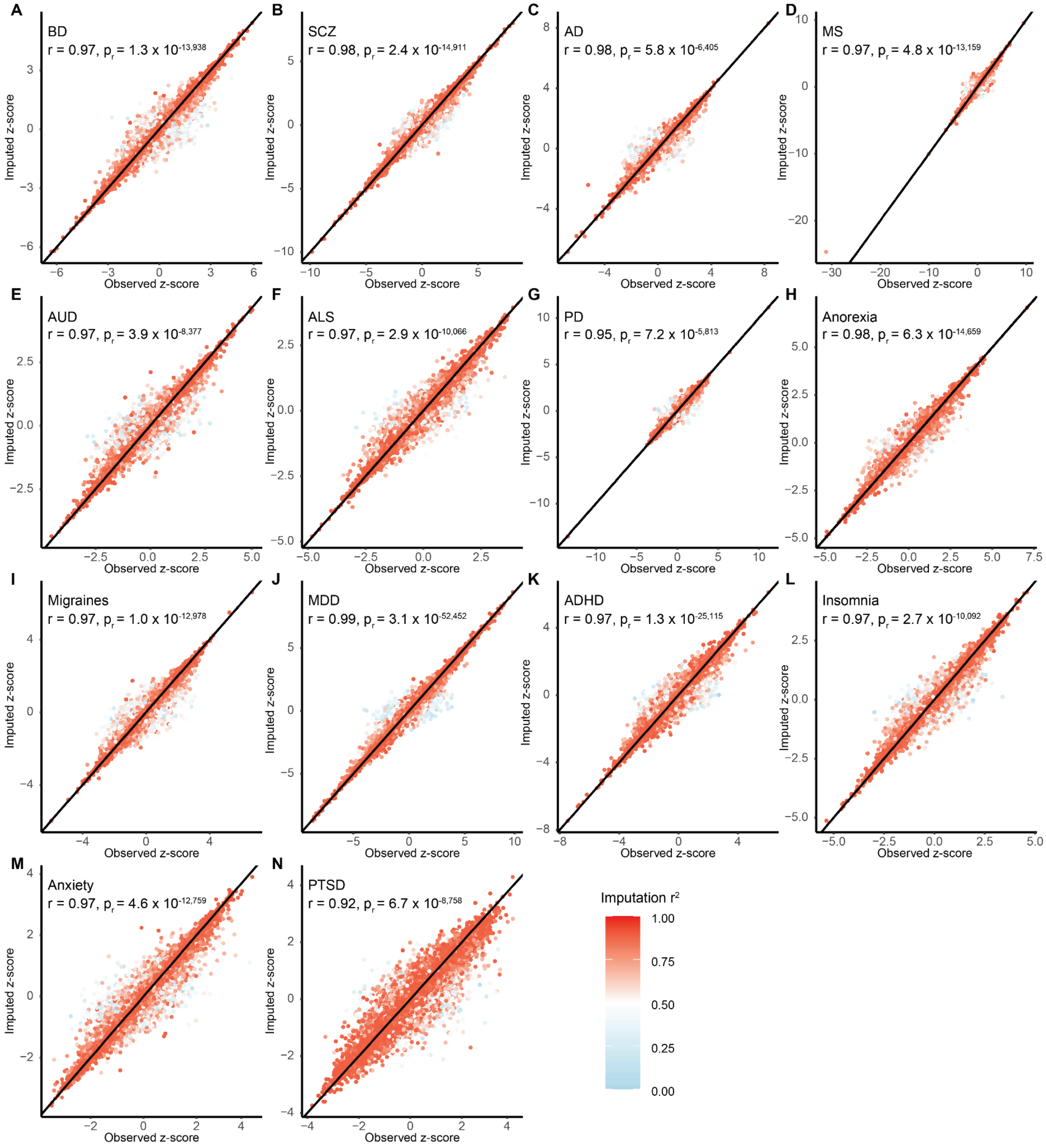

**Supplementary Fig. 19 | GWAS imputation accuracy.** We randomly selected SNPs for each GWAS (Table S17) (mean number of selected SNPs was 23,018 with a standard deviation of 14,334). For all selected SNPs, we performed GWAS imputation as if the SNP was missing so that we could observe the accuracy of GWAS imputation on missing data. GWAS imputation is described for each included trait: BD (**A**), SCZ (**B**), AD (**C**), MS (**D**), AUD (**E**), ALS (**F**), PD (**G**), Anorexia (**H**), Migraines (**I**), MDD (**J**), ADHD (**K**), Insomnia (**L**), Anxiety (**M**), PTSD (**N**). Every plot is annotated with the Pearson's correlation coefficient (r) and p-value. Color of the point indicates the imputation $r^2$. Only missing SNPs with imputation $r^2 \geq 0.7$ were retained for further analysis. Black diagonal line is a reference line with a slope of 1.

### SUPPLEMENTARY NOTES

#### Other Acknowledgments

##### VA Million Veteran Program: Core Acknowledgements for Publications May 2024

**MVP Program Office**
Sumitra Muralidhar, Ph.D., Program Director[1]
Jennifer Moser, Ph.D., Associate Director, Scientific Programs[1]
Jennifer E. Deen, B.S., Associate Director, Cohort & Public Relations[1]

**MVP Executive Committee**
Co-Chair: Philip S. Tsao, Ph.D.[2]
Co-Chair: Sumitra Muralidhar, Ph.D.[1]
J. Michael Gaziano, M.D., M.P.H.[3]
Elizabeth Hauser, Ph.D.[4]
Amy Kilbourne, Ph.D., M.P.H.[5]
Michael Matheny, M.D., M.S., M.P.H.[6]
Dave Oslin, M.D.[7]
Deepak Voora, MD[4]

**MVP Co-Principal Investigators**
J. Michael Gaziano, M.D., M.P.H.[3]
Philip S. Tsao, Ph.D.[2]

**MVP Core Operations**
Jessica V. Brewer, M.P.H., Director, MVP Cohort Operations[3]
Mary T. Brophy M.D., M.P.H., Director, VA Central Biorepository[3]
Kelly Cho, M.P.H, Ph.D., Director, MVP Phenomics[3]
Lori Churby, B.S., Director, MVP Regulatory Affairs[2]
Scott L. DuVall, Ph.D., Director, VA Informatics and Computing Infrastructure (VINCI)[8]
Saiju Pyarajan Ph.D., Director, Data and Computational Sciences[3]
Robert Ringer, Pharm.D., Director, VA Albuquerque Central Biorepository[9]
Luis E. Selva, Ph.D., Director, MVP Biorepository Coordination[3]
Shahpoor (Alex) Shayan, M.S., Director, MVP PRE Informatics[3]
Brady Stephens, M.S., Principal Investigator, MVP Information Center[10]
Stacey B. Whitbourne, Ph.D., Director, MVP Cohort Development and Management[3]

**MVP Publications and Presentations Committee**
Co-Chair: Themistocles L. Assimes, M.D., Ph. D[2]
Co-Chair: Adriana Hung, M.D.; M.P.H[6]

Co-Chair: Henry Kranzler, M.D.[7]

**MVP Affilations**
1: US Department of Veterans Affairs, 810 Vermont Avenue NW, Washington, DC 20420
2: VA Palo Alto Health Care System, 3801 Miranda Avenue, Palo Alto, CA 94304
3: VA Boston Healthcare System, 150 S. Huntington Avenue, Boston, MA 02130
4: Durham VA Medical Center, 508 Fulton Street, Durham, NC 27705
5: VA HSR&D, 2215 Fuller Road, Ann Arbor, MI 48105
6: VA Tennessee Valley Healthcare System, 1310 24th Ave. South, Nashville, TN 37212
7: Philadelphia VA Medical Center, 3900 Woodland Avenue, Philadelphia, PA 19104
8: VA Salt Lake City Health Care System, 500 Foothill Drive, Salt Lake City, UT 84148
9: New Mexico VA Health Care System, 1501 San Pedro Drive SE, Albuquerque, NM 87108
10: Canandaigua VA Medical Center, 400 Fort Hill Avenue, Canandaigua, NY 14424

#### PsychAD Acknowledgements

**PsychAD Consortium Authors**
Aram Hong (1, 4, 6, 7); Athan Z. Li (10, 12); Biao Zeng (1, 4, 6, 7); Chenfeng He (9, 12); Chirag Gupta (9, 12); Christian Porras (1, 4, 6, 7); Clara Casey (1, 4, 6, 7); Colleen A. McClung (18); Collin Spencer (1, 4, 6, 7); Daifeng Wang (9, 10, 12); David A. Bennett (19); David Burstein (1, 2, 4, 6, 7, 8); Deepika Mathur (1, 4, 6, 7); Donghoon Lee (1, 4, 6, 7); Fotios Tsetsos (1, 2, 4, 6, 7); Gabriel E. Hoffman (1, 2, 4, 6, 7, 8); Genadi Ryan (13, 17); Georgios Voloudakis (1, 2, 3, 4, 6, 7, 8); Hui Yang (1, 4, 6, 7); Jaroslav Bendl (1, 4, 6, 7); Jerome J. Choi (11, 12); John F. Fullard (1, 4, 6, 7); Kalpana H. Arachchilage (9, 12); Karen Therrien (1, 4, 6, 7); Kiran Girdhar (1, 4, 6, 7); Lars J. Jensen (21); Lisa L. Barnes (19); Logan C. Dumitrescu (22, 23); Lyra Sheu (1, 4, 6, 7); Madeline R. Scott (18); Marcela Alvia (1, 4, 6, 7); Marios Anyfantakis (1, 4, 6, 7); Maxim Signaevsky (6, 7); Mikaela Koutrouli (1, 4, 6, 7, 21); Milos Pjanic (1, 4, 6, 7); Monika Ahirwar (13, 17); Nicolas Y. Masse (1, 4, 6, 7); Noah Cohen Kalafut (10, 12); Panos Roussos (1, 2, 4, 6, 7, 8); Pavan K. Auluck (20); Pavel Katsel (6); Pengfei Dong (1, 4, 6, 7); Pramod B. Chandrashekar (9, 12); Prashant N.M. (1, 4, 6, 7); Rachel Bercovitch (1, 4, 6, 7); Roman Kosoy (1, 4, 6, 7); Sanan Venkatesh (1, 2, 4, 6, 7); Saniya Khullar (9, 12); Sayali A. Alatkar (10, 12); Seon Kinrot (1, 4, 6, 7); Stathis Argyriou (1, 4, 6, 7); Stefano Marenco (20); Steven Finkbeiner (13, 14, 15, 16, 17); Steven P. Kleopoulos (1, 4, 6, 7); Tereza Clarence (1, 4, 6, 7); Timothy J. Hohman (22, 23); Ting Jin (9, 12); Vahram Haroutunian (5, 6, 7, 8); Vivek G. Ramaswamy (13, 17); Xiang Huang (12); Xinyi Wang (1, 4, 6, 7); Zhenyi Wu (1, 4, 6, 7); Zhiping Shao (1, 4, 6, 7)

**PsychAD Consortium Affiliations**
1: Center for Disease Neurogenomics, Icahn School of Medicine at Mount Sinai, New York, NY, USA
2: Center for Precision Medicine and Translational Therapeutics, James J. Peters VA Medical Center, Bronx, NY, USA
3: Department of Artificial Intelligence and Human Health, Icahn School of Medicine at Mount Sinai, New York, NY, USA
4: Department of Genetics and Genomic Sciences, Icahn School of Medicine at Mount Sinai, New York, NY, USA
5: Department of Neuroscience, Icahn School of Medicine at Mount Sinai, New York, NY, USA

6: Department of Psychiatry, Icahn School of Medicine at Mount Sinai, New York, NY, USA
7: Friedman Brain Institute, Icahn School of Medicine at Mount Sinai, New York, NY, USA
8: Mental Illness Research, Education and Clinical Center VISN2, James J. Peters VA Medical Center, Bronx, NY, USA
9: Department of Biostatistics and Medical Informatics, University of Wisconsin-Madison, Madison, WI, USA
10: Department of Computer Sciences, University of Wisconsin-Madison, Madison, WI, USA
11: Department of Population Health Sciences, University of Wisconsin-Madison, Madison, WI, USA
12: Waisman Center, University of Wisconsin-Madison, Madison, WI, USA
13: Center for Systems and Therapeutics, Gladstone Institutes, San Francisco, CA, USA
14: Department of Neurology, University of California San Francisco, San Francisco, CA, USA
15: Department of Physiology, University of California San Francisco, San Francisco, CA, USA
16: Neuroscience and Biomedical Sciences Graduate Programs, University of California San Francisco, San Francisco, CA, USA
17: Taube/Koret Center for Neurodegenerative Disease Research, Gladstone Institutes, San Francisco, CA, USA
18: Department of Psychiatry, University of Pittsburgh School of Medicine, Pittsburgh, PA, USA
19: Rush Alzheimer's Disease Center and Department of Neurological Sciences, Rush University Medical Center, Chicago, IL, USA
20: Human Brain Collection Core, National Institute of Mental Health-Intramural Research Program, Bethesda, MD, USA
21: Novo Nordisk Foundation Center for Protein Research, Faculty of Health and Medical Sciences, University of Copenhagen, Copenhagen, Denmark
22: Vanderbilt Genetics Institute, Vanderbilt University Medical Center, Nashville, TN, USA
23: Vanderbilt Memory & Alzheimer's Center, Vanderbilt University Medical Center, Nashville, TN, USA
